# Recurrent SARS-CoV-2 Mutations in Immunodeficient Patients

**DOI:** 10.1101/2022.03.02.22271697

**Authors:** Sam AJ Wilkinson, Alex Richter, Anna Casey, Husam Osman, Jeremy D Mirza, Joanne Stockton, Josh Quick, Liz Ratcliffe, Natalie Sparks, Nicola Cumley, Radoslaw Poplawski, Sam Nicholls, Beatrix Kele, Kathryn Harris, The COVID-19 Genomics UK (COG-UK) consortium, Thomas P Peacock, Nicholas J Loman

**Affiliations:** Institute of Microbiology and Infection, School of Biosciences, University of Birmingham, UK, B15 2TT; Institute of Immunology and Immunotherapy (III), College of Medical and Dental Sciences, University of Birmingham, UK, B15 2TT; Queen Elizabeth Hospital, University Hospitals Birmingham, Birmingham, UK, B15 2TH; Virology Department, NHS East and South East London Pathology Partnership, Royal London Hospital, Barts Health NHS Trust, UK, EC1A 7BE; Department of Infectious Disease, Imperial College London, London, UK, W2 1PG

**Author notes:** https://www.cogconsortium.uk - Full list of consortium names and affiliations are in the appendix.

## Abstract

Long-term SARS-CoV-2 infections in immunodeficient patients are an important source of variation for the virus but are understudied. Many case studies have been published which describe one or a small number of long-term infected individuals but no study has combined these sequences into a cohesive dataset. This work aims to rectify this and study the genomics of this patient group through a combination of literature searches as well as identifying new case series directly from the COG-UK dataset. The spike gene receptor binding domain (RBD) and N-terminal domains (NTD) were identified as mutation hotspots. Numerous mutations associated with variants of concern were observed to emerge recurrently. Additionally a mutation in the envelope gene, - T30I was determined to be the most recurrent frequently occurring mutation arising in persistent infections. A high proportion of recurrent mutations in immunodeficient individuals are associated with ACE2 affinity, immune escape, or viral packaging optimisation.

There is an apparent selective pressure for mutations which aid *intra-*host transmission or persistence which are often different to mutations which aid *inter-*host transmission, although the fact that multiple recurrent *de novo* mutations are considered defining for variants of concern strongly indicates that this potential source of novel variants should not be discounted.

## Introduction

Long-term SARS-CoV-2 infections in immunodeficient patients are important, but understudied (Moran et al., 2021). Evolution of viruses during long-term infection is an important source of novel variation and is thought to be a key influence of the evolutionary dynamics of SARS-CoV-2 generally, and the emergence of new variants specifically. Notably Alpha and Omicron, which were responsible for recent epidemic waves globally, are hypothesised by some to have arisen during long-term infections (Msomi et al., 2021; Rambaut et al., 2020). The Alpha variant (B.1.1.7) emerged abruptly with a constellation of novel mutations and a long branch length from its nearest common ancestor in the B.1.1 clade, during a time of extremely high surveillance in the UK (Rambaut et al., 2020). A likely explanation is that the Alpha variant evolved within a single long-term host over a long period before emergence back into the general population. Evolution during long-term infection has been associated with the rapid accumulation of many mutations within a short period (Avanzato et al., 2020; Baang et al., 2021; Choi et al., 2020; Jensen et al., 2021; Karim et al., 2021; Peacock et al., 2021; Riddell et al., 2022). The Beta (B.1.351), Gamma (P.1), and Omicron (B.1.1.529) variants all emerged in similar circumstances to alpha, potentially suggesting that they also emerged from long-term infections.

To better understand evolutionary pressures associated with viral evolution during long-term infections a dataset composed of 168 SARS-CoV-2 genomes associated with 28 patients with a range of conditions that result in immunodeficiency significant enough to prevent rapid viral clearance was compiled to examine the frequency of recurrent mutations. This builds upon previous work performing a similar analysis using case studies which included a total of ten patients (Peacock et al., 2021). This analysis expands on previous studies by utilising a significantly larger dataset which increases the power, also many of the cases included are the alpha variant which have not been discussed in the context of long-term SARS-CoV-2 cases previously and potentially gives insight into future variant emergence, and lastly all genome series’ were analysed using a single analysis pipeline.

## Methods

### Dataset assembly

Patient-associated genome series were selected for inclusion via a literature search for case studies using the following search terms and filters: After 2019, “SARS-CoV-2”, “nCoV-2019”, “Immunodeficient”, “Immunocompromised”, “long-term”, all searches took place between the dates 01/08/2021 and 30/11/2021.

Other genome series were extracted from the COG-UK dataset, a UK-wide genomic surveillance repository (COVID-19 Genomics UK (COG-UK), 2020; Nicholls et al., 2021). Genome series were only included if they met the following criteria: at least two genomes available on either public databases or via a request, evidence of long-term viral infection for a period no less than 28 days (some genome series covered a shorter period but the clinical information met this criterion), clinical information available was sufficient to indicate the nature of the patient’s immune deficiency. For all genome series included in the dataset a Civet report (O’Toole et al., 2021a) was generated using Civet v3.0. These reports confirm that all genomes were the result of long-term infections rather than a super-infection or independent infection events by virtue of individual genomes sharing a recent common ancestor with a step-wise accumulation of mutations over time. A single genome from patient 11 was excluded due to a probable superinfection as described by (Tarhini et al., 2021). Figures were generated for each phylogeny generated with civet using ggtree (Yu et al., 2018) and are included within the supplementary material.

When a genome series was selected for inclusion all genomes were placed within an individual multi-fasta file with a header identifying the patient via an identifier (“pt-1”, pt-2” etc) and the number of days passed since the initial genome available within that genome series (the day 0 genome), in several cases this genome was collected after a lengthy period of active infection but only the time period covered by the genome series was considered in the analysis.

### Mutation Calling of Genomes

Mutation calling was automated with an R script adapted from (Mercatelli et al., 2021) which utilises Nucleotide mummer (NUCmer) (Marçais et al., 2018) for genome alignment to an annotated SARS-CoV-2 reference sequence (Wu et al., 2020) and defines SNPs, insertions, deletions, frameshifts, and inversions relative to this reference sequence (NCBI accession NC_045512.2). One change was made to the annotations of the reference in the case of the ORF1ab polyprotein gene NSP12 where the position was adjusted by a single nucleotide so that all mutation calls would be relative to the reading frame post the ribosomal frame-shift for simplicity; zero mutations were detected in the pre-ribosomal frame-shift region of NSP12 therefore no mutations were incorrectly annotated as a result.

### De-novo Mutation Cumulative Occurrence Analysis Pipeline

Processing of the mutation calls was performed with a Python script (https://github.com/BioWilko/recurrent-sars-cov-2-mutations/blob/main/mutation_call_analysis.py) to investigate *de novo* mutations, which were defined as observed mutations within a genome series which were not present at day 0 of the genome series. A cumulative count of each observed *de novo* mutation (DNM) was performed for each day between 0 and the maximum genome series length (218 days). When a deletion was observed all deletions with a reference position within 18 nucleotides of the reference position of the initial deletion regardless of length or position were clustered as a single region. Ambiguous nucleotides were not considered in mutation calling. The resultant dataframe was finally formatted with an R script and figures generated using ggplot2 (Wickham, 2016).

## Results

The SARS-CoV-2 spike gene (S) demonstrated the greatest number of recurrent mutations in the dataset (Figure 2, Figure 1) with 10 substitutions - S:S13I, S:T95I, S:G142V, S:L452R, S:E484K, S:E484G, S:F486I, S:F490L, S:Q493K, and S:Q498R. The domain where the highest number of DNM occurrences were observed was the Receptor Binding Domain (RBD) with seven, followed by the N-terminal Domain (NTD) with five, and the signal peptide (SP) with one for a total of thirteen. Clustering mutations by AA loci additionally revealed the following sites as notable : S:484, S:501, S:330, and S:440. The domain with the highest number of AA loci with DNMs was the RBD with nine, followed by the NTD with five, and the SP with one. The most frequently occuring DNM was S:E484K with 8 occurrences, when all DNMs at the S:484 loci are clustered (Figure 2); the number of occurrences is increased to twelve clearly demonstrating an enrichment of DNMs at this loci. The DNMs at the loci S:484 consist of: eight S:E484K, two S:E484G, and one each of S:E484Q, and S:E484A. AA loci clustering highlighted the loci S:330, S:440, and S:501 as recurrent for DNMs (≥ two occurrences in the period).

**Figure 1:**
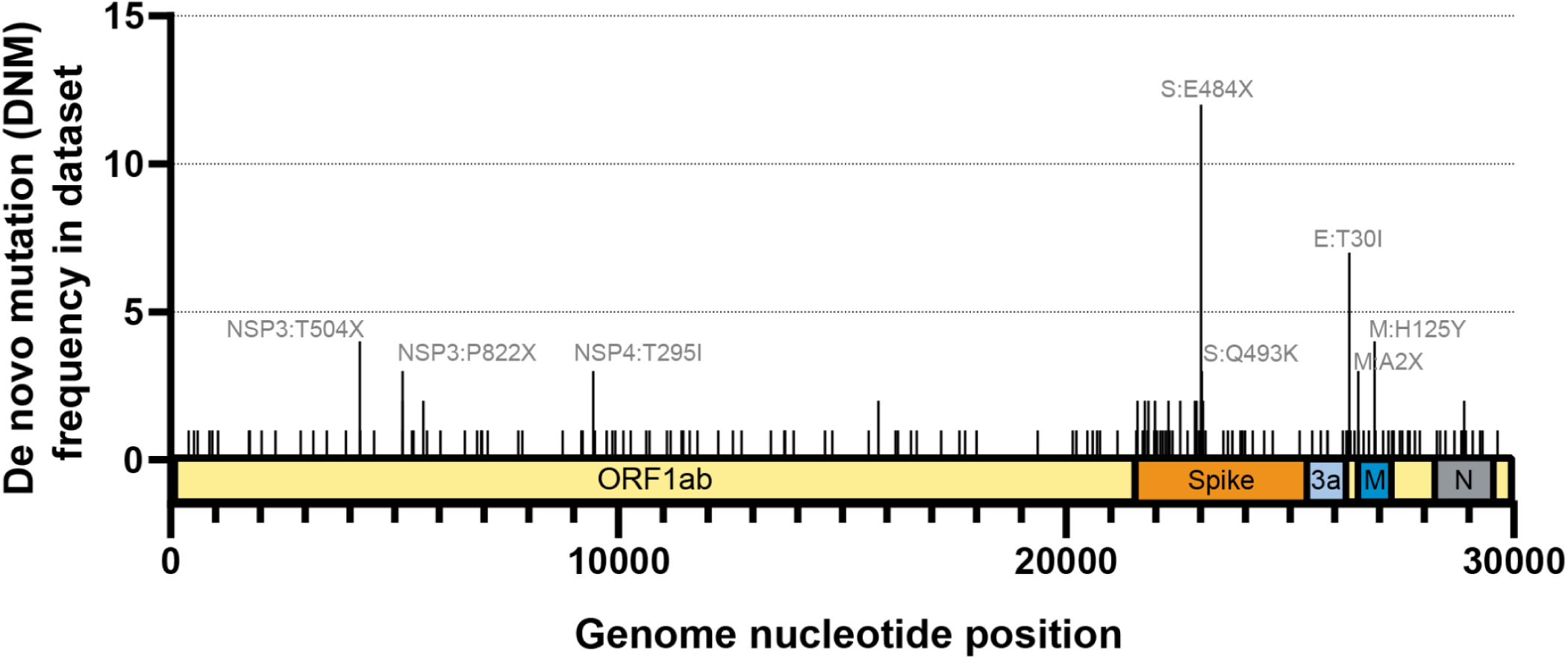
Distribution of *De Novo* Mutations included in this study across the entire SARS-CoV-2 genome. Schematic of SARS-CoV-2 genome with relevant ORFs annotated. DNMs with the highest frequency annotated by amino acid position and substitutions - X indicates multiple amino acids form DNMs at this position.

**Figure 2:**
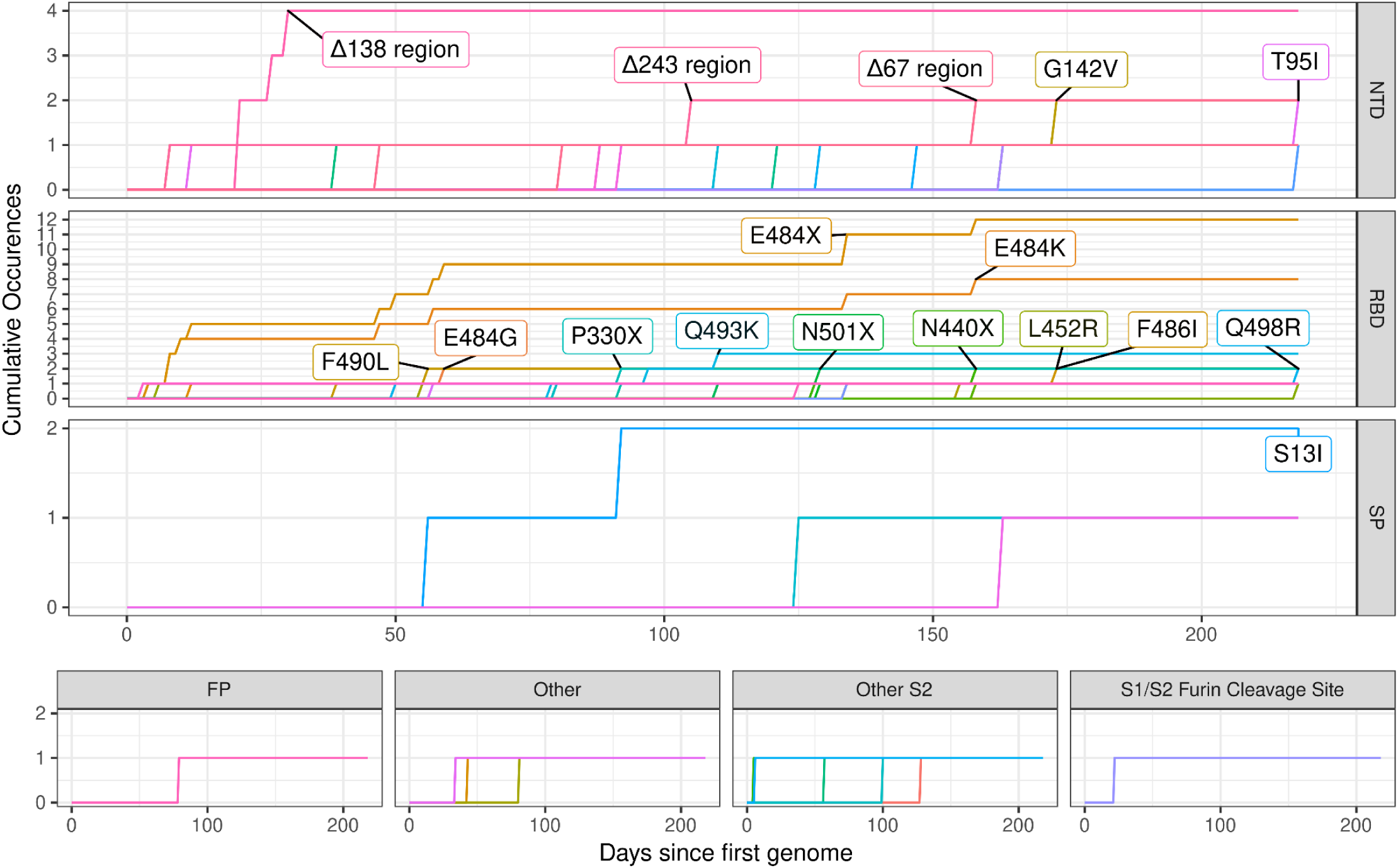
Cumulative occurrences of non-synonymous *de-novo* mutations in S-gene divided by gene domain in 168 genomes obtained from twenty-eight patients. Substitution mutations were clustered by amino acid loci, this is notated with the IUPAC ambiguity code **X** to indicate any possible amino acid. Only loci which were notable when clustered (significant difference with non-clustered equivalent, or loci not highlighted without clustering) were included in the figure. Mutations were observed in the following domains; the S1/S2 Furin Cleavage Site, Fusion Peptide (**FP**), N-terminal Domain (**NTD**), Receptor Binding Domain (**RBD**), Signal Peptide (**SP**), and **Other S2** (Xia, 2021). Deletions (Δ) were clustered within a window of 6 amino acids (AA) regardless of length or position of deletion; full details of the breakdown can be found at https://github.com/BioWilko/recurrent-sars-cov-2-mutations/blob/main/dataset/mutation_calls.csv. The first genome from each patient was considered to be day 0. The sampling periods and frequencies within the dataset was highly variable, 218 days was the longest time-period covered within the dataset but the majority were much shorter, the full details of the dataset are available in **supplementary table 1**. All mutations with cumulative frequencies ≥ 2 were labelled on-graph.

The only recurrent deletions observed in the dataset were located within the NTD of S-gene; S:Δ67 region (recurrent deletion region 1/RDR1), S:Δ138 region (RDR2), and S:Δ243 region (RDR4) (McCarthy et al., 2021). S:Δ138 region was the most frequent with four occurrences, followed by S:Δ67 region and S:Δ138 region with two occurrences respectively. Deletions within the S:67 region consisted of one S:Δ67 and one S:Δ69-70, the unconventional annotation is the result of the algorithm utilised to cluster deletions, the genome series in which S:Δ67 occurred already possessed S:Δ69 in its day 0 genome. S-gene constitutes just over one eighth of the overall SARS-CoV-2 genome by length; despite this ∼34% (79/234) of the total DNM occurrences were observed within S-gene as well as 59% (13/22) of the recurrent DNMs.

Non-spike, non-ORF1ab SARS-CoV-2 genes demonstrated a lower number of DNM occurrences (Figure 3, Figure 1). Three mutations within Matrix (M) and Envelope (E) were notable in their frequency (≥ 2 occurrences in the period); E:T30I, and M:H125Y. E:T30I was the only recurrent DNM observed within E-gene and the second most frequent DNM revealed by the analysis overall at 6 occurrences. E:T30I occurrences were not observed to be associated with any particular source study, geographical region, or SARS-CoV-2 lineage suggesting this may be a sensitive marker for persistent infection. Within M-gene M:H125Y was the only recurrent DNM with four occurrences.

**Figure 3:**
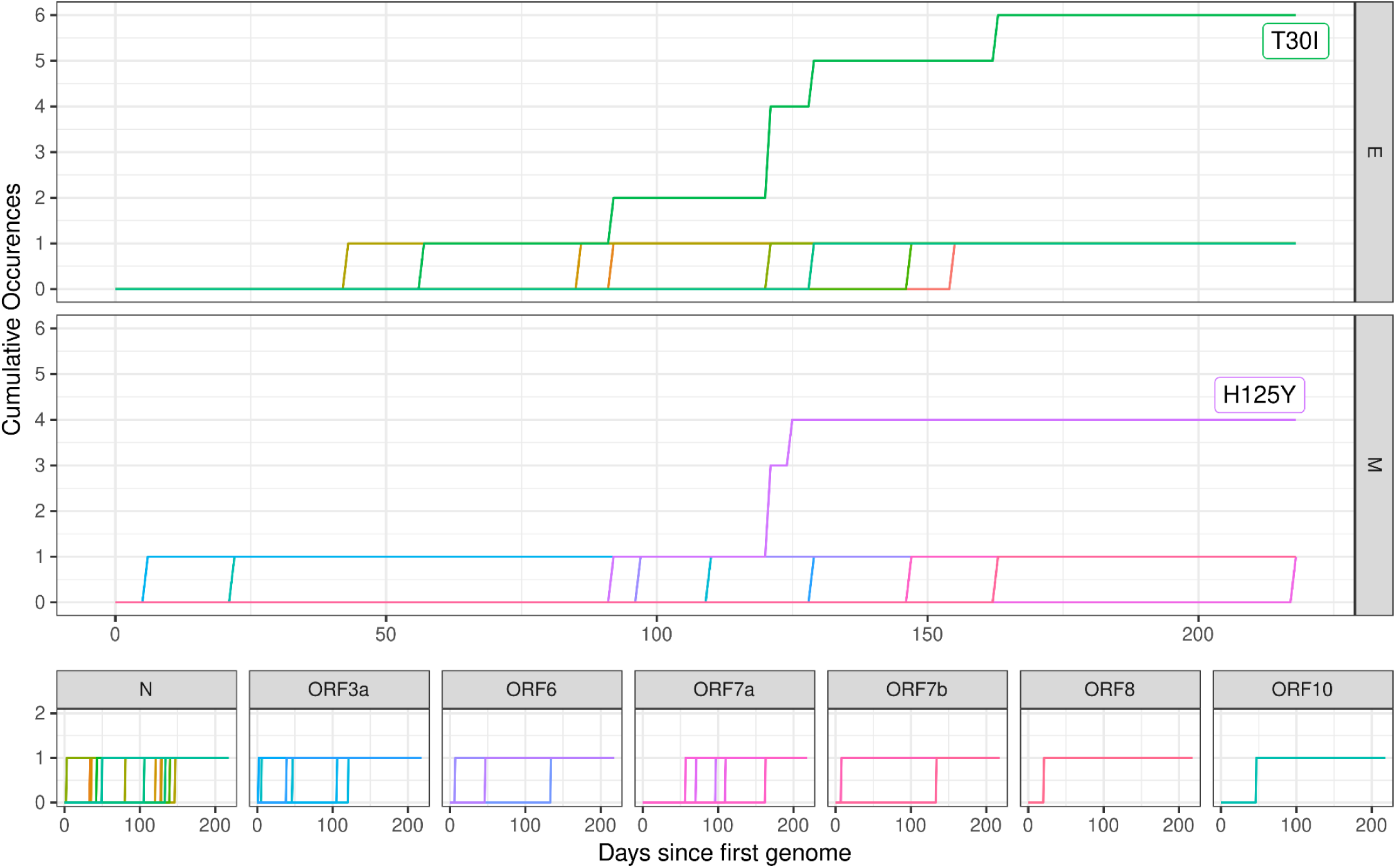
Cumulative occurrences of non-synonymous de-novo mutations in genes other than S or ORF1ab subdivided by gene in 168 genomes obtained from 28 patients. Mutations were observed in the following genes; **E** (encodes envelope protein), **M** (encodes membrane glycoprotein), **N** (encodes nucleocapsid phosphoprotein), **ORF10** (encodes ORF10 protein), **ORF3a** (encodes ORF3a protein), **ORF6** (encodes ORF6 protein), **ORF7a** (encodes ORF7a protein), and **ORF8** (encodes ORF8 protein) the full details of the gene definitions used are available from (Wu et al., 2020).The first genome from each patient was considered to be day 0. The sampling periods and frequencies within the dataset was highly variable, 218 days was the longest time-period covered within the dataset but the majority were much shorter, the full details of the dataset are available in **supplementary table 1**. All mutations with cumulative frequencies ≥ 2 were labelled on-graph.

When DNMs observed in these genes were clustered by AA loci the findings remained almost entirely unchanged other than in the case of the loci M:2 which was raised to three DNM occurrences by day 218 rather than the two presented in (Figure 3).

ORF1ab polyprotein genes, constituting the Non-structural proteins (NSPs) of SARS-CoV-2, demonstrated a larger number of recurrent mutations but still far fewer than in spike (Figure 4). Six DNMs were notable for their occurrence frequency; NSP3:T504P, NSP3:T820I, NSP3:P822L, NSP3:K977Q, NSP4:T295I, and NSP12:V792I. ORF1ab contained eighty-six out of the 195 DNMs observed, but only six of the total of twenty-one of the recurrent DNMs. ORF1ab constitutes more than two-thirds of the overall SARS-CoV-2 genome by length making the number of overall DNMs within the polyprotein disproportionately lower than would be expected if the distribution were random.

**Figure 4:**
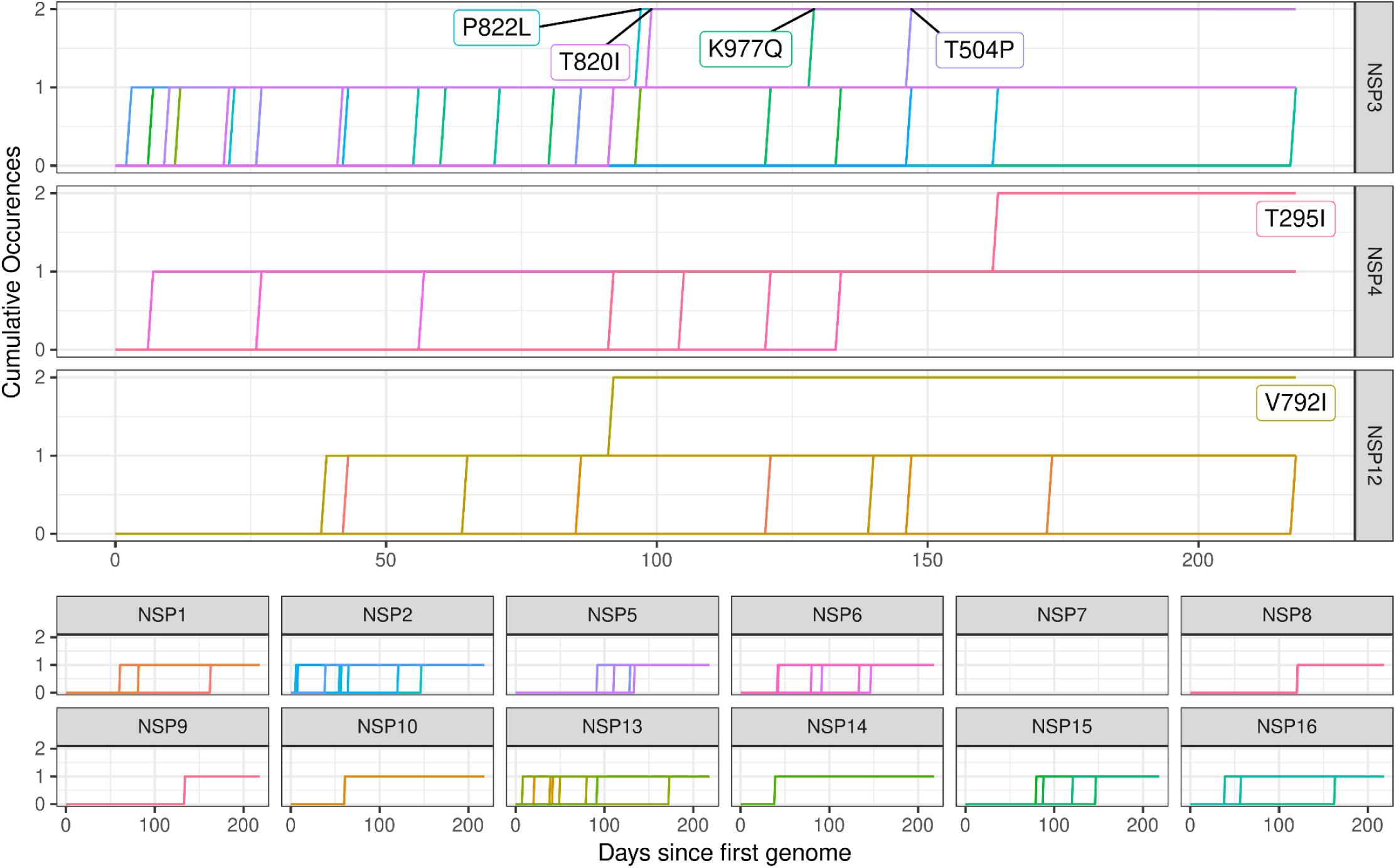
Cumulative occurrences of non-synonymous de-novo mutations in ORF1ab polyprotein subdivided by gene in 168 genomes obtained from 28 patients. **NSP12** mutations were annotated relative to the reading frame post ribosomal frameshift, no mutations were observed within **NSP12** prior to this loci (13,468). The first genome from each patient was considered to be day 0. The sampling periods and frequencies within the dataset was highly variable, 218 days was the longest time-period covered within the dataset but the majority were much shorter, the full details of the dataset are available in **supplementary table 1**. All mutations with cumulative frequencies ≥ 2 were labelled on-graph.

When DNMs observed within ORF1ab were clustered by AA loci the overall shape of the results remain broadly identical with two exceptions; NSP3:T504 and NSP3:P822 where their day 218 occurrences are raised to 3 and 4, respectively.

The relative frequencies for each recurrent mutation observed in the DNM occurrence analysis was compared to their prevalence within the COG-UK dataset (on the date 23/11/2021) (Table 1). As in the initial analysis S:E484K, E:T30I, and M:H125Y are noteworthy in their frequency especially compared to their low frequency in the larger COG-UK dataset.

**Table 1:** DNM Occurrence Frequencies for all Recurrent DNMs in this analysis and the COG-UK dataset (n=1,576,942). COG-UK dataset figures were generated using the dataset as it existed on 07/12/2021. Data was generated via CLIMB-Covid (Nicholls et al., 2021).

| DNM Annotation | Frequency in DNM Occurrence Analysis | Frequency in COG-UK Dataset | Percentage of genome series in which DNM occurred | Percentage of Genomes in COG-UK with DNM |
| --- | --- | --- | --- | --- |
| S:E484K | 8 | 3,437 | 28.57% | 0.2180% |
| E:T30I | 6 | 208 | 21.42% | 0.0132% |
| M:H125Y | 4 | 2,188 | 14.29% | 0.1387% |
| S:Δ138 region | 4 | 283,289 | 14.29% | 17.9645% |
| NSP4:T295I | 3 | 1,933 | 10.71% | 0.1226% |
| S:Q493K | 3 | 59 | 10.71% | 0.0037% |
| S:Δ67 region | 2 | 292,969 | 7.14% | 18.5783% |
| S:S13I | 2 | 211 | 7.14% | 0.0134% |
| NSP12:V792I | 2 | 10 | 7.14% | 0.0006% |
| NSP3:P822L | 2 | 28,410 | 7.14% | 1.8016% |
| NSP3:T820I | 2 | 442 | 7.14% | 0.0280% |
| NSP3:T504P | 2 | 18 | 7.14% | 0.0011% |
| S:L452R | 2 | 1,010,866 | 7.14% | 64.1029% |
| S:Q498R | 2 | 225 | 7.14% | 0.0143% |
| S:E484G | 2 | 46 | 7.14% | 0.0029% |
| S:Δ243 region | 2 | 546 | 7.14% | 0.0346% |
| S:F486I | 2 | 6 | 7.14% | 0.0004% |
| S:G142V | 2 | 1,361 | 7.14% | 0.0863% |
| S:T95I | 2 | 682,286 | 7.14% | 43.2664% |
| NSP3:K977Q | 2 | 391 | 7.14% | 0.0248% |
| S:F490L | 2 | 463 | 7.14% | 0.0294% |

Each observed recurrent DNM was compared to the UKHSA VOC/VUI definition files (Table 2). S:E484K was the most frequent DNM to appear in VOC/VUI definitions with eleven appearances, then S:L452R with four, then S:T95I and S:Δ138/RDR2 region with three each, followed by NSP3:K977Q, NSP3:P822L, S:Q498R, S:Δ67/RDR1 region, and S:Δ243/RDR4 region with one each. Of the twenty-one recurrent DNMs observed in the analysis nine of them are considered defining mutations for a VOC/VUI.

**Table 2:** Recurrent mutations which are variant defining based upon UKHSA variant definitions. Variant definitions were parsed from the UKHSA variant definition files available at: https://github.com/phe-genomics/variant_definitions. Lineages were called using pangolin (O’Toole et al., 2021b).

| Mutation Annotation | Pango Lineage | UKHSA Label | WHO Label |
| --- | --- | --- | --- |
| NSP3:K977Q | P.1 | VOC-21JAN-02 | Gamma |
| NSP3:P822L | AV.1 | VUI-21MAY-01 | n/a |
| S:E484K | B.1.351 | VOC-20DEC-02 | Beta |
| S:E484K | B.1.525 | VUI-21FEB-03 | Eta |
| S:E484K | P.1 | VOC-21JAN-02 | Gamma |
| S:E484K | A.23.1 | VUI-21FEB-01 | n/a |
| S:E484K | AV.1 | VUI-21MAY-01 | n/a |
| S:E484K | B.1.1.318 | VUI-21FEB-04 | n/a |
| S:E484K | B.1.1.7 (with E484K) | VOC-21FEB-02 | n/a |
| S:E484K | B.1.324.1 | VUI-21MAR-01 | n/a |
| S:E484K | P.3 | VUI-21MAR-02 | Theta |
| S:E484K | P.2 | VUI-21JAN-01 | Zeta |
| S:E484K | B.1.621 | VUI-21JUL-01 | n/a |
| S:L452R | B.1.617.2 | VOC-21APR-02 | Delta |
| S:L452R | B.1.617.1 | VUI-21APR-01 | Kappa |
| S:L452R | B.1.617.3 | VUI-21APR-03 | n/a |
| S:L452R | C.36.3 | VUI-21MAY-02 | n/a |
| S:Q498R | BA.1 | VOC-21NOV-01 | Omicron |
| S:T95I | AV.1 | VUI-21MAY-01 | n/a |
| S:T95I | B.1.1.318 | VUI-21FEB-04 | n/a |
| S:T95I | B.1.621 | VUI-21JUL-01 | n/a |
| S:Δ67 region/RDR1 | B.1.1.7 | VOC-20DEC-01 | Alpha |
| S:Δ138 region/RDR2 | B.1.1.7 | VOC-20DEC-01 | Alpha |
| S:Δ138 region/RDR2 | AV.1 | VUI-21MAY-01 | n/a |
| S:Δ138 region/RDR2 | B.1.1.318 | VUI-21FEB-04 | n/a |
| S:Δ243 region/RDR4 | C.37 | VUI-21JUN-01 | Lambda |

## Discussion

Not all mutations are discussed in detail, while a literature search has been performed for every recurrent DNM only those with sufficient literature available for discussion to be informative were included below.

### S-Gene - Receptor Binding Domain Recurrent Mutations

The frequency of RBD DNMs observed in this analysis is a significant finding; the RBD is a relatively small region of the SARS-CoV-2 genome making up less than two percent of the genome by length, but these account for 17 percent of all DNMs observed (Figure 1). It is clear that RBD mutations were the most strongly selected for in the immunocompromised patients included within the dataset.

The sharp rise of S:E484K occurrences early in the period is biased due to the data from (Jensen et al., 2021) as a result of their sampling strategy and research focus. (Jensen et al., 2021) specifically discussed the emergence of S:E484K in long-term immunocompromised patients and published short periods of surveillance of these cases when the patients in question had significantly longer shedding periods to demonstrate this. However, even if this study is excluded S:E484K remains the most frequently occurring DNM within spike.

The high frequency of the S:E484K occurrences is suggestive of a strong selective pressure; this is further demonstrated by the total of twelve DNMs observed at the S:484 loci. The two occurrences of S:E484G in the dataset also suggest that the glycine substitution is subject to differing selection pressures than the lysine substitution in S:E484K although this may be host dependent. In one of the two occurrences of S:E484G this change was transient and was replaced by S:E484K. There are two possible explanations for this observation; a secondary mutation, or both mutations occurred within the patient and the S:E484K subpopulation outcompeted the S:E484G population to become dominant. There is no single nucleotide change by which a G -> K AA change might occur, supporting the second possibility. If the second explanation is correct it would suggest that S:484 mutations are selected for generally. The large difference between the frequency of S:E484K in this dataset compared to the national COG-UK dataset further suggests that the selection pressures which caused S:E484K to be so frequent within this analysis are not true of the majority of hosts (Table 1). S:E484K is also considered a defining mutation for a large number of variants, further indicating a strong selection pressure for the mutation (Table 2). Despite its presence within a large number of variants it is only present within a small proportion of the COG-UK dataset suggesting that on a population level it may have a deleterious effect on transmission, however this may be explained by other factors such as variants with S:E484K not being common in the UK generally.

A strong selective pressure for S:E484K was also observed by Zahradník et al, (2021) who discovered using an *in vitro* experimental evolution model, that >70% of clones in one library gained S:E484K and S:N501Y which were associated with a significant increase in ACE2 affinity. Furthermore they observed the occurrence of the mutation S:Q498R alongside S:N501Y in two repeats, this combination was observed to lead to significantly greater affinity to ACE2 compared to both wild-type and Alpha which rose further alongside S:E484K. This combination was only observed within a single patient (pt 19) although the combination E484G, Q498R and N501Y did arise in a further patient (pt 17), in both cases the infections were Alpha and therefore already possessed S:N501Y. At the time Zahradník et al was published this constellation of mutations had not been observed in wild virus but with the emergence of Omicron this combination has become significantly more frequent (albeit with E484A rather than E484K).

The low occurrence frequency of S:N501Y compared to that observed by (Zahradník et al., 2021) is also notable but is partly explained by its high (nine out of twenty-eight) day zero frequency in the genome series’, due to the high amount of long-term Alpha infections included in this study. When DNMs were clustered by AA loci S:501 was highlighted as recurrent however.

Another notable observation is the two *de novo* occurrences of S:L452R (a defining mutation of Delta, Kappa and Epsilon variants) which aids both immune evasion and ACE2 affinity (Motozono et al., 2021).

S:Q493K has previously been identified by Huang et al, (2021) as a highly beneficial adaptation to a mouse host, improving spike binding affinity to murine ACE2 (Huang et al., 2021), its rarity in the overall SARS-CoV-2 population (58 in COG-UK dataset) suggests that it is not strongly selected for in a human host generally. The three occurrences in this dataset may suggest that S:Q493K does confer a benefit to the virus within the context of a long-term infection but not in transient infection. A highly similar mutation, S:Q493R, is a defining mutation of the Omicron variant.

S:F486I has been observed to decrease the affinity of some neutralising antibodies to spike protein (Xu et al., 2021), and may decrease the affinity of spike to ACE2 (Clark et al., 2021), S:F486I has furthermore been associated with mink adaptation (Zhou et al., 2021). S:490L has been observed to reduce the affinity of multiple mAbs as well as decreasing the neutralisation sensitivity of pseudovirus to convalescent sera, however it does not appear to have an impact on viral infectivity (Li et al., 2020). It is noteworthy that a large number of mutations described in this present study are associated with enhanced human ACE2 affinity including Q493K, Q498R and N501Y (Starr et al., 2020)

When AA loci clustering was performed recurrent DNMs at S:330 and S:440 were observed.

Finally, although most of this study has considered mutations in isolation, several of the late stage long-term infections showed interesting combinations of mutations, particularly within Spike (figure 5). Patient 19 for example was an Alpha infection that had picked up a large number of mutations, many of which were in common with, or similar to Omicron, for example A67D, G142V, T95I, Δ210/L212I, E484K, and Q498R. A further case, patient 17 also contained E484G and Q498R alongside the Alpha lineage defining mutation, N501Y and pt 27 contained T95I, a further deletion at S:Δ138 region and G496S, in common with Omicron.

**Figure 5:**
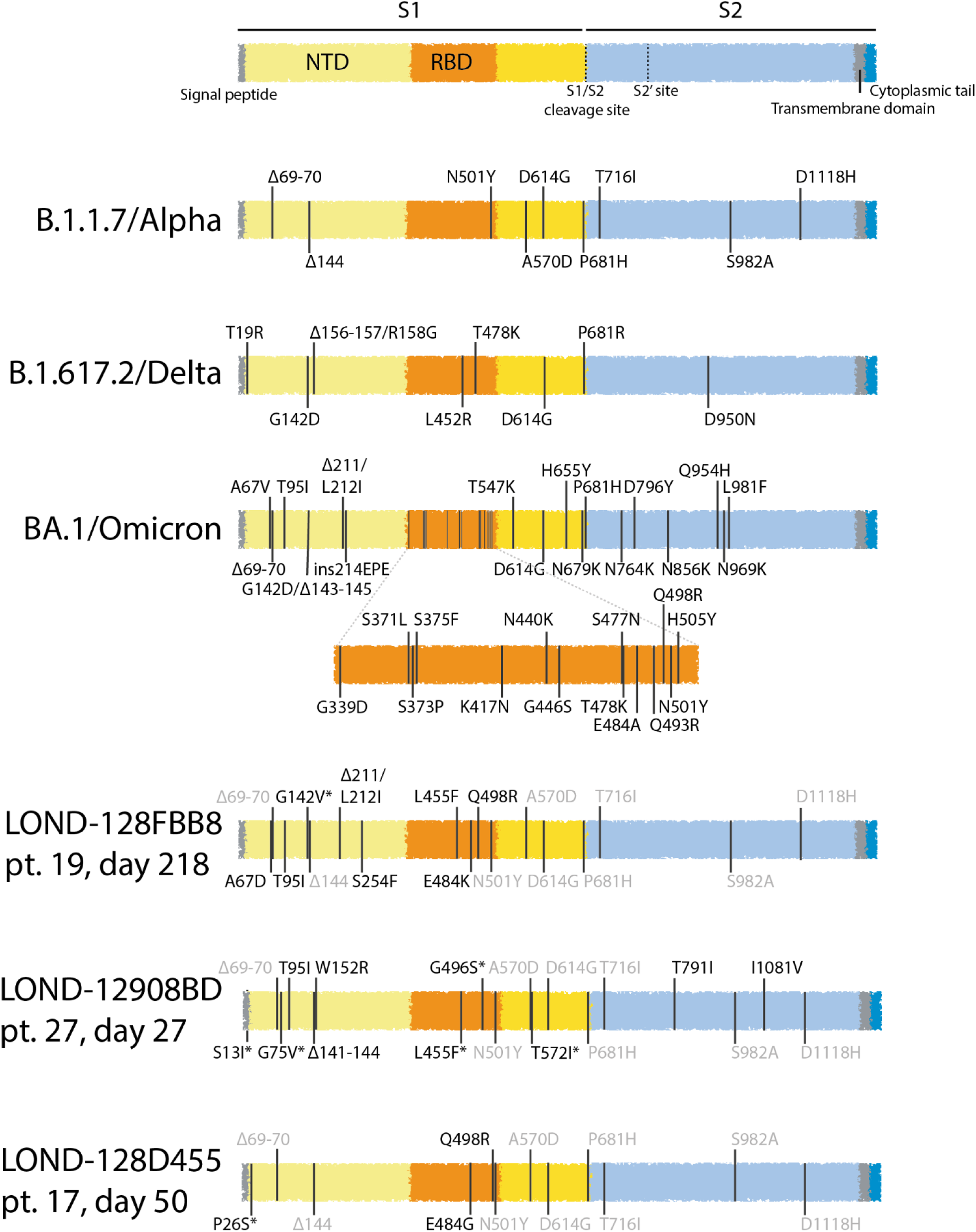
Spike mutational profiles of particular interest described by this study. Select spikes from late sequencing of 3 long-term Alpha infections shown as Spike schematics. Spike variants from WT Alpha, Delta and BA.1 Omicron shown for comparison. Mutations shown in grey are existing lineage-defining Alpha mutations. Mutations marked with an asterisk indicate mixed, but resolvable bases in the sequence.

### S-Gene N-terminal Domain Recurrent Mutations

T95I has been shown to bind of the human Tyrosine-protein kinase receptor UFO (AXL) and it has been suggested by (Singh et al., 2021) that AXL facilitates SARS-CoV-2 cell entry to the same extent as ACE2 in AXL overexpressed cell culture. NTD also has a substantial role in the antigenicity of spike with multiple escape mutations identified in this domain (Harvey et al., 2021).

All recurrent deletions within the SARS-CoV-2 genome were observed within the NTD (S:Δ67 region/RDR1, S:Δ138/RDR2 region, and S:Δ243/RDR4 region). Deletions within the S:69-70 region are commonly observed (McCarthy et al., 2021; Meng et al., 2021). Meng et al, (2021) characterised the common S:Δ69-70 deletion as contributing to infectivity by improving incorporation of cleaved spike protein into virions and possibly has a compensatory effect on mutations in the RBD associated with Ab escape such as N439K and Y453F. Of the two observations of deletions within the S:67-70 region one was S:Δ69-70 whereas the other was S:Δ67 which has not been commonly observed, but it is notable that the genome series in which S:Δ67 was observed already possessed S:Δ69 at day 0. S:Δ69-70 is also a defining mutation of the Alpha and Omicron variants and is responsible for the S-gene target failure observed in the PCR testing of alpha variant samples with TaqPath SARS-CoV-2 PCR kits (Kidd et al., 2021).

*De novo* occurrences of slightly differing deletions within the S:Δ138/RDR2 region were observed 4 times. This region makes up part of the “NTD antigenic supersite” which the majority of neutralising antibodies against the NTD target (McCallum et al., 2021b). S:Δ140 has consequently been associated with a significant decrease in Ab neutralisation (Andreano et al., 2021; Liu et al., 2021). Based upon the high number of occurrences, it appears likely that deletions in this region confer some benefit to the virus during long-term infections. As with S:N501Y, as well as S:Δ67, it is worth noting a substantial proportion of long-term infections already carried deletions in the S:Δ138 region at day 0 due to being the Alpha variant.

Two occurrences of S:Δ243, another NTD supersite mutation, were also observed, another deletion which has been demonstrated to decrease Ab neutralisation *in vitro* (McCallum et al., 2021b; McCarthy et al., 2021).

### S-Gene Signal Peptide Recurrent Mutations

The single recurrent signal peptide DMN, S:S13I, has been previously shown to mediate a shift of the cleavage site of the signal peptide which in turn facilitates immune evasion by causing a significant re-arrangement of the NTD antigenic supersite and its constituent internal disulphide bonding (McCallum et al., 2021a, 2021b).

### E-Gene Recurrent Mutations

The most frequent *de novo* mutation observed outside of the spike gene is Envelope:T30I (the second most frequent mutation overall after S:E484X). This mutation was observed by Chaudhry et al, (2020) in a cell-culture passage experiment, where it conferred a growth advantage in Calu-3 cells but slowed growth in Vero E6 cells (Chaudhry et al., 2020).

The high frequency of E:T30I is strongly suggestive of a selective pressure during long-term infections and further suggests that the conditions experienced by the virus in immunocompromised patients may exist in a similar selective environment as cell culture, potentially due to a lack of stability needed for transmission. The significant enrichment of E:T30I in this analysis compared to the COG-UK dataset (Table 2) suggests that E:T30I may be a deleterious mutation within the circulating SARS-CoV-2 population. A single variant lineage, B.1.616, does contain E:T30I as a lineage-defining mutation. Interestingly, B.1.616 was associated with an extremely localised, largely nosocomial-associated outbreak, suggesting the possibility this may have been the emergence of a virus from a long-term infection (Fillâtre et al., 2021). This also raises the hypothetical possibility that E:T30I may be considered a marker of long-term SARS-CoV-2 infections. Further study is necessary to determine the phenotypic effect of this mutation and its role in influencing within and between host fitness.

### ORF1ab-NSP3 Recurrent Mutations

Literature concerning mutations in ORF1ab is generally observational rather than experimental due to the current lack of tractable models to study them *in vitro*. The concentration of higher frequency mutations within the NSP3 gene is not surprising considering it is the largest gene within the ORF1ab polyprotein and is known to be a bulky, modular protein which may have some flexible linker regions which are fairly hypermutatable. Stanevich et al identified NSP3:T504P as a mutation associated with cytotoxic T cell epitope immune escape (Stanevich et al., 2021).

## Conclusions

This work sought to determine recurrent mutations across the SARS-CoV-2 genome associated with long-term infections in immunodeficient patients. This study has several notable limitations: importantly a significant publication bias is likely to be present which may overemphasis the importance of some mutations. S:E484K especially is affected by this, the six genome series obtained from Jensen et al., (2021) were published to demonstrate the emergence of S:E484K within immunocompromised patients. Further work will attempt to avoid this by utilising less biased sampling strategies from long-term infected patients, requiring a prospective study design that aims to regularly sample genomes from long-term infected patients.

The majority of recurrently observed DNMs have been associated with immune escape, increased ACE2 affinity, or improved viral packaging and are generally not highly prevalent within the wider SARS-CoV-2 population (with the exception of some SARS-CoV-2 variants).

These factors suggest that the conditions during long-term infections at least partly select for mutations which aid the virus with *intra*-host replication and persistence as opposed to the general SARS-CoV-2 population, where mutations which aid *inter*-host transmission are more strongly selected for. E:T30I in particular is worthy of further study as a potential marker of long-term SARS-CoV-2 infections.

However, the large number of occurrences overlapping with variant defining mutations observed does indicate that patients within this category should not be discounted as a potential source of previous, or indeed future variants. The potential of mutations which aid cell-cell transmission within the host or improve viral packaging may affect virulence and any mutations within this category which do not impact viral transmissibility could have a significant impact. This is highly relevant as many of the most abundant mutations described in this dataset are found across many variant lineages. Furthermore, its possible sub-neutralising levels of antibodies which may be present in some cases (either homologous or from heterologous convalescent or monoclonal antibody treatments) could be selecting for the acquisition of antigenic mutations seen in several of these cases (Kemp et al., 2021).

At present it is unresolved where SARS-CoV-2 variants emerge from. One prevailing hypothesis is that some variants emerged from long-term chronic infections, generating novel advantageous combinations of mutations without the stringent selection pressure of transmission, eventually resulting in an outbreak and onward transmission. We have compared common mutations arising during chronic infections and described how many are shared with SARS-CoV-2 variant lineages. Furthermore we present evidence, based on a rare mutational signature, that the French B.1.616 variant lineage arose from a direct and recent spillover from a chronic infection. Overall the data presented here is consistent and supportive of the chronic infection hypothesis of SARS-CoV-2 variant emergence. Therefore we suggest identifying and curing chronic infections, preferably with combined antiviral therapy as would be used for more traditionally chronic viruses (HIV, HCV), both to the infected individual, but also to global health. Intrahost variation of SARS-CoV-2 is likely to play a significant role within this patient group however the lack of raw data availability for the majority of the samples within this dataset makes this challenging (Chaudhry et al., 2020).

We anticipate this dataset will be maintained as a public resource to enable the study of long-term SARS-CoV-2 infections in immunodeficient patients for as long as it is deemed relevant to enable other researchers to contribute to this understudied, highly important, patient group (https://github.com/BioWilko/recurrent-sars-cov-2-mutations/blob/main/dataset/mutation_calls.csv).

## Data Availability

All data included in the present work are available online or upon reasonable request to the producing author. The full details of the data are available within the supplement.

https://github.com/BioWilko/recurrent-sars-cov-2-mutations

## Acknowledgments

*COG-UK is supported by funding from the Medical Research Council (MRC) part of UK Research & Innovation (UKRI), the National Institute of Health Research (NIHR) [grant code: MC_PC_19027], and Genome Research Limited, operating as the Wellcome Sanger Institute*.

*The COG-UK study protocol was approved by the Public Health England Research Ethics Governance Group (reference: R&D NR0195). Authors only had access to anonymised data. No individual patient consent was required*.

**Queen Elizabeth Hospital, University Hospitals Birmingham, Birmingham B15 2TH, UK**.

- Mark Garvey, Anna Casey, Liz Ratcliffe, Husam Osman
- Contact

**Choi, B**., **Choudhary, M.C**., **Regan, J**., **Sparks, J.A**., **Padera, R.F**., **et al**., **2020. Persistence and Evolution of SARS-CoV-2 in an Immunocompromised Host. N. Engl. J. Med. 383, 2291–2293. https://doi.org/10.1056/NEJMc2031364**

- Bina Choi, M.D., Manish C. Choudhary, Ph.D., James Regan, B.S., Jeffrey A. Sparks, M.D., Robert F. Padera, M.D., Ph.D. : **Brigham and Women’s Hospital, Boston, MA**. Xueting Qiu, Ph.D. : **Harvard T.H. Chan School of Public Health, Boston, MA**. Isaac H. Solomon, M.D., Ph.D. : **Brigham and Women’s Hospital, Boston, MA**. Hsiao-Hsuan Kuo, Ph.D., Julie Boucau, Ph.D., Kathryn Bowman, M.D., U. Das Adhikari, Ph.D., : **Ragon Institute of MGH, MIT, and Harvard, Cambridge, MA**. Marisa L. Winkler, M.D., Ph.D., Alisa A. Mueller, M.D., Ph.D., Tiffany Y.-T. Hsu, M.D., Ph.D., Michaël Desjardins, M.D., Lindsey R. Baden, M.D., Brian T. Chan, M.D., M.P.H. : **Brigham and Women’s Hospital, Boston, MA**. Bruce D. Walker, M.D. : **Ragon Institute of MGH, MIT, and Harvard, Cambridge, MA**. Mathias Lichterfeld, M.D., Ph.D., Manfred Brigl, M.D. : **Brigham and Women’s Hospital, Boston, MA**. Douglas S. Kwon, M.D., Ph.D. : **Ragon Institute of MGH, MIT, and Harvard, Cambridge, MA**. Sanjat Kanjilal, M.D., M.P.H.cool : **Brigham and Women’s Hospital, Boston, MA**. Eugene T. Richardson, M.D., Ph.D. : **Harvard Medical School, Boston, MA**. A. Helena Jonsson, M.D., Ph.D. : **Brigham and Women’s Hospital, Boston, MA**. Galit Alter, Ph.D., Amy K. Barczak, M.D. : **Ragon Institute of MGH, MIT and Harvard, Cambridge, MA**. William P. Hanage, Ph.D. : **Harvard T.H. Chan School of Public Health, Boston, MA**. Xu G. Yu, M.D., Gaurav D. Gaiha, M.D., D.Phil., : **Ragon Institute of MGH, MIT and Harvard, Cambridge, MA**. Michael S. Seaman, Ph.D. : **Beth Israel Deaconess Medical Center, Boston, MA**. Manuela Cernadas, M.D., Jonathan Z. Li, M.D. : **Brigham and Women’s Hospital, Boston, MA**.
- **Contact:** Manuela Cernadas

**Avanzato, V.A**., **Matson, M.J**., **Seifert, S.N**., **Pryce, R**., **Williamson, B.N**., **et al**., **2020. Case Study: Prolonged Infectious SARS-CoV-2 Shedding from an Asymptomatic Immunocompromised Individual with Cancer. Cell 183, 1901-1912.e9. https://doi.org/10.1016/j.cell.2020.10.049**

- Victoria A.Avanzato, Jeremiah Matson, Stephanie N.Seifert, Rhys Pryce, Brandi N.Williamson, Sarah L.Anzick, Kent Barbian, Seth D.Judson, Elizabeth R.Fischer, Craig Martens, Thomas A.Bowden, Emmiede Wit, Francis X.Riedo, Vincent J.Munster.
- **Contact:**

**Reuken, P.A**., **Stallmach, A**., **Pletz, M.W**., **Brandt, C**., **Andreas, N**., **et al**., **2021. Severe clinical relapse in an immunocompromised host with persistent SARS-CoV-2 infection. Leukemia 35, 920–923. https://doi.org/10.1038/s41375-021-01175-8**

- Philipp A. Reuken, Andreas Stallmach, Mathias W. Pletz, Christian Brandt, Nico Andreas, Sabine Hahnfeld, Bettina Löffler, Sabine Baumgart, Thomas Kamradt & Michael Bauer
- **Contact:**

**Tarhini, H**., **Recoing, A**., **Bridier-nahmias, A**., **Rahi, M**., **Lambert, C**., **et al**., **2021. Long-Term Severe Acute Respiratory Syndrome Coronavirus 2 (SARS-CoV-2) Infectiousness Among Three Immunocompromised Patients: From Prolonged Viral Shedding to SARS-CoV-2 Superinfection. J. Infect. Dis. 223, 1522–1527. https://doi.org/10.1093/infdis/jiab075**

- Hassan Tarhini, Amélie Recoing, Antoine Bridier-nahmias, Mayda Rahi, Céleste Lambert, Pascale Martres, Jean-Christophe Lucet, Christophe Rioux, Donia Bouzid, Samuel Lebourgeois, Diane Descamps, Yazdan Yazdanpanah, Quentin Le Hingrat, François-Xavier Lescure, Benoit Visseaux
- **Contact:**

**Kemp, S.A**., **Collier, D.A**., **Datir, R.P**., **Ferreira, I.A.T.M**., **Gayed, S**., **et al**., **2021. SARS-CoV-2 evolution during treatment of chronic infection. Nature 592, 277–282. https://doi.org/10.1038/s41586-021-03291-y**

- Steven A. Kemp, Dami A. Collier, Rawlings P. Datir, Isabella A. T. M. Ferreira, Salma Gayed, Aminu Jahun, Myra Hosmillo, Chloe Rees-Spear, Petra Mlcochova, Ines Ushiro Lumb, David J. Roberts, Anita Chandra, Nigel Temperton, The CITIID-NIHR BioResource COVID-19 Collaboration, The COVID-19 Genomics UK (COG-UK) Consortium, Katherine Sharrocks, Elizabeth Blane, Yorgo Modis, Kendra E. Leigh, John A. G. Briggs, Marit J. van Gils, Kenneth G. C. Smith, John R. Bradley, Chris Smith, Rainer Doffinger, Lourdes Ceron-Gutierrez, Gabriela Barcenas-Morales, David D. Pollock, Richard A. Goldstein, Anna Smielewska, Jordan P. Skittrall, Theodore Gouliouris, Ian G. Goodfellow, Effrossyni Gkrania-Klotsas, Christopher J. R. Illingworth, Laura E. McCoy & Ravindra K. Gupta.
- **Contact:**

**Baang, J.H**., **Smith, C**., **Mirabelli, C**., **Valesano, A.L**., **Manthei, D.M**., **et al**., **2021. Prolonged Severe Acute Respiratory Syndrome Coronavirus 2 Replication in an Immunocompromised Patient. J. Infect. Dis. 223, 23–27. https://doi.org/10.1093/infdis/jiaa666**

- Ji Hoon Baang, Christopher Smith, Carmen Mirabelli, Andrew L Valesano, David M Manthei, Michael A Bachman, Christiane E Wobus, Michael Adams, Laraine Washer, Emily T Martin, Adam S Lauring.
- **Contact:**

**Stanevich, O**., **Alekseeva, E**., **Sergeeva, M**., **Fadeev, A**., **Komissarova, K**., **et al**., **2021. SARS-CoV-2 escape from cytotoxic T cells during long-term COVID-19 (preprint). In Review. https://doi.org/10.21203/rs.3.rs-750741/v1**

- Oksana Stanevich, Evgeniia Alekseeva, Maria Sergeeva, Artem Fadeev, Kseniya Komissarova, Anna Ivanova, Tamara Simakova, Kirill Vasilyev, Anna-Polina Shurygina, Marina Stukova, Ksenia Safina, Elena Nabieva, Sofya Garushyants, Galya Klink, Evgeny Bakin, Jullia Zabutova, Anastasia Kholodnaia, Olga Lukina, Irina Skorokhod, Viktoria Ryabchikova, Nadezhda Medvedeva, Dmitry Lioznov, Daria Danilenko, Dmitriy Chudakov, Andrey Komissarov, Georgii Bazykin.
- **Contact:**

**Khatamzas, E**., **Rehn, A**., **Muenchhoff, M**., **Hellmuth, J**., **Gaitzsch, E**., **et al**., **2021. Emergence of multiple SARS-CoV-2 mutations in an immunocompromised host. https://doi.org/10.1101/2021.01.10.20248871**

- Elham Khatamzas, Alexandra Rehn, Maximilian Muenchhoff, Johannes Hellmuth, Erik Gaitzsch, Tobias Weiglein, Enrico Georgi, Clemens Scherer, Stephanie Stecher, Oliver Weigert, Philipp Girl, Sabine Zange, Oliver T. Keppler, Joachim Stemmler, Michael von Bergwelt-Baildon, Roman Wölfel, Markus Antwerpen.
- **Contact:**

**Borges, V**., **Isidro, J**., **Cunha, M**., **Cochicho, D**., **Martins, L**., **et al**., **2021. Long-Term Evolution of SARS-CoV-2 in an Immunocompromised Patient with Non-Hodgkin Lymphoma. mSphere 6, e0024421. https://doi.org/10.1128/mSphere.00244-21**

- Vítor Borges, Joana Isidro, Mário Cunha, Daniela Cochicho, Luis Martins, Luis Banha, Margarida Figueiredo, Leonor Rebelo, Maria Céu Trindade, Sílvia Duarte, Luís Vieira, Maria João Alves, Inês Costa, Raquel Guiomar, Madalena Santos, Rita Cortê-Real, André Dias, Diana Póvoas, João Cabo, Carlos Figueiredo, Maria José Manata, Fernando Maltez, Maria Gomes da Silva, João Paulo Gomes.
- **Contact:**

**Virology Department, NHS East and South East London Pathology Partnership, Royal London Hospital, Barts Health NHS Trust:**

- Beatrix Kele, Kathryn Harris, Theresa Cutino-Moguel, Dola Owoyemi, Shahiba Sultanam, Abril Romero.
- **Contact:**

**Ciuffreda, L**., **Lorenzo-Salazar, J.M**., **Alcoba-Florez, J**., **Rodriguez-Pérez, H**., **Gil-Campesino, H**., **et al**., **2021. Longitudinal study of a SARS-CoV-2 infection in an immunocompromised patient with X-linked agammaglobulinemia. J. Infect. 0. https://doi.org/10.1016/j.jinf.2021.07.028**

- Laura Ciuffreda, José M. Lorenzo-Salazar, Julia Alcoba-Florez, Héctor Rodriguez-Pérez, Helena Gil-Campesino, Antonio íñigo-Campos, Diego García-Martínez de Artola, Agustín Valenzuela-Fernández, Marcelino Hayek-Peraza, Susana Rojo-Alba, Marta Elena Alvarez-Argüelles, Oscar Díez-Gil, Rafaela González-Montelongo, Carlos Flores.
- **Contact:**

**Jensen, B**., **Luebke, N**., **Feldt, T**., **Keitel, V**., **Brandenburger, T**., **et al**., **2021. Emergence of the E484K mutation in SARS-COV-2-infected immunocompromised patients treated with bamlanivimab in Germany. Lancet Reg. Health – Eur. 8. https://doi.org/10.1016/j.lanepe.2021.100164**

- Bjoern Jensen, Nadine Luebke, Torsten Feldt, Verena Keite, Timo Brandenburge, Detlef Kindgen-Mille, Matthias Lutterbec, Noemi F Freis, David Schoele, Rainer Haa, Alexander Dilthe, Ortwin Adam, Andreas Walker, Joerg Timm, Tom Luedde.
- **Contact:**

**Weigang, S**., **Fuchs, J**., **Zimmer, G**., **Schnepf, D**., **Kern, L**., **et al**., **2021. Within-host evolution of SARS-CoV-2 in an immunosuppressed COVID-19 patient as a source of immune escape variants. Nat. Commun. 12, 6405. https://doi.org/10.1038/s41467-021-26602-3**

- Sebastian Weigang, Jonas Fuchs, Gert Zimmer, Daniel Schnepf, Lisa Kern, Julius Beer, Hendrik Luxenburger, Jakob Ankerhold, Valeria Falcone, Janine Kemming, Maike Hofmann, Robert Thimme, Christoph Neumann-Haefelin, Svenja Ulferts, Robert Grosse, Daniel Hornuss, Yakup Tanriver, Siegbert Rieg, Dirk Wagner, Daniela Huzly, Martin Schwemmle, Marcus Panning, Georg Kochs.
- **Contact:** **&**

## The COVID-19 Genomics UK (COG-UK) consortium

**June 2021 V.1**

**Funding acquisition, Leadership and supervision, Metadata curation, Project administration, Samples and logistics, Sequencing and analysis, Software and analysis tools, and Visualisation:**

Samuel C Robson ^13, 84^

**Funding acquisition, Leadership and supervision, Metadata curation, Project administration, Samples and logistics, Sequencing and analysis, and Software and analysis tools:**

Thomas R Connor ^11, 74^ and Nicholas J Loman ^43^

**Leadership and supervision, Metadata curation, Project administration, Samples and logistics, Sequencing and analysis, Software and analysis tools, and Visualisation:**

Tanya Golubchik ^5^

**Funding acquisition, Leadership and supervision, Metadata curation, Samples and logistics, Sequencing and analysis, and Visualisation:**

Rocio T Martinez Nunez ^46^

**Funding acquisition, Leadership and supervision, Project administration, Samples and logistics, Sequencing and analysis, and Software and analysis tools:**

David Bonsall ^5^

**Funding acquisition, Leadership and supervision, Project administration, Sequencing and analysis, Software and analysis tools, and Visualisation:**

Andrew Rambaut ^104^

**Funding acquisition, Metadata curation, Project administration, Samples and logistics, Sequencing and analysis, and Software and analysis tools:**

Luke B Snell ^12^

**Leadership and supervision, Metadata curation, Project administration, Samples and logistics, Software and analysis tools, and Visualisation:**

Rich Livett ^116^

**Funding acquisition, Leadership and supervision, Metadata curation, Project administration, and Samples and logistics:**

Catherine Ludden ^20, 70^

**Funding acquisition, Leadership and supervision, Metadata curation, Samples and logistics, and Sequencing and analysis:**

Sally Corden ^74^ and Eleni Nastouli ^96, 95, 30^

**Funding acquisition, Leadership and supervision, Metadata curation, Sequencing and analysis, and Software and analysis tools:**

Gaia Nebbia ^12^

**Funding acquisition, Leadership and supervision, Project administration, Samples and logistics, and Sequencing and analysis:**

Ian Johnston ^116^

**Leadership and supervision, Metadata curation, Project administration, Samples and logistics, and Sequencing and analysis:**

Katrina Lythgoe ^5^, M. Estee Torok ^19, 20^ and Ian G Goodfellow ^24^

**Leadership and supervision, Metadata curation, Project administration, Samples and logistics, and Visualisation:**

Jacqui A Prieto ^97, 82^ and Kordo Saeed ^97, 83^

**Leadership and supervision, Metadata curation, Project administration, Sequencing and analysis, and Software and analysis tools:**

David K Jackson ^116^

**Leadership and supervision, Metadata curation, Samples and logistics, Sequencing and analysis, and Visualisation:**

Catherine Houlihan ^96, 94^

**Leadership and supervision, Metadata curation, Sequencing and analysis, Software and analysis tools, and Visualisation:**

Dan Frampton ^94, 95^

**Metadata curation, Project administration, Samples and logistics, Sequencing and analysis, and Software and analysis tools:**

William L Hamilton ^19^ and Adam A Witney ^41^

**Funding acquisition, Samples and logistics, Sequencing and analysis, and Visualisation:**

Giselda Bucca ^101^

**Funding acquisition, Leadership and supervision, Metadata curation, and Project administration:**

Cassie F Pope ^40, 41^

**Funding acquisition, Leadership and supervision, Metadata curation, and Samples and logistics:**

Catherine Moore ^74^

**Funding acquisition, Leadership and supervision, Metadata curation, and Sequencing and analysis:**

Emma C Thomson ^53^

**Funding acquisition, Leadership and supervision, Project administration, and Samples and logistics:**

Ewan M Harrison ^116, 102^

**Funding acquisition, Leadership and supervision, Sequencing and analysis, and Visualisation:**

Colin P Smith ^101^

**Leadership and supervision, Metadata curation, Project administration, and Sequencing and analysis:**

Fiona Rogan ^77^

**Leadership and supervision, Metadata curation, Project administration, and Samples and logistics:**

Shaun M Beckwith ^6^, Abigail Murray ^6^, Dawn Singleton ^6^, Kirstine Eastick ^37^, Liz A Sheridan ^98^, Paul Randell ^99^, Leigh M Jackson ^105^, Cristina V Ariani ^116^ and Sónia Gonçalves ^116^

**Leadership and supervision, Metadata curation, Samples and logistics, and Sequencing and analysis:**

Derek J Fairley ^3, 77^, Matthew W Loose ^18^ and Joanne Watkins ^74^

**Leadership and supervision, Metadata curation, Samples and logistics, and Visualisation:**

Samuel Moses ^25, 106^

**Leadership and supervision, Metadata curation, Sequencing and analysis, and Software and analysis tools:**

Sam Nicholls ^43^, Matthew Bull ^74^ and Roberto Amato ^116^

**Leadership and supervision, Project administration, Samples and logistics, and Sequencing and analysis:**

Darren L Smith ^36, 65, 66^

**Leadership and supervision, Sequencing and analysis, Software and analysis tools, and Visualisation:**

David M Aanensen ^14, 116^ and Jeffrey C Barrett ^116^

**Metadata curation, Project administration, Samples and logistics, and Sequencing and analysis:**

Dinesh Aggarwal ^20, 116, 70^, James G Shepherd ^53^, Martin D Curran ^71^ and Surendra Parmar ^71^

**Metadata curation, Project administration, Sequencing and analysis, and Software and analysis tools:**

Matthew D Parker ^109^

**Metadata curation, Samples and logistics, Sequencing and analysis, and Software and analysis tools:**

Catryn Williams ^74^

**Metadata curation, Samples and logistics, Sequencing and analysis, and Visualisation:**

Sharon Glaysher ^68^

**Metadata curation, Sequencing and analysis, Software and analysis tools, and Visualisation:**

Anthony P Underwood ^14, 116^, Matthew Bashton ^36, 65^, Nicole Pacchiarini ^74^, Katie F Loveson ^84^ and Matthew Byott ^95, 96^

**Project administration, Sequencing and analysis, Software and analysis tools, and Visualisation:**

Alessandro M Carabelli ^20^

**Funding acquisition, Leadership and supervision, and Metadata curation:**

Kate E Templeton ^56, 104^

**Funding acquisition, Leadership and supervision, and Project administration:**

Thushan I de Silva ^109^, Dennis Wang ^109^, Cordelia F Langford ^116^ and John Sillitoe ^116^

**Funding acquisition, Leadership and supervision, and Samples and logistics:**

Rory N Gunson ^55^

**Funding acquisition, Leadership and supervision, and Sequencing and analysis:**

Simon Cottrell ^74^, Justin O’Grady ^75, 103^ and Dominic Kwiatkowski ^116, 108^

**Leadership and supervision, Metadata curation, and Project administration:**

Patrick J Lillie ^37^

**Leadership and supervision, Metadata curation, and Samples and logistics:**

Nicholas Cortes ^33^, Nathan Moore ^33^, Claire Thomas ^33^, Phillipa J Burns ^37^, Tabitha W Mahungu ^80^ and Steven Liggett ^86^

**Leadership and supervision, Metadata curation, and Sequencing and analysis:**

Angela H Beckett ^13, 81^ and Matthew TG Holden ^73^

**Leadership and supervision, Project administration, and Samples and logistics:**

Lisa J Levett ^34^, Husam Osman ^70, 35^ and Mohammed O Hassan-Ibrahim ^99^

**Leadership and supervision, Project administration, and Sequencing and analysis:**

David A Simpson ^77^

**Leadership and supervision, Samples and logistics, and Sequencing and analysis:**

Meera Chand ^72^, Ravi K Gupta ^102^, Alistair C Darby ^107^ and Steve Paterson ^107^

**Leadership and supervision, Sequencing and analysis, and Software and analysis tools:**

Oliver G Pybus ^23^, Erik M Volz ^39^, Daniela de Angelis ^52^, David L Robertson ^53^, Andrew J Page ^75^ and Inigo Martincorena ^116^

**Leadership and supervision, Sequencing and analysis, and Visualisation:**

Louise Aigrain ^116^ and Andrew R Bassett ^116^

**Metadata curation, Project administration, and Samples and logistics:**

Nick Wong ^50^, Yusri Taha ^89^, Michelle J Erkiert ^99^ and Michael H Spencer Chapman ^116, 102^

**Metadata curation, Project administration, and Sequencing and analysis:**

Rebecca Dewar ^56^ and Martin P McHugh ^56, 111^

**Metadata curation, Project administration, and Software and analysis tools:**

Siddharth Mookerjee ^38, 57^

**Metadata curation, Project administration, and Visualisation:**

Stephen Aplin ^97^, Matthew Harvey ^97^, Thea Sass ^97^, Helen Umpleby ^97^ and Helen Wheeler ^97^

**Metadata curation, Samples and logistics, and Sequencing and analysis:**

James P McKenna ^3^, Ben Warne ^9^, Joshua F Taylor ^22^, Yasmin Chaudhry ^24^, Rhys Izuagbe ^24^, Aminu S Jahun ^24^, Gregory R Young ^36, 65^, Claire McMurray ^43^, Clare M McCann ^65, 66^, Andrew Nelson ^65, 66^ and Scott Elliott ^68^

**Metadata curation, Samples and logistics, and Visualisation:**

Hannah Lowe ^25^

**Metadata curation, Sequencing and analysis, and Software and analysis tools:**

Anna Price ^11^, Matthew R Crown ^65^, Sara Rey ^74^, Sunando Roy ^96^ and Ben Temperton ^105^

**Metadata curation, Sequencing and analysis, and Visualisation:**

Sharif Shaaban ^73^ and Andrew R Hesketh ^101^

**Project administration, Samples and logistics, and Sequencing and analysis:**

Kenneth G Laing ^41^, Irene M Monahan ^41^ and Judith Heaney ^95, 96, 34^

**Project administration, Samples and logistics, and Visualisation:**

Emanuela Pelosi ^97^, Siona Silviera ^97^ and Eleri Wilson-Davies ^97^

**Samples and logistics, Software and analysis tools, and Visualisation:**

Helen Fryer ^5^

**Sequencing and analysis, Software and analysis tools, and Visualization:**

Helen Adams ^4^, Louis du Plessis ^23^, Rob Johnson ^39^, William T Harvey ^53, 42^, Joseph Hughes ^53^, Richard J Orton ^53^, Lewis G Spurgin ^59^, Yann Bourgeois ^81^, Chris Ruis ^102^, Áine O’Toole ^104^, Marina Gourtovaia ^116^ and Theo Sanderson ^116^

**Funding acquisition, and Leadership and supervision:**

Christophe Fraser ^5^, Jonathan Edgeworth ^12^, Judith Breuer ^96, 29^, Stephen L Michell ^105^ and John A Todd ^115^

**Funding acquisition, and Project administration:**

Michaela John ^10^ and David Buck ^115^

**Leadership and supervision, and Metadata curation:**

Kavitha Gajee ^37^ and Gemma L Kay ^75^

**Leadership and supervision, and Project administration:**

Sharon J Peacock ^20, 70^ and David Heyburn ^74^

**Leadership and supervision, and Samples and logistics:**

Katie Kitchman ^37^, Alan McNally ^43, 93^, David T Pritchard ^50^, Samir Dervisevic ^58^, Peter Muir ^70^, Esther Robinson ^70, 35^, Barry B Vipond ^70^, Newara A Ramadan ^78^, Christopher Jeanes ^90^, Danni Weldon ^116^, Jana Catalan ^118^ and Neil Jones ^118^

**Leadership and supervision, and Sequencing and analysis:**

Ana da Silva Filipe ^53^, Chris Williams ^74^, Marc Fuchs ^77^, Julia Miskelly ^77^, Aaron R Jeffries ^105^, Karen Oliver ^116^ and Naomi R Park ^116^

**Metadata curation, and Samples and logistics:**

Amy Ash ^1^, Cherian Koshy ^1^, Magdalena Barrow ^7^, Sarah L Buchan ^7^, Anna Mantzouratou ^7^, Gemma Clark ^15^, Christopher W Holmes ^16^, Sharon Campbell ^17^, Thomas Davis ^21^, Ngee Keong Tan ^22^, Julianne R Brown ^29^, Kathryn A Harris ^29, 2^, Stephen P Kidd ^33^, Paul R Grant ^34^, Li Xu-McCrae ^35^, Alison Cox ^38, 63^, Pinglawathee Madona ^38, 63^, Marcus Pond ^38, 63^, Paul A Randell ^38, 63^, Karen T Withell ^48^, Cheryl Williams ^51^, Clive Graham ^60^, Rebecca Denton-Smith ^62^, Emma Swindells ^62^, Robyn Turnbull ^62^, Tim J Sloan ^67^, Andrew Bosworth ^70, 35^, Stephanie Hutchings ^70^, Hannah M Pymont ^70^, Anna Casey ^76^, Liz Ratcliffe ^76^, Christopher R Jones ^79, 105^, Bridget A Knight ^79, 105^, Tanzina Haque ^80^, Jennifer Hart ^80^, Dianne Irish-Tavares ^80^, Eric Witele ^80^, Craig Mower ^86^, Louisa K Watson ^86^, Jennifer Collins ^89^, Gary Eltringham ^89^, Dorian Crudgington ^98^, Ben Macklin ^98^, Miren Iturriza-Gomara ^107^, Anita O Lucaci ^107^ and Patrick C McClure ^113^

**Metadata curation, and Sequencing and analysis:**

Matthew Carlile ^18^, Nadine Holmes ^18^, Christopher Moore ^18^, Nathaniel Storey ^29^, Stefan Rooke ^73^, Gonzalo Yebra ^73^, Noel Craine ^74^, Malorie Perry ^74^, Nabil-Fareed Alikhan ^75^, Stephen Bridgett ^77^, Kate F Cook ^84^, Christopher Fearn ^84^, Salman Goudarzi ^84^, Ronan A Lyons ^88^, Thomas Williams ^104^, Sam T Haldenby ^107^, Jillian Durham ^116^ and Steven Leonard ^116^

**Metadata curation, and Software and analysis tools:**

Robert M Davies ^116^

**Project administration, and Samples and logistics:**

Rahul Batra ^12^, Beth Blane ^20^, Moira J Spyer ^30, 95, 96^, Perminder Smith ^32, 112^, Mehmet Yavus ^85, 109^, Rachel J Williams ^96^, Adhyana IK Mahanama ^97^, Buddhini Samaraweera ^97^, Sophia T Girgis ^102^, Samantha E Hansford ^109^, Angie Green ^115^, Charlotte Beaver ^116^, Katherine L Bellis ^116, 102^, Matthew J Dorman ^116^, Sally Kay ^116^, Liam Prestwood ^116^ and Shavanthi Rajatileka ^116^

**Project administration, and Sequencing and analysis:**

Joshua Quick ^43^

**Project administration, and Software and analysis tools:**

Radoslaw Poplawski ^43^

**Samples and logistics, and Sequencing and analysis:**

Nicola Reynolds ^8^, Andrew Mack ^11^, Arthur Morriss ^11^, Thomas Whalley ^11^, Bindi Patel ^12^, Iliana Georgana ^24^, Myra Hosmillo^24^, Malte L Pinckert ^24^, Joanne Stockton ^43^, John H Henderson ^65^, Amy Hollis ^65^, William Stanley ^65^, Wen C Yew ^65^, Richard Myers ^72^, Alicia Thornton ^72^, Alexander Adams ^74^, Tara Annett ^74^, Hibo Asad ^74^, Alec Birchley ^74^, Jason Coombes ^74^, Johnathan M Evans ^74^, Laia Fina ^74^, Bree Gatica-Wilcox ^74^, Lauren Gilbert ^74^, Lee Graham ^74^, Jessica Hey ^74^, Ember Hilvers ^74^, Sophie Jones ^74^, Hannah Jones ^74^, Sara Kumziene-Summerhayes ^74^, Caoimhe McKerr ^74^, Jessica Powell ^74^, Georgia Pugh ^74^, Sarah Taylor ^74^, Alexander J Trotter ^75^, Charlotte A Williams ^96^, Leanne M Kermack ^102^, Benjamin H Foulkes ^109^, Marta Gallis ^109^, Hailey R Hornsby ^109^, Stavroula F Louka ^109^, Manoj Pohare ^109^, Paige Wolverson ^109^, Peijun Zhang ^109^, George MacIntyre-Cockett ^115^, Amy Trebes ^115^, Robin J Moll ^116^, Lynne Ferguson ^117^, Emily J Goldstein ^117^, Alasdair Maclean ^117^ and Rachael Tomb ^117^

**Samples and logistics, and Software and analysis tools:**

Igor Starinskij ^53^

**Sequencing and analysis, and Software and analysis tools:**

Laura Thomson ^5^, Joel Southgate ^11, 74^, Moritz UG Kraemer ^23^, Jayna Raghwani ^23^, Alex E Zarebski ^23^, Olivia Boyd ^39^, Lily Geidelberg ^39^, Chris J Illingworth ^52^, Chris Jackson ^52^, David Pascall ^52^, Sreenu Vattipally ^53^, Timothy M Freeman ^109^, Sharon N Hsu ^109^, Benjamin B Lindsey ^109^, Keith James ^116^, Kevin Lewis ^116^, Gerry Tonkin-Hill ^116^ and Jaime M Tovar-Corona ^116^

**Sequencing and analysis, and Visualisation:**

MacGregor Cox ^20^

**Software and analysis tools, and Visualisation:**

Khalil Abudahab ^14, 116^, Mirko Menegazzo ^14^, Ben EW Taylor MEng ^14, 116^, Corin A Yeats ^14^, Afrida Mukaddas ^53^, Derek W Wright ^53^, Leonardo de Oliveira Martins ^75^, Rachel Colquhoun ^104^, Verity Hill ^104^, Ben Jackson ^104^, JT McCrone ^104^, Nathan Medd ^104^, Emily Scher ^104^ and Jon-Paul Keatley ^116^

**Leadership and supervision:**

Tanya Curran ^3^, Sian Morgan ^10^, Patrick Maxwell ^20^, Ken Smith ^20^, Sahar Eldirdiri ^21^, Anita Kenyon ^21^, Alison H Holmes ^38, 57^, James R Price ^38, 57^, Tim Wyatt ^69^, Alison E Mather ^75^, Timofey Skvortsov ^77^ and John A Hartley ^96^

**Metadata curation:**

Martyn Guest ^11^, Christine Kitchen ^11^, Ian Merrick ^11^, Robert Munn ^11^, Beatrice Bertolusso ^33^, Jessica Lynch ^33^, Gabrielle Vernet ^33^, Stuart Kirk ^34^, Elizabeth Wastnedge ^56^, Rachael Stanley ^58^, Giles Idle ^64^, Declan T Bradley ^69, 77^, Jennifer Poyner ^79^ and Matilde Mori ^110^

**Project administration:**

Owen Jones ^11^, Victoria Wright ^18^, Ellena Brooks ^20^, Carol M Churcher ^20^, Mireille Fragakis ^20^, Katerina Galai ^20, 70^, Andrew Jermy ^20^, Sarah Judges ^20^, Georgina M McManus ^20^, Kim S Smith ^20^, Elaine Westwick ^20^, Stephen W Attwood ^23^, Frances Bolt ^38, 57^, Alisha Davies ^74^, Elen De Lacy ^74^, Fatima Downing ^74^, Sue Edwards ^74^, Lizzie Meadows ^75^, Sarah Jeremiah ^97^, Nikki Smith ^109^ and Luke Foulser ^116^

**Samples and logistics:**

Themoula Charalampous ^12, 46^, Amita Patel ^12^, Louise Berry ^15^, Tim Boswell ^15^, Vicki M Fleming ^15^, Hannah C Howson-Wells ^15^, Amelia Joseph ^15^, Manjinder Khakh ^15^, Michelle M Lister ^15^, Paul W Bird ^16^, Karlie Fallon ^16^, Thomas Helmer ^16^, Claire L McMurray ^16^, Mina Odedra ^16^, Jessica Shaw ^16^, Julian W Tang ^16^, Nicholas J Willford ^16^, Victoria Blakey ^17^, Veena Raviprakash ^17^, Nicola Sheriff ^17^, Lesley-Anne Williams ^17^, Theresa Feltwell ^20^, Luke Bedford ^26^, James S Cargill ^27^, Warwick Hughes ^27^, Jonathan Moore ^28^, Susanne Stonehouse ^28^, Laura Atkinson ^29^, Jack CD Lee ^29^, Dr Divya Shah ^29^, Adela Alcolea-Medina ^32, 112^, Natasha Ohemeng-Kumi ^32, 112^, John Ramble ^32, 112^, Jasveen Sehmi ^32, 112^, Rebecca Williams ^33^, Wendy Chatterton ^34^, Monika Pusok ^34^, William Everson ^37^, Anibolina Castigador ^44^, Emily Macnaughton ^44^, Kate El Bouzidi ^45^, Temi Lampejo ^45^, Malur Sudhanva ^45^, Cassie Breen ^47^, Graciela Sluga ^48^, Shazaad SY Ahmad ^49, 70^, Ryan P George ^49^, Nicholas W Machin ^49, 70^, Debbie Binns ^50^, Victoria James ^50^, Rachel Blacow ^55^, Lindsay Coupland ^58^, Louise Smith ^59^, Edward Barton ^60^, Debra Padgett ^60^, Garren Scott ^60^, Aidan Cross ^61^, Mariyam Mirfenderesky ^61^, Jane Greenaway ^62^, Kevin Cole ^64^, Phillip Clarke ^67^, Nichola Duckworth ^67^, Sarah Walsh ^67^, Kelly Bicknell ^68^, Robert Impey ^68^, Sarah Wyllie ^68^, Richard Hopes ^70^, Chloe Bishop ^72^, Vicki Chalker ^72^, Ian Harrison ^72^, Laura Gifford ^74^, Zoltan Molnar ^77^, Cressida Auckland ^79^, Cariad Evans ^85, 109^, Kate Johnson ^85, 109^, David G Partridge ^85, 109^, Mohammad Raza ^85, 109^, Paul Baker ^86^, Stephen Bonner ^86^, Sarah Essex ^86^, Leanne J Murray ^86^, Andrew I Lawton ^87^, Shirelle Burton-Fanning ^89^, Brendan AI Payne ^89^, Sheila Waugh ^89^, Andrea N Gomes ^91^, Maimuna Kimuli ^91^, Darren R Murray ^91^, Paula Ashfield ^92^, Donald Dobie ^92^, Fiona Ashford ^93^, Angus Best ^93^, Liam Crawford ^93^, Nicola Cumley ^93^, Megan Mayhew ^93^, Oliver Megram ^93^, Jeremy Mirza ^93^, Emma Moles-Garcia ^93^, Benita Percival ^93^, Megan Driscoll ^96^, Leah Ensell ^96^, Helen L Lowe ^96^, Laurentiu Maftei ^96^, Matteo Mondani ^96^, Nicola J Chaloner ^99^, Benjamin J Cogger ^99^, Lisa J Easton ^99^, Hannah Huckson ^99^, Jonathan Lewis ^99^, Sarah Lowdon ^99^, Cassandra S Malone ^99^, Florence Munemo ^99^, Manasa Mutingwende ^99^, Roberto Nicodemi ^99^, Olga Podplomyk ^99^, Thomas Somassa ^99^, Andrew Beggs ^100^, Alex Richter ^100^, Claire Cormie ^102^, Joana Dias ^102^, Sally Forrest ^102^, Ellen E Higginson ^102^, Mailis Maes ^102^, Jamie Young ^102^, Rose K Davidson ^103^, Kathryn A Jackson ^107^, Lance Turtle ^107^, Alexander J Keeley ^109^, Jonathan Ball ^113^, Timothy Byaruhanga ^113^, Joseph G Chappell ^113^, Jayasree Dey ^113^, Jack D Hill ^113^, Emily J Park ^113^, Arezou Fanaie ^114^, Rachel A Hilson ^114^, Geraldine Yaze ^114^ and Stephanie Lo ^116^

**Sequencing and analysis:**

Safiah Afifi ^10^, Robert Beer ^10^, Joshua Maksimovic ^10^, Kathryn McCluggage ^10^, Karla Spellman ^10^, Catherine Bresner ^11^, William Fuller ^11^, Angela Marchbank ^11^, Trudy Workman ^11^, Ekaterina Shelest ^13, 81^, Johnny Debebe ^18^, Fei Sang ^18^, Marina Escalera Zamudio ^23^, Sarah Francois ^23^, Bernardo Gutierrez ^23^, Tetyana I Vasylyeva ^23^, Flavia Flaviani ^31^, Manon Ragonnet-Cronin ^39^, Katherine L Smollett ^42^, Alice Broos ^53^, Daniel Mair ^53^, Jenna Nichols ^53^, Kyriaki Nomikou ^53^, Lily Tong ^53^, Ioulia Tsatsani ^53^, Sarah O’Brien ^54^, Steven Rushton ^54^, Roy Sanderson ^54^, Jon Perkins ^55^, Seb Cotton ^56^, Abbie Gallagher ^56^, Elias Allara ^70, 102^, Clare Pearson ^70, 102^, David Bibby ^72^, Gavin Dabrera ^72^, Nicholas Ellaby ^72^, Eileen Gallagher ^72^, Jonathan Hubb ^72^, Angie Lackenby ^72^, David Lee ^72^, Nikos Manesis ^72^, Tamyo Mbisa ^72^, Steven Platt ^72^, Katherine A Twohig ^72^, Mari Morgan ^74^, Alp Aydin ^75^, David J Baker ^75^, Ebenezer Foster-Nyarko ^75^, Sophie J Prosolek ^75^, Steven Rudder ^75^, Chris Baxter ^77^, Sílvia F Carvalho ^77^, Deborah Lavin ^77^, Arun Mariappan ^77^, Clara Radulescu ^77^, Aditi Singh ^77^, Miao Tang ^77^, Helen Morcrette ^79^, Nadua Bayzid ^96^, Marius Cotic ^96^, Carlos E Balcazar ^104^, Michael D Gallagher ^104^, Daniel Maloney ^104^, Thomas D Stanton ^104^, Kathleen A Williamson ^104^, Robin Manley ^105^, Michelle L Michelsen ^105^, Christine M Sambles ^105^, David J Studholme ^105^, Joanna Warwick-Dugdale ^105^, Richard Eccles ^107^, Matthew Gemmell ^107^, Richard Gregory ^107^, Margaret Hughes ^107^, Charlotte Nelson ^107^, Lucille Rainbow ^107^, Edith E Vamos ^107^, Hermione J Webster ^107^, Mark Whitehead ^107^, Claudia Wierzbicki ^107^, Adrienn Angyal ^109^, Luke R Green ^109^, Max Whiteley ^109^, Emma Betteridge ^116^, Iraad F Bronner ^116^, Ben W Farr ^116^, Scott Goodwin ^116^, Stefanie V Lensing ^116^, Shane A McCarthy ^116, 102^, Michael A Quail ^116^, Diana Rajan ^116^, Nicholas M Redshaw ^116^, Carol Scott ^116^, Lesley Shirley ^116^ and Scott AJ Thurston ^116^

**Software and analysis tools:**

Will Rowe ^43^, Amy Gaskin ^74^, Thanh Le-Viet ^75^, James Bonfield ^116^, Jennifier Liddle ^116^ and Andrew Whitwham ^116^

**1** Barking, Havering and Redbridge University Hospitals NHS Trust, **2** Barts Health NHS Trust, **3** Belfast Health & Social Care Trust, **4** Betsi Cadwaladr University Health Board, **5** Big Data Institute, Nuffield Department of Medicine, University of Oxford, **6** Blackpool Teaching Hospitals NHS Foundation Trust, **7** Bournemouth University, **8** Cambridge Stem Cell Institute, University of Cambridge, **9** Cambridge University Hospitals NHS Foundation Trust, **10** Cardiff and Vale University Health Board, **11** Cardiff University, **12** Centre for Clinical Infection and Diagnostics Research, Department of Infectious Diseases, Guy’s and St Thomas’ NHS Foundation Trust, **13** Centre for Enzyme Innovation, University of Portsmouth, **14** Centre for Genomic Pathogen Surveillance, University of Oxford, **15** Clinical Microbiology Department, Queens Medical Centre, Nottingham University Hospitals NHS Trust, **16** Clinical Microbiology, University Hospitals of Leicester NHS Trust, **17** County Durham and Darlington NHS Foundation Trust, **18** Deep Seq, School of Life Sciences, Queens Medical Centre, University of Nottingham, **19** Department of Infectious Diseases and Microbiology, Cambridge University Hospitals NHS Foundation Trust, **20** Department of Medicine, University of Cambridge, **21** Department of Microbiology, Kettering General Hospital, **22** Department of Microbiology, South West London Pathology, **23** Department of Zoology, University of Oxford, **24** Division of Virology, Department of Pathology, University of Cambridge, **25** East Kent Hospitals University NHS Foundation Trust, **26** East Suffolk and North Essex NHS Foundation Trust, **27** East Sussex Healthcare NHS Trust, **28** Gateshead Health NHS Foundation Trust, **29** Great Ormond Street Hospital for Children NHS Foundation Trust, **30** Great Ormond Street Institute of Child Health (GOS ICH), University College London (UCL), **31** Guy’s and St. Thomas’ Biomedical Research Centre, **32** Guy’s and St. Thomas’ NHS Foundation Trust, **33** Hampshire Hospitals NHS Foundation Trust, **34** Health Services Laboratories, **35** Heartlands Hospital, Birmingham, **36** Hub for Biotechnology in the Built Environment, Northumbria University, **37** Hull University Teaching Hospitals NHS Trust, **38** Imperial College Healthcare NHS Trust, **39** Imperial College London, **40** Infection Care Group, St George’s University Hospitals NHS Foundation Trust, **41** Institute for Infection and Immunity, St George’s University of London, **42** Institute of Biodiversity, Animal Health & Comparative Medicine, **43** Institute of Microbiology and Infection, University of Birmingham, **44** Isle of Wight NHS Trust, **45** King’s College Hospital NHS Foundation Trust, **46** King’s College London, **47** Liverpool Clinical Laboratories, **48** Maidstone and Tunbridge Wells NHS Trust, **49** Manchester University NHS Foundation Trust, **50** Microbiology Department, Buckinghamshire Healthcare NHS Trust, **51** Microbiology, Royal Oldham Hospital, **52** MRC Biostatistics Unit, University of Cambridge, **53** MRC-University of Glasgow Centre for Virus Research, **54** Newcastle University, **55** NHS Greater Glasgow and Clyde, **56** NHS Lothian, **57** NIHR Health Protection Research Unit in HCAI and AMR, Imperial College London, **58** Norfolk and Norwich University Hospitals NHS Foundation Trust, **59** Norfolk County Council, **60** North Cumbria Integrated Care NHS Foundation Trust, **61** North Middlesex University Hospital NHS Trust, **62** North Tees and Hartlepool NHS Foundation Trust, **63** North West London Pathology, **64** Northumbria Healthcare NHS Foundation Trust, **65** Northumbria University, **66** NU-OMICS, Northumbria University, **67** Path Links, Northern Lincolnshire and Goole NHS Foundation Trust, **68** Portsmouth Hospitals University NHS Trust, **69** Public Health Agency, Northern Ireland, **70** Public Health England, **71** Public Health England, Cambridge, **72** Public Health England, Colindale, **73** Public Health Scotland, **74** Public Health Wales, **75** Quadram Institute Bioscience, **76** Queen Elizabeth Hospital, Birmingham, **77** Queen’s University Belfast, **78** Royal Brompton and Harefield Hospitals, **79** Royal Devon and Exeter NHS Foundation Trust, **80** Royal Free London NHS Foundation Trust, **81** School of Biological Sciences, University of Portsmouth, **82** School of Health Sciences, University of Southampton, **83** School of Medicine, University of Southampton, **84** School of Pharmacy & Biomedical Sciences, University of Portsmouth, **85** Sheffield Teaching Hospitals NHS Foundation Trust, **86** South Tees Hospitals NHS Foundation Trust, **87** Southwest Pathology Services, **88** Swansea University, **89** The Newcastle upon Tyne Hospitals NHS Foundation Trust, **90** The Queen Elizabeth Hospital King’s Lynn NHS Foundation Trust, **91** The Royal Marsden NHS Foundation Trust, **92** The Royal Wolverhampton NHS Trust, **93** Turnkey Laboratory, University of Birmingham, **94** University College London Division of Infection and Immunity, **95** University College London Hospital Advanced Pathogen Diagnostics Unit, **96** University College London Hospitals NHS Foundation Trust, **97** University Hospital Southampton NHS Foundation Trust, **98** University Hospitals Dorset NHS Foundation Trust, **99** University Hospitals Sussex NHS Foundation Trust, **100** University of Birmingham, **101** University of Brighton, **102** University of Cambridge, **103** University of East Anglia, **104** University of Edinburgh, **105** University of Exeter, **106** University of Kent, **107** University of Liverpool, **108** University of Oxford, **109** University of Sheffield, **110** University of Southampton, **111** University of St Andrews, **112** Viapath, Guy’s and St Thomas’ NHS Foundation Trust, and King’s College Hospital NHS Foundation Trust, **113** Virology, School of Life Sciences, Queens Medical Centre, University of Nottingham, **114** Watford General Hospital, **115** Wellcome Centre for Human Genetics, Nuffield Department of Medicine, University of Oxford, **116** Wellcome Sanger Institute, **117** West of Scotland Specialist Virology Centre, NHS Greater Glasgow and Clyde, **118** Whittington Health NHS Trust

