## supplementary for "Recurrent SARS-CoV-2 Mutations in Immunodeficient Patients"

**Supplementary Material**

| ID | Source study ID | Time Covered (days) | N samples | Pango Lineage | Nature of Immune Deficiency | Citation | Genome Accessions |
|---|---|---|---|---|---|---|---|
| **1** | n/a | 121 | 2 | B.1.177 | Combined | Obtained from COG-UK Dataset | ERS6811185 ERS6814017 |
| **2** | n/a | 82 | 3 | B.1.1.7 | Combined | Obtained from COG-UK Dataset | ERS5942556 ERS6278124 ERS6812732 |
| **3** | n/a | 121 | 3 | B.1.1.7 | B cell dominant | Obtained from COG-UK Dataset | ERS6460486 ERS6460503 ERS6813850 |
| **4** | n/a | 39 | 2 | B.1.1.7 | Combined | Obtained from COG-UK Dataset | ERS5944756 ERS6279465 |
| **5** | n/a | 84 | 5 | B.1.1.7 | Combined | Obtained from COG-UK Dataset | ERS10290349 ERS6812591 ERS6201166 ERS6813151 ERS6369109 |
| **6** | n/a | 58 | 2 | B.1.1.7 | Combined | Obtained from COG-UK Dataset | ERS6203792 ERS6151321 |
| **7** | NA | 152 | 9 | B.1.486 | Combined | (Choi et al., 2020) | EPI_ISL_593480 EPI_ISL_593557 EPI_ISL_593558 EPI_ISL_593555 EPI_ISL_593478 EPI_ISL_593479 EPI_ISL_593556 EPI_ISL_593553 EPI_ISL_593554 |
| **8** | NA | 105 | 6 | A.1 | B cell dominant | (Avanzato et al., 2020) | MT982403 MT982402 MT982405 MT982406 MT982401 MT982404 |
| **9** | NA | ~120 | 2 | B.1 | B cell dominant | (Reuken et al., 2021) | EPI_ISL_632938 EPI_ISL_728323 |

| | | | | | | | |
|---|---|---|---|---|---|---|---|
| **10** | 1 | 56 | 4 | B.1 | T cell dominant | (Tarhini et al., 2021) | EPI_ISL_833191 EPI_ISL_833192 EPI_ISL_833193 EPI_ISL_833194 |
| **11** | 3 | 82 | 3 | B.1.356 | Combined | (Tarhini et al., 2021) | EPI_ISL_833199 EPI_ISL_833200 |
| **12** | X1 | 101 | 22 | B.1.1.1 | Combined | (Kemp et al., 2021) | ERS4673958 ERS4839477 ERS4840613 ERS4840145 ERS4841656 ERS4935380 ERS4935270 ERS4935069 ERS4935223 ERS4935334 ERS4952415 ERS4984866 ERS4984867 ERS4984887 ERS5052541 ERS5050904 ERS5050906 ERS5050902 ERS5050908 ERS5050894 ERS5050903 ERS6052283 |
| **13** | NA | | 9 | B.1 | B cell dominant | (Baang et al., 2021) | Available from Lauring lab github repository: https://github.com/lauringlab/ProlongedReplicationCase/blob/master/data/raw/all.consensus.fasta |
| **14** | NA | 153 | 7 | B.1.1.57 | B cell dominant | (Stanevich et al., 2021) | EPI_ISL_1039159 EPI_ISL_1372287 EPI_ISL_1372288 EPI_ISL_3029841 EPI_ISL_3029842 EPI_ISL_596228 |
| **15** | A | 134 | 10 | B.1.1.515 | B cell dominant | (Khatamzas et al., 2021) | EPI_ISL_732538 EPI_ISL_732658 |
| **16** | NA | 164 | 2 | B.1.1.401 | B cell dominant | (Borges et al., 2021) | EPI_ISL_941339 EPI_ISL_941340 |
| **17** | n/a | 61 | 6 | B.1.1.7 | T cell dominant | (Riddell et al., 2022) | ERS6333433 ERS6334312 ERS6334379 ERS6333918 ERS6279567 ERS6333855 |
| **18** | n/a | 111 | 10 | B.1.1.7 | T cell dominant | (Riddell et al., 2022) | ERS6334750 ERS6333015 ERS6332824 ERS6334364 |

| | | | | | | | |
|---|---|---|---|---|---|---|---|
| | | | | | | | ERS6334845 ERS6334159 ERS6333088 ERS6459881 ERS6812137 ERS6932781 |
| **19** | n/a | 229 | 16 | B.1.1.7 | T cell dominant | (Riddell et al., 2022) | ERS6334509 ERS6201120 ERS6333136 ERS6333653 ERS7549836 ERS6456939 ERS6457012 ERS6688703 ERS7543909 ERS7537861 ERS7546481 ERS7541762 ERS7452312 ERS7547817 ERS7452298 ERS7538781 ERS7975598 ERS8012725 ERS7539593 ERS7782693 |
| **20** | NA | 149 | 16 | A.2 | B cell dominant | (Ciuffreda et al., 2021) | EPI_ISL_7473140 EPI_ISL_7473130 EPI_ISL_7473141 EPI_ISL_7473135 EPI_ISL_7473136 EPI_ISL_7473137 EPI_ISL_7473138 EPI_ISL_7473131 EPI_ISL_7473142 EPI_ISL_7473132 EPI_ISL_7473143 EPI_ISL_7473133 EPI_ISL_7473144 EPI_ISL_7473134 EPI_ISL_7473139 EPI_ISL_7473129 |
| **21** | A | 14 | 5 | B.1 | Combined | (Jensen et al., 2021) | Available from corresponding author of citation. |
| **22** | B | 22 | 8 | B.1.1 | T cell dominant | (Jensen et al., 2021) | Available from corresponding author of citation. |
| **23** | C | 12 | 5 | B.1.177.62 | Combined | (Jensen et al., 2021) | Available from corresponding author of citation. |
| **24** | D | 8 | 3 | B.1.177.62 | Combined | (Jensen et al., 2021) | Available from corresponding author of citation. |

| | | | | | | | |
|---|---|---|---|---|---|---|---|
| **25** | E | 28 | 6 | B.1.258.14 | B cell dominant | (Jensen et al., 2021) | Available from corresponding author of citation. |
| **26** | F | 19 | 7 | B.1.160.14 | Combined | (Jensen et al., 2021) | Available from corresponding author of citation. |
| **27** | NA | 15 | 4 | B.1.1.7 | T cell dominant | (Riddell et al., 2022) | ERS7905450 ERS7844082 ERS7844642 ERS7981088 |
| **28** | NA | 140 | 9 | B.1.1 | Combined | (Weigang et al., 2021) | EPI_ISL_852806 EPI_ISL_852809 EPI_ISL_852807 EPI_ISL_852805 EPI_ISL_852808 EPI_ISL_852667 EPI_ISL_852659 EPI_ISL_852709 EPI_ISL_852671 |

**Subgroup Analysis Figures**

Patients were subdivided into groups based upon the details of their immunodeficiency but all groups lost power, figures based on this subgroup analysis are included below for posterity.

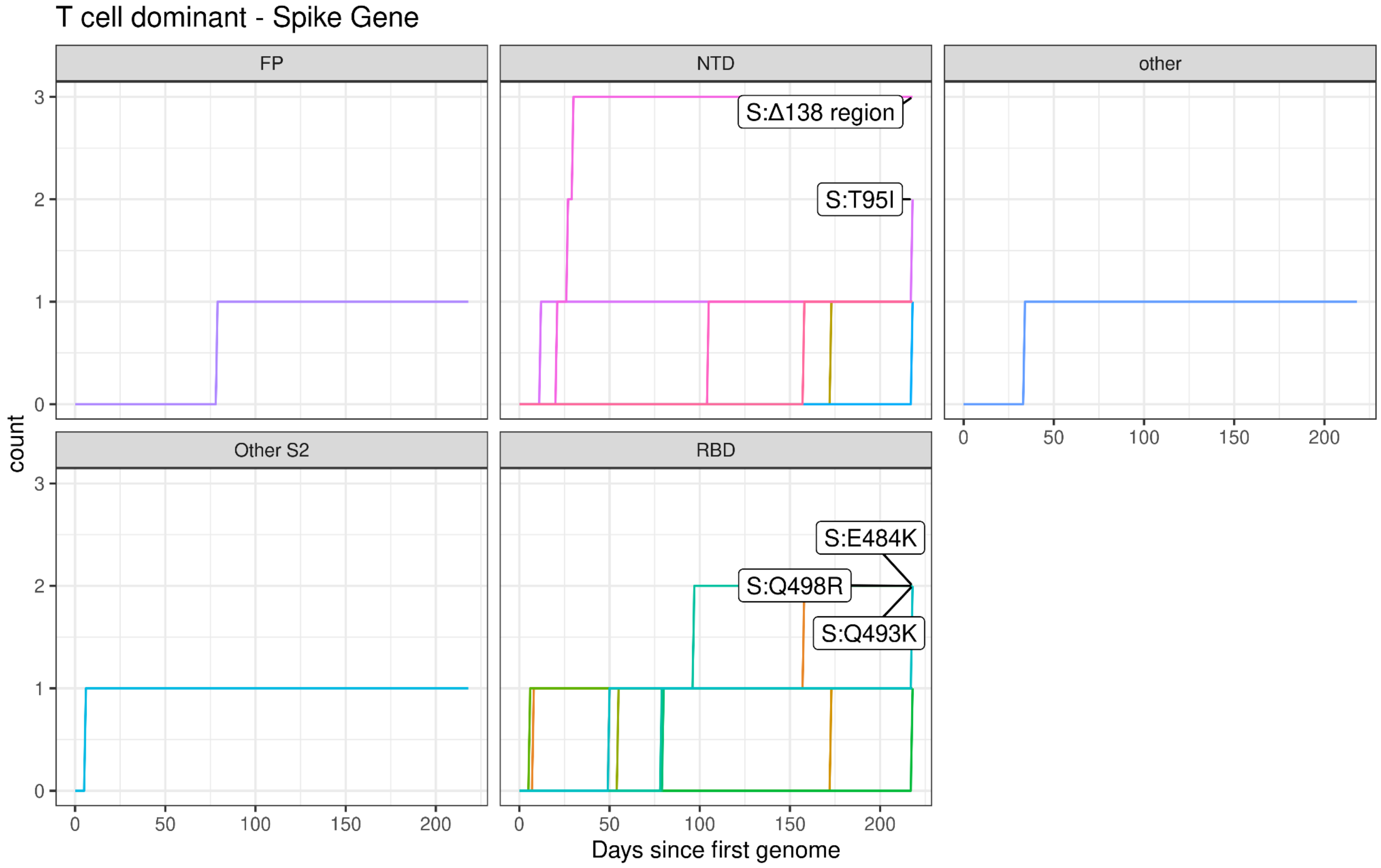

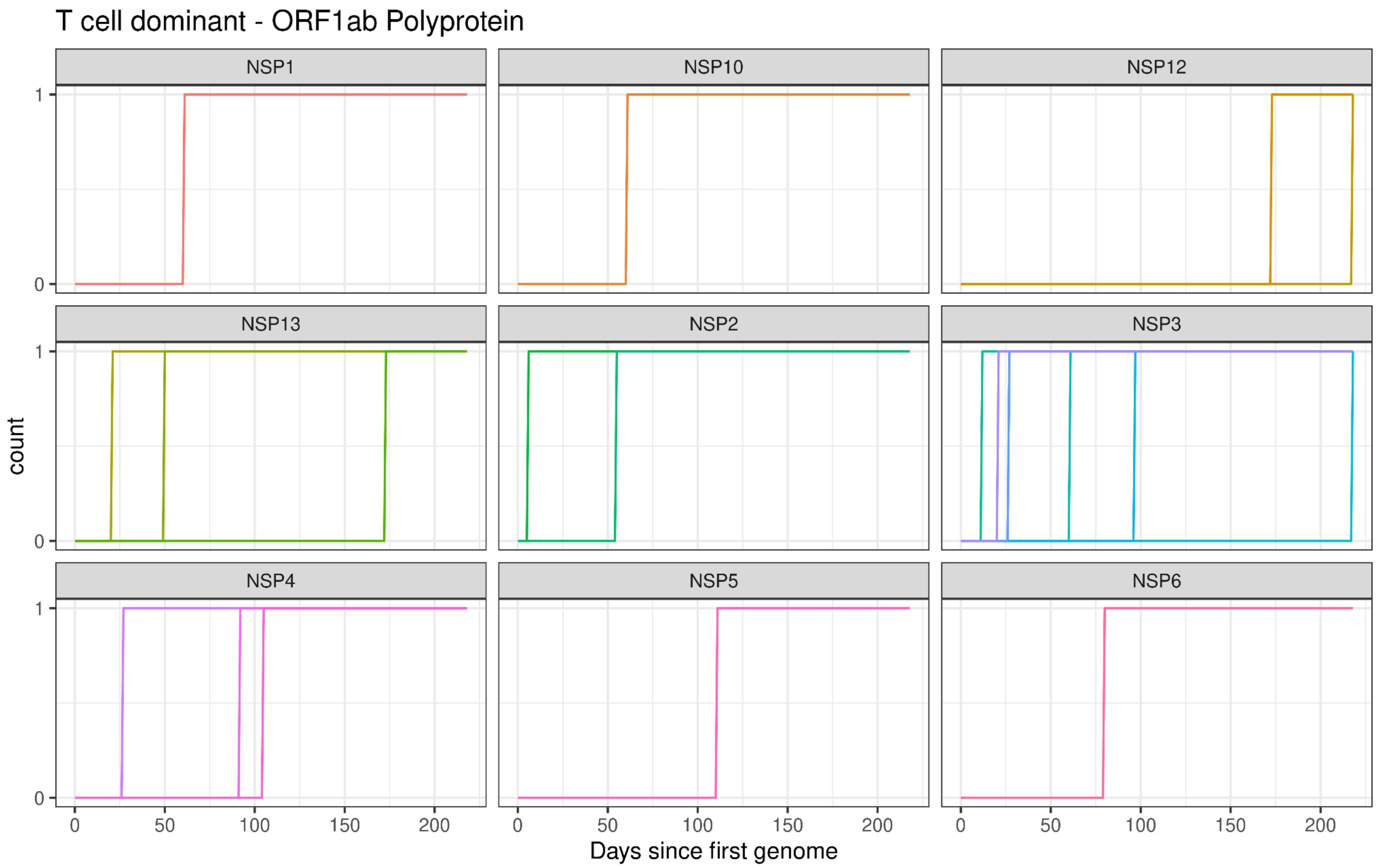
T cell dominant - ORF1ab Polyprotein
NSP1
NSP10
NSP12
NSP13
NSP2
NSP3
NSP4
NSP5
NSP6
count
0
1
0
50
100
150
200
Days since first genome

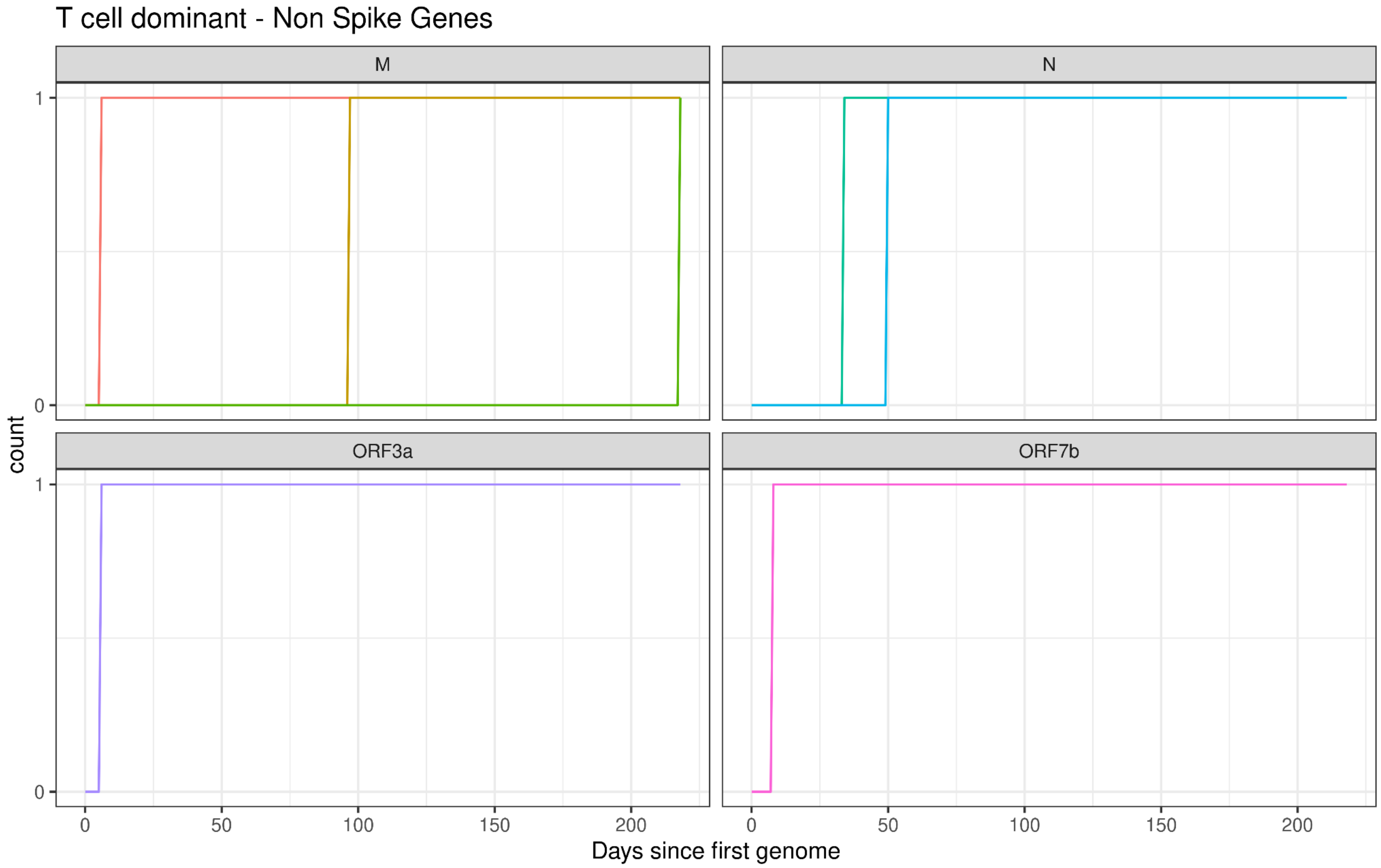
T cell dominant - Non Spike Genes
M
N
ORF3a
ORF7b
count
0
1
0
50
100
150
200
Days since first genome

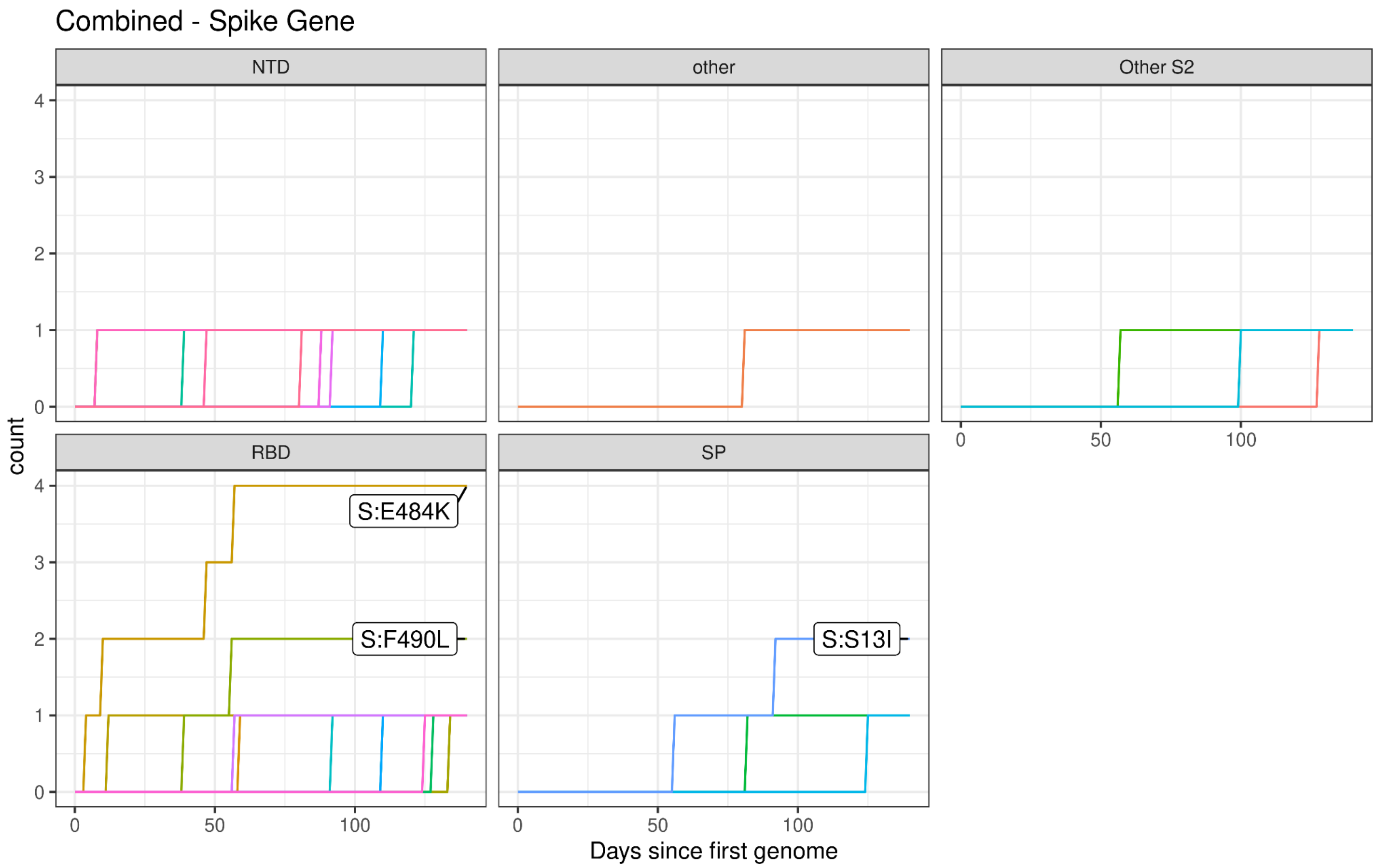

Combined - Spike Gene
NTD
other
Other S2
RBD
SP
S:E484K
S:F490L
S:S13I
count
Days since first genome


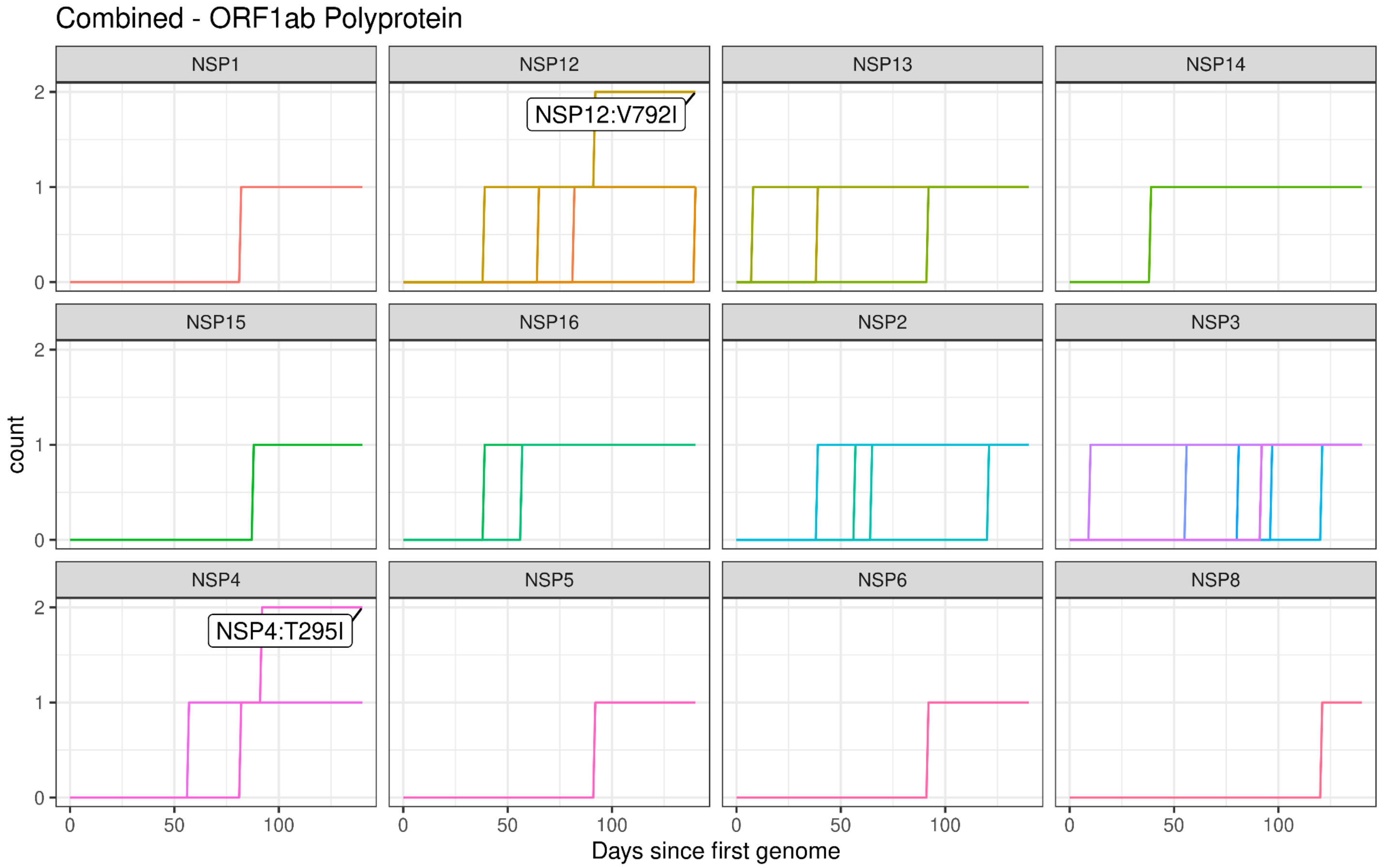

Combined - ORF1ab Polyprotein
NSP1
NSP12
NSP13
NSP14
NSP15
NSP16
NSP2
NSP3
NSP4
NSP5
NSP6
NSP8
NSP12:V792I
NSP4:T295I
count
Days since first genome

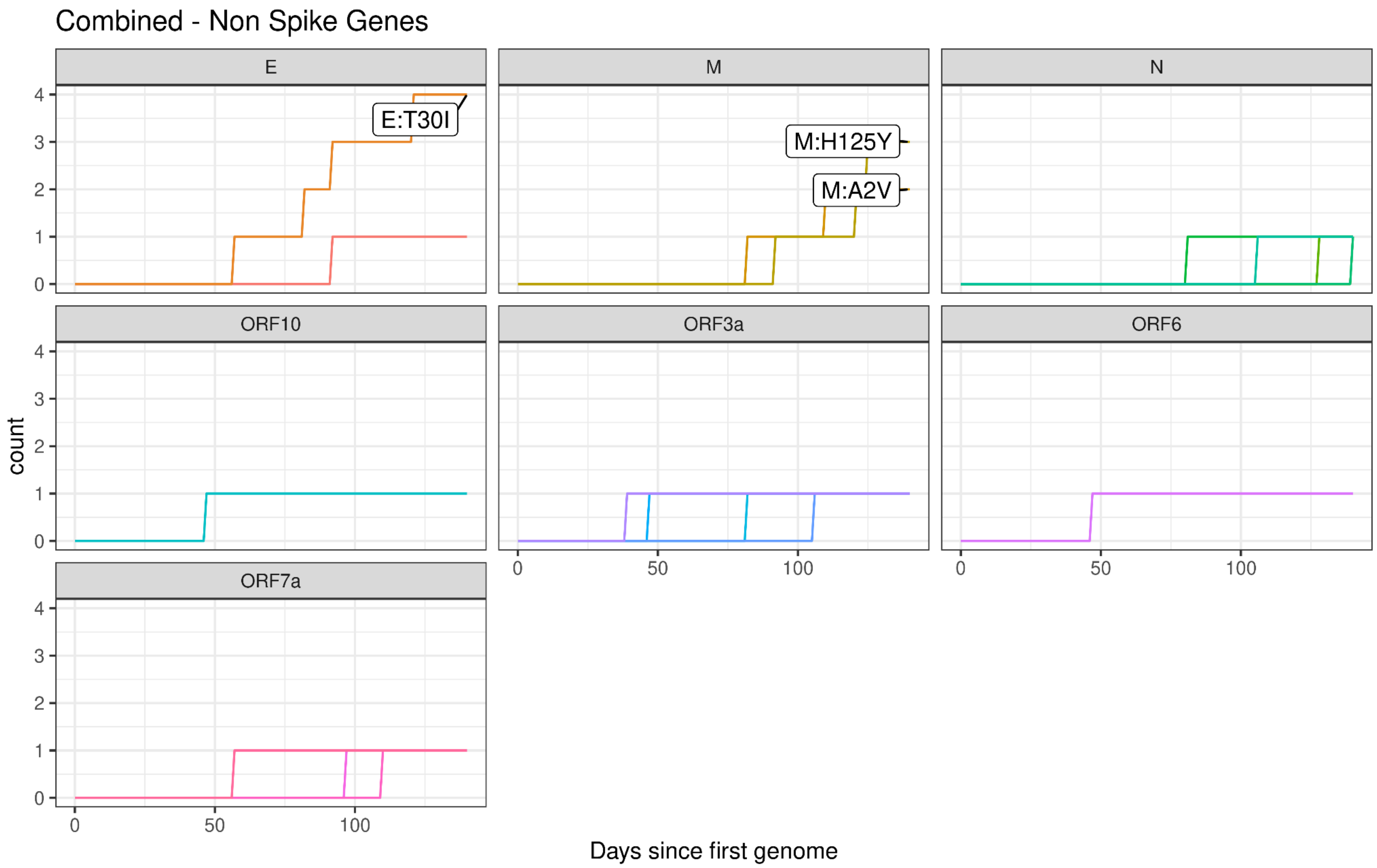
Combined - Non Spike Genes
E
M
N
ORF10
ORF3a
ORF6
ORF7a
E:T30I
M:H125Y
M:A2V
count
0
1
2
3
4
0
50
100
Days since first genome

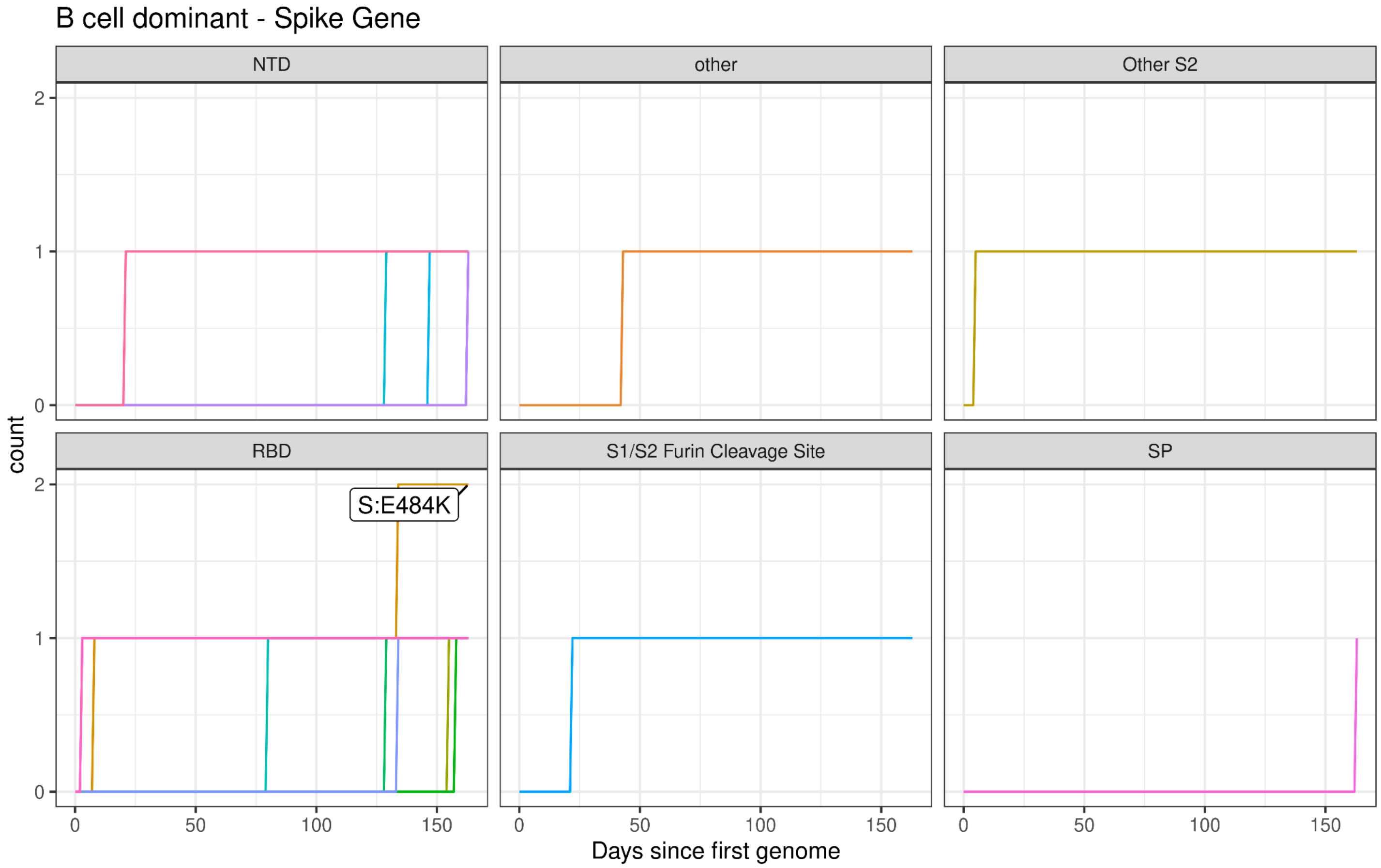
B cell dominant - Spike Gene
NTD
other
Other S2
RBD
S1/S2 Furin Cleavage Site
SP
S:E484K
count
0
1
2
0
50
100
150
Days since first genome

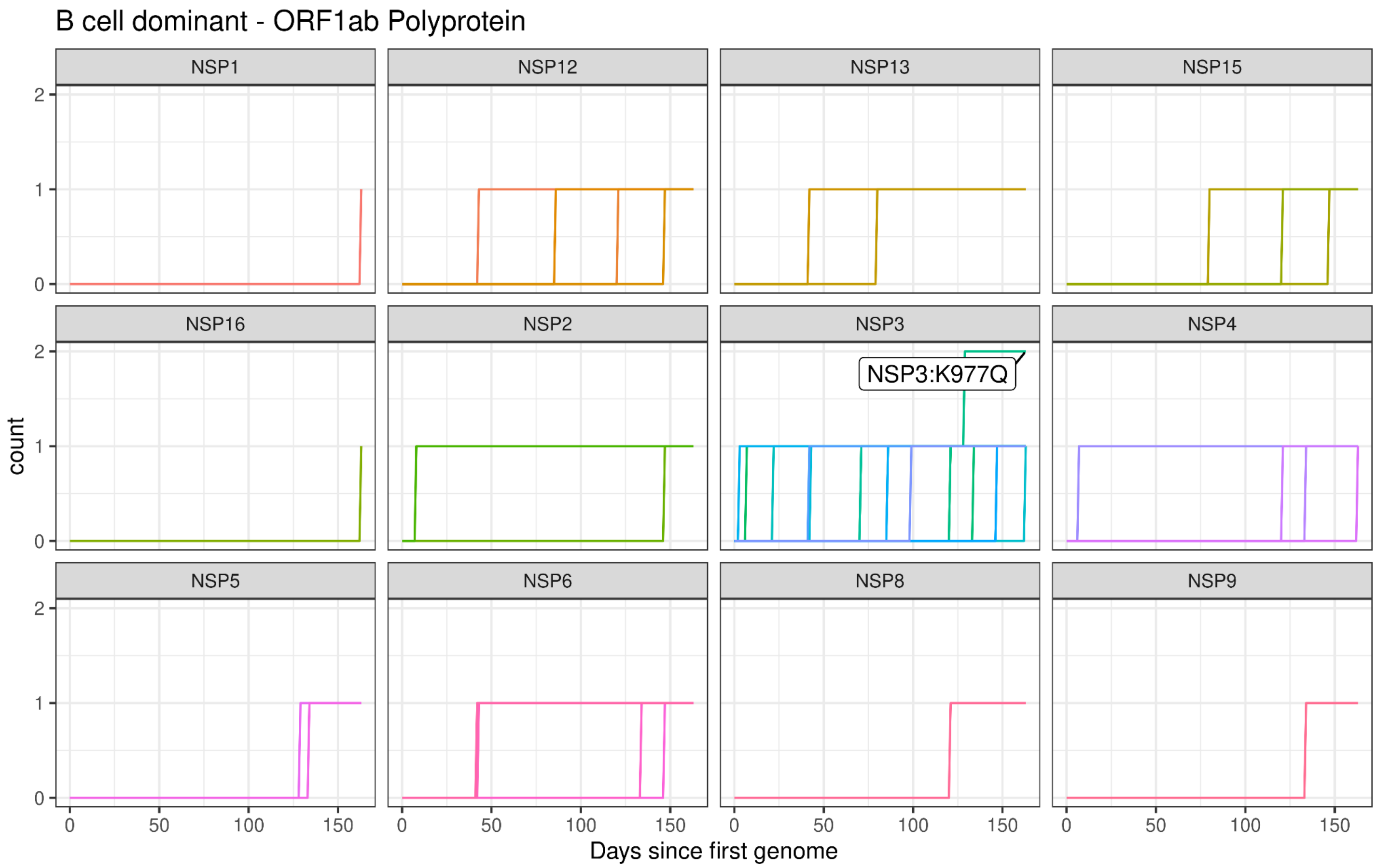
B cell dominant - ORF1ab Polyprotein
NSP1
NSP12
NSP13
NSP15
NSP16
NSP2
NSP3
NSP4
NSP5
NSP6
NSP8
NSP9
NSP3:K977Q
count
0
1
2
0
50
100
150
Days since first genome

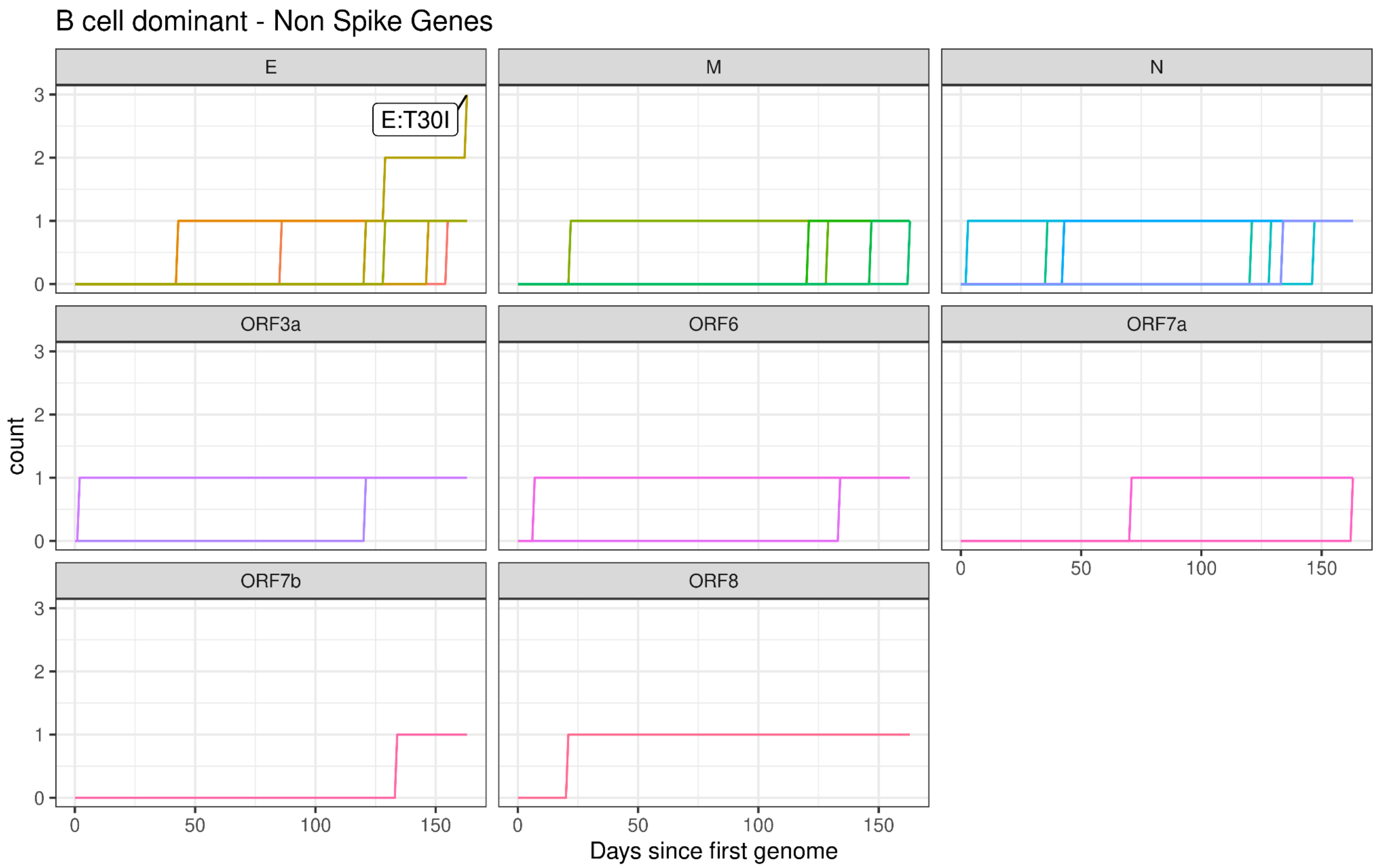
B cell dominant - Non Spike Genes
E
M
N
ORF3a
ORF6
ORF7a
ORF7b
ORF8
E:T30I
count
0
1
2
3
0
50
100
150
Days since first genome

**Civet Trees**

Trees generated by Civet (O'Toole et al., 2021a), red nodes indicate patient samples and in the case of patient 11 the blue node indicates a genome excluded from the dataset due to a probable superinfection.

Pt-1 Civet Tree

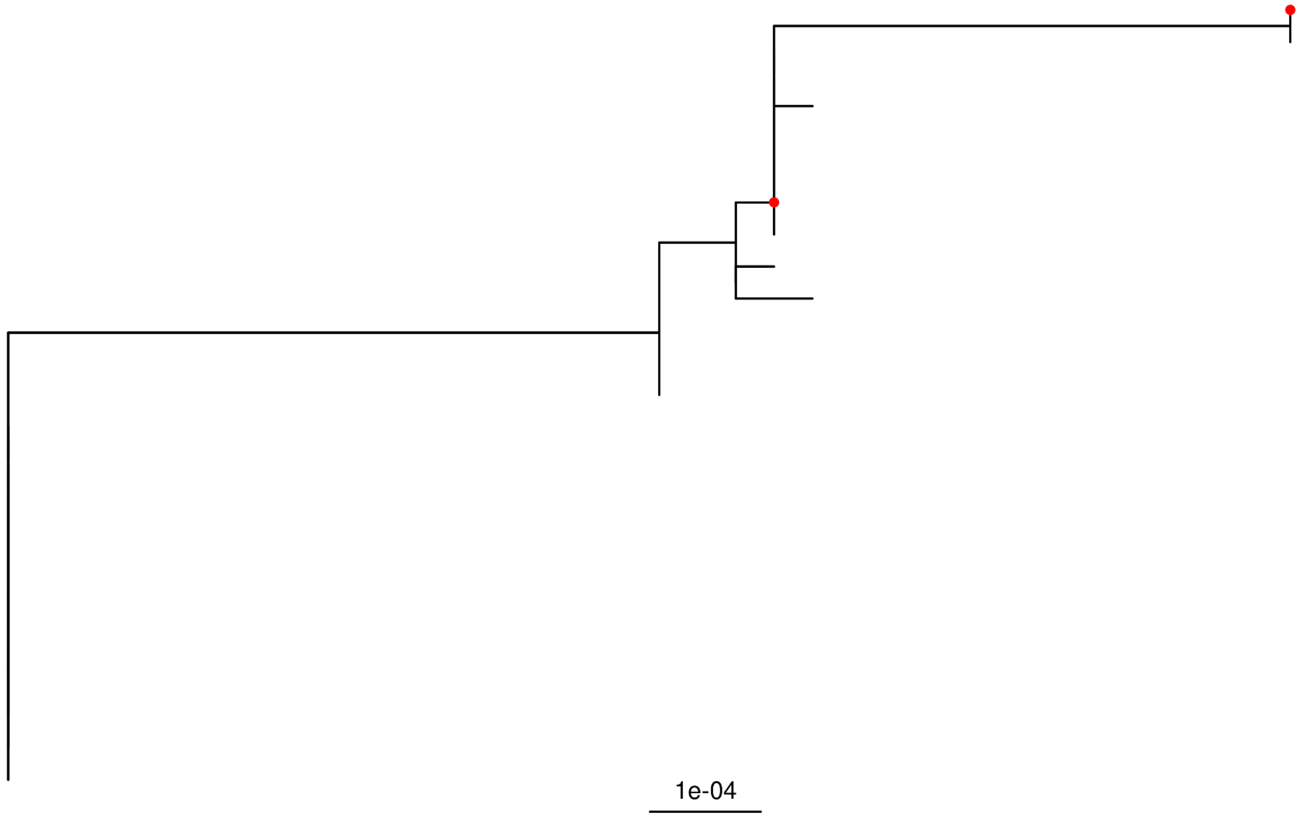

Pt-2 Civet Tree

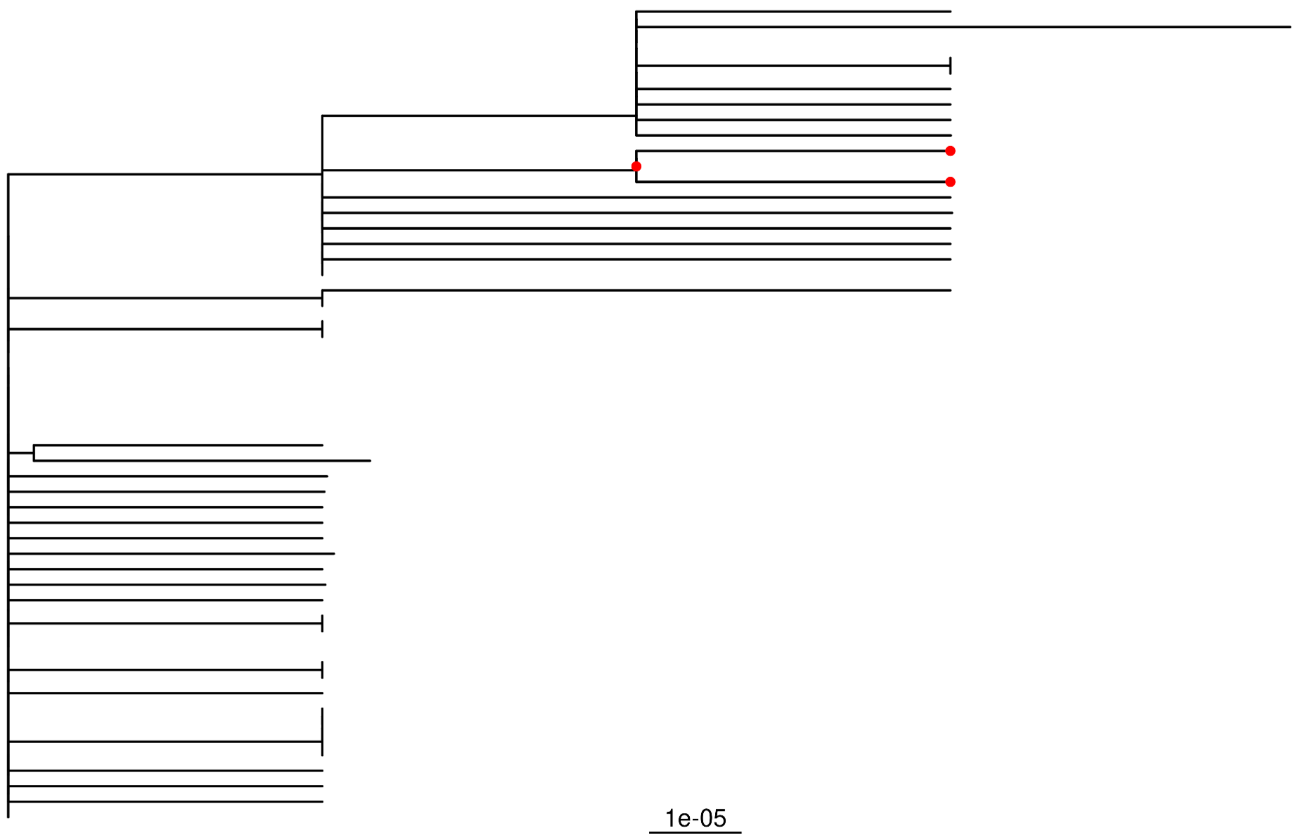


Pt-3 Civet Tree

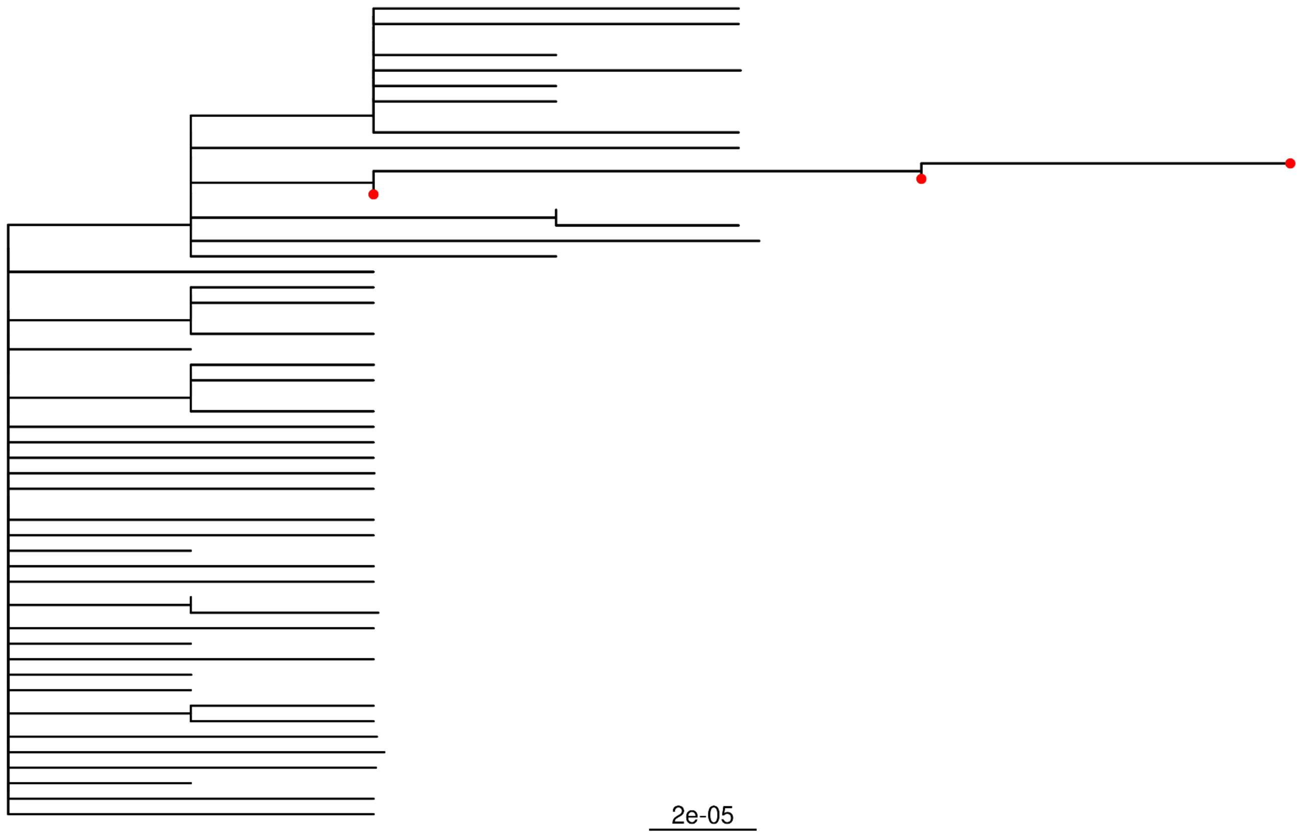

Pt-4 Civet Tree

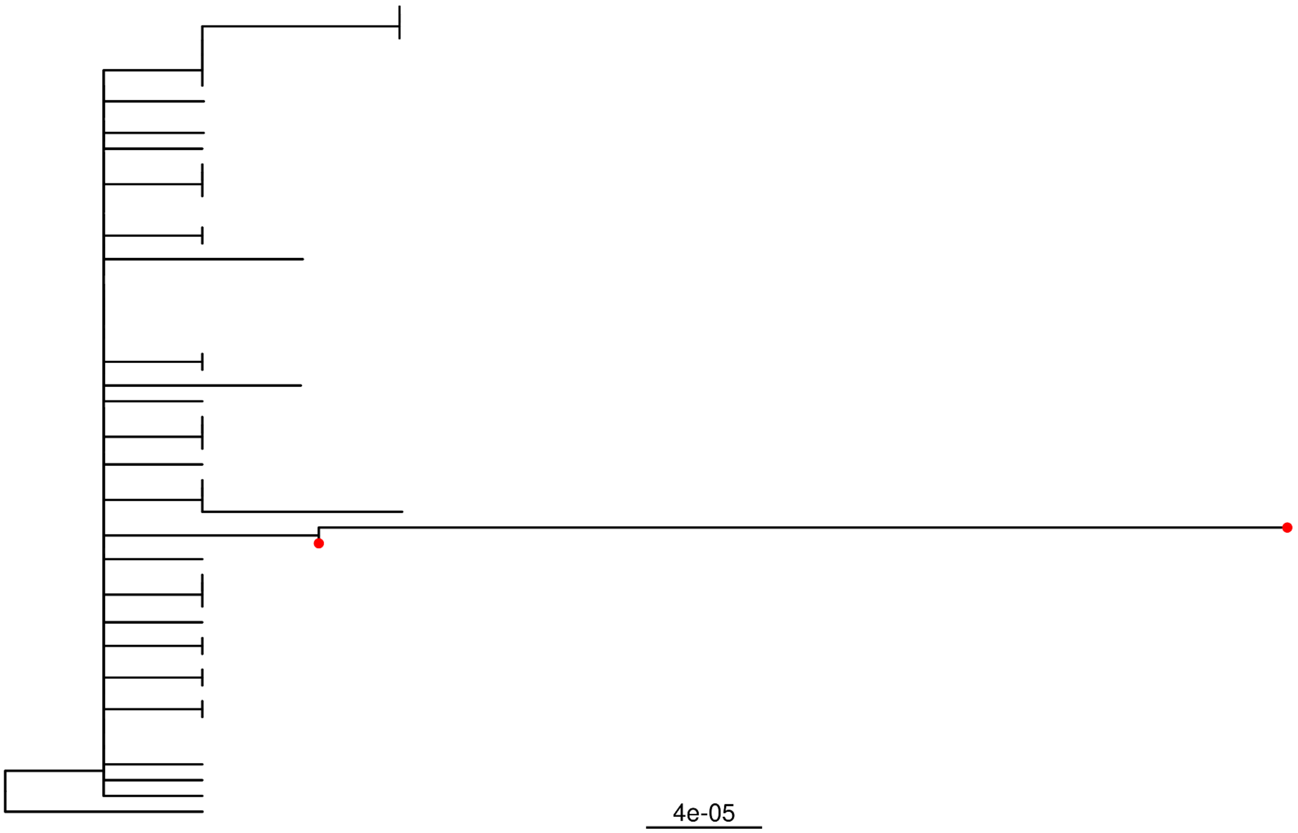


Pt-5 Civet Tree

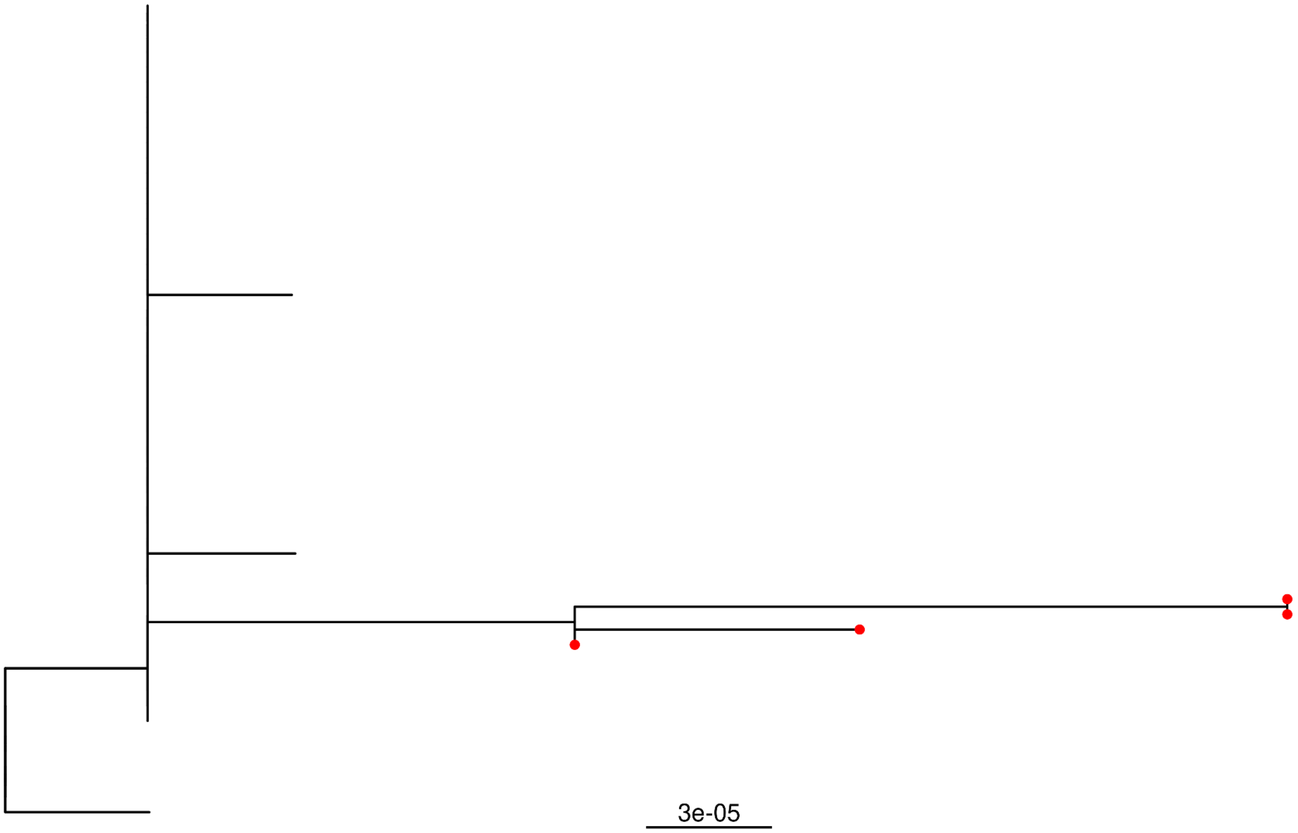

Pt-6 Civet Tree

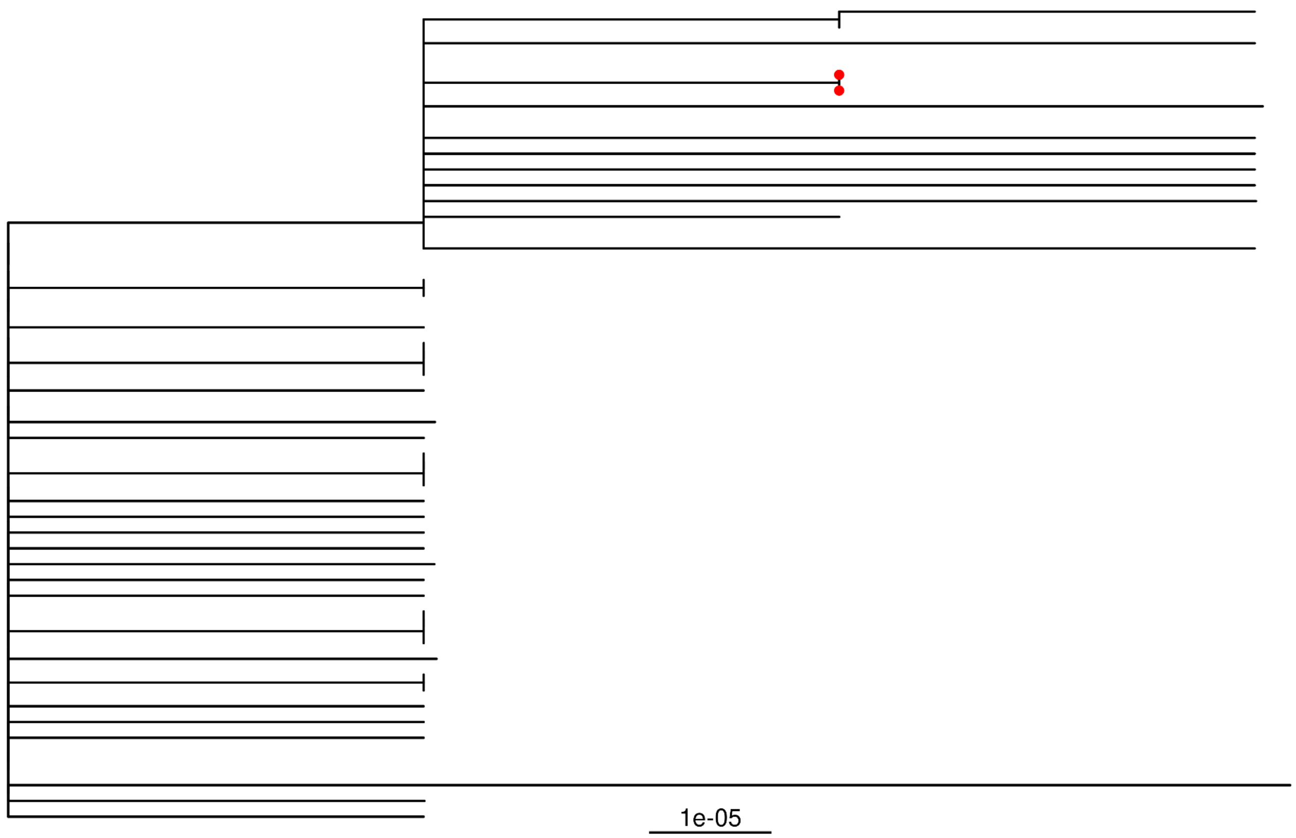


Pt-7 Civet Tree

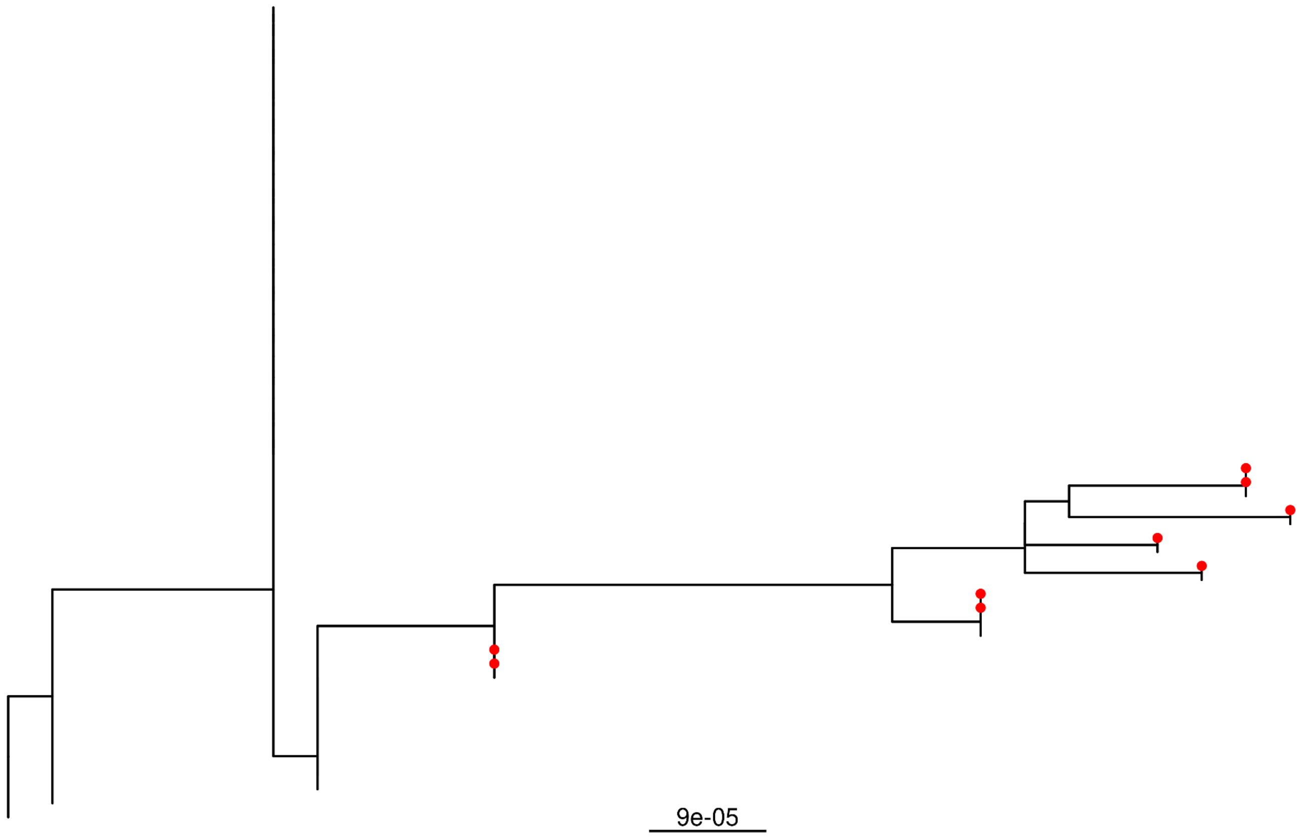

Pt-8 Civet Tree

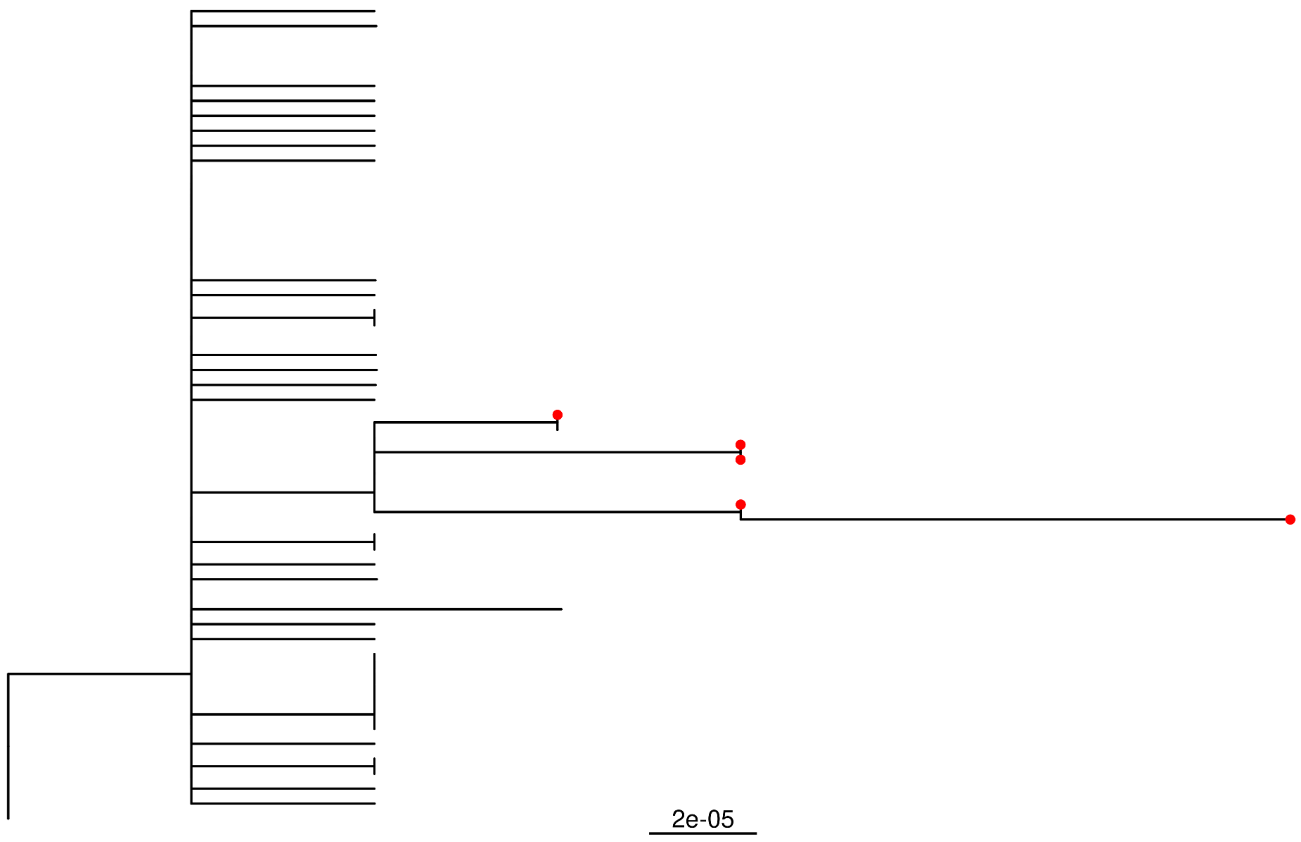


Pt-9 Civet Tree

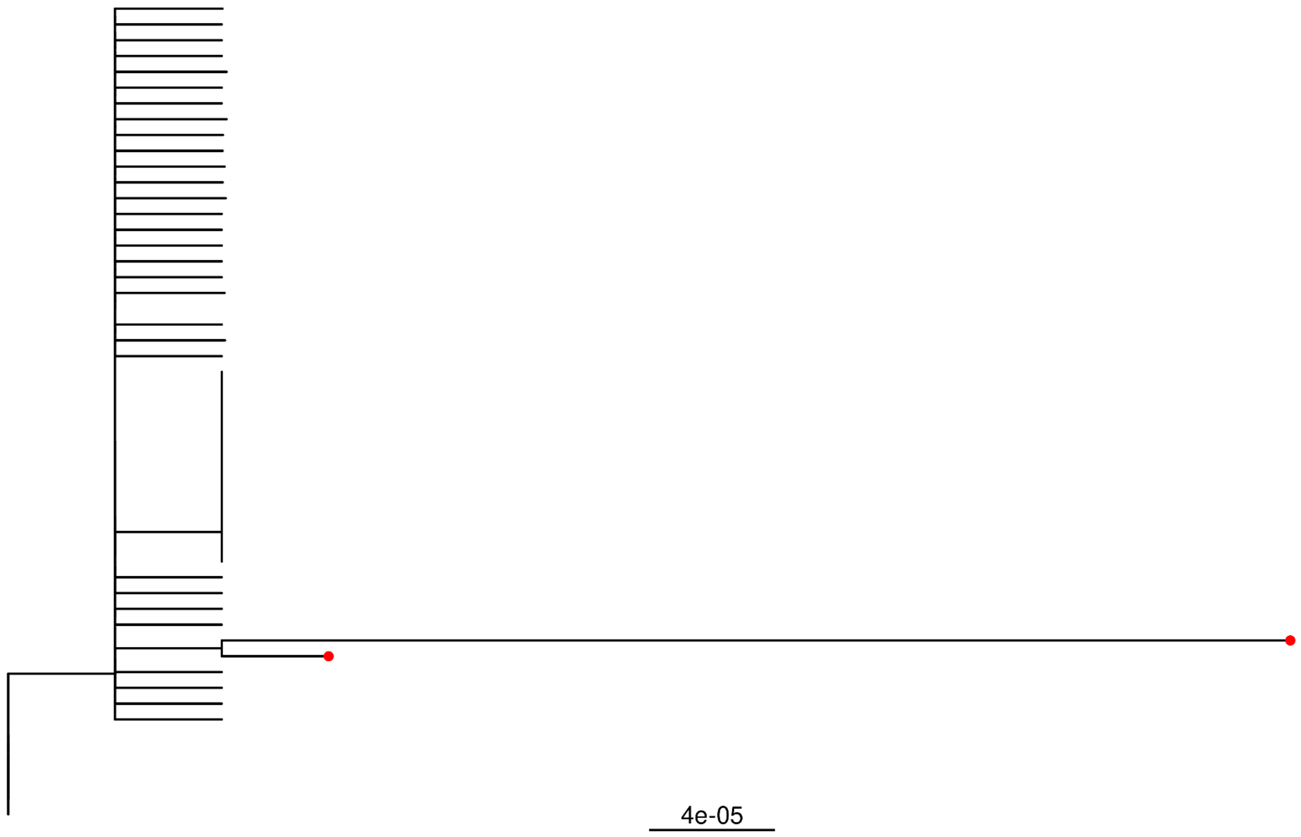

Pt-10 Civet Tree

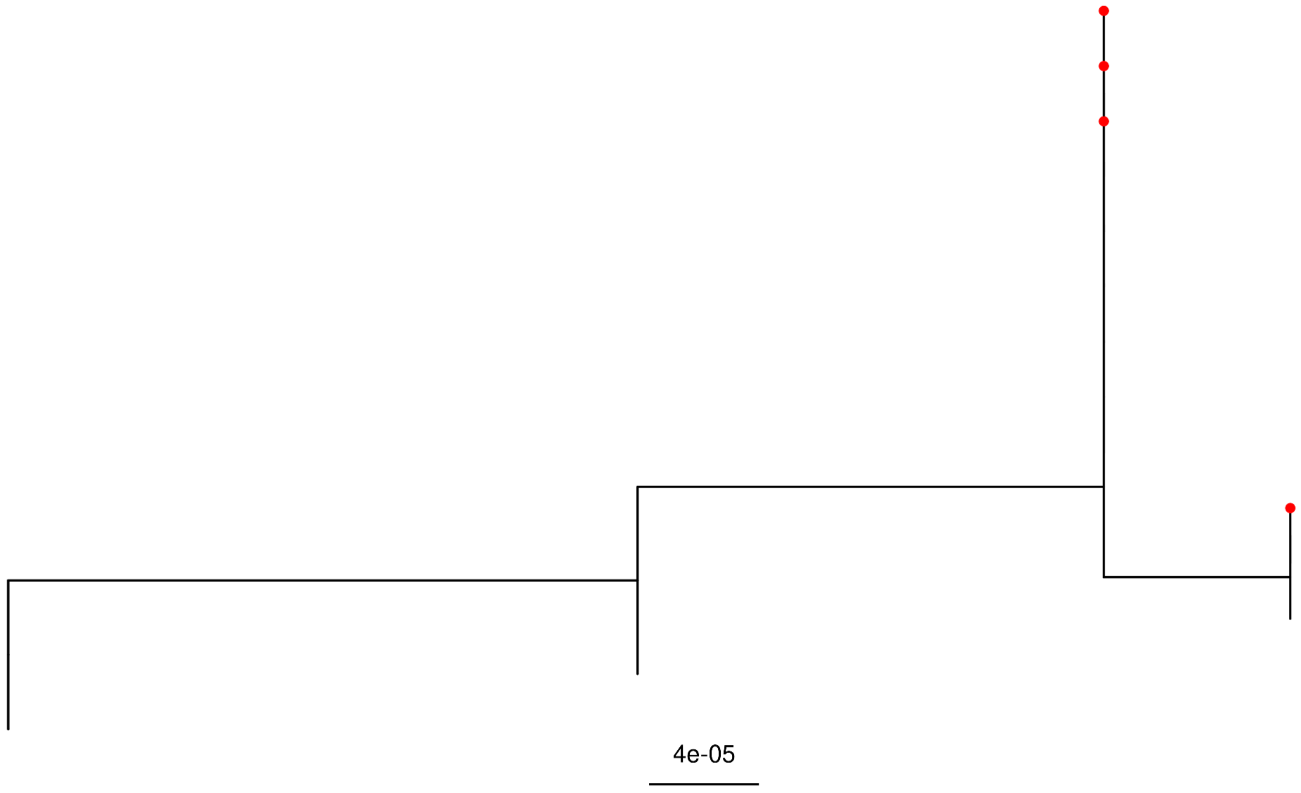


Pt-11 Civet Tree

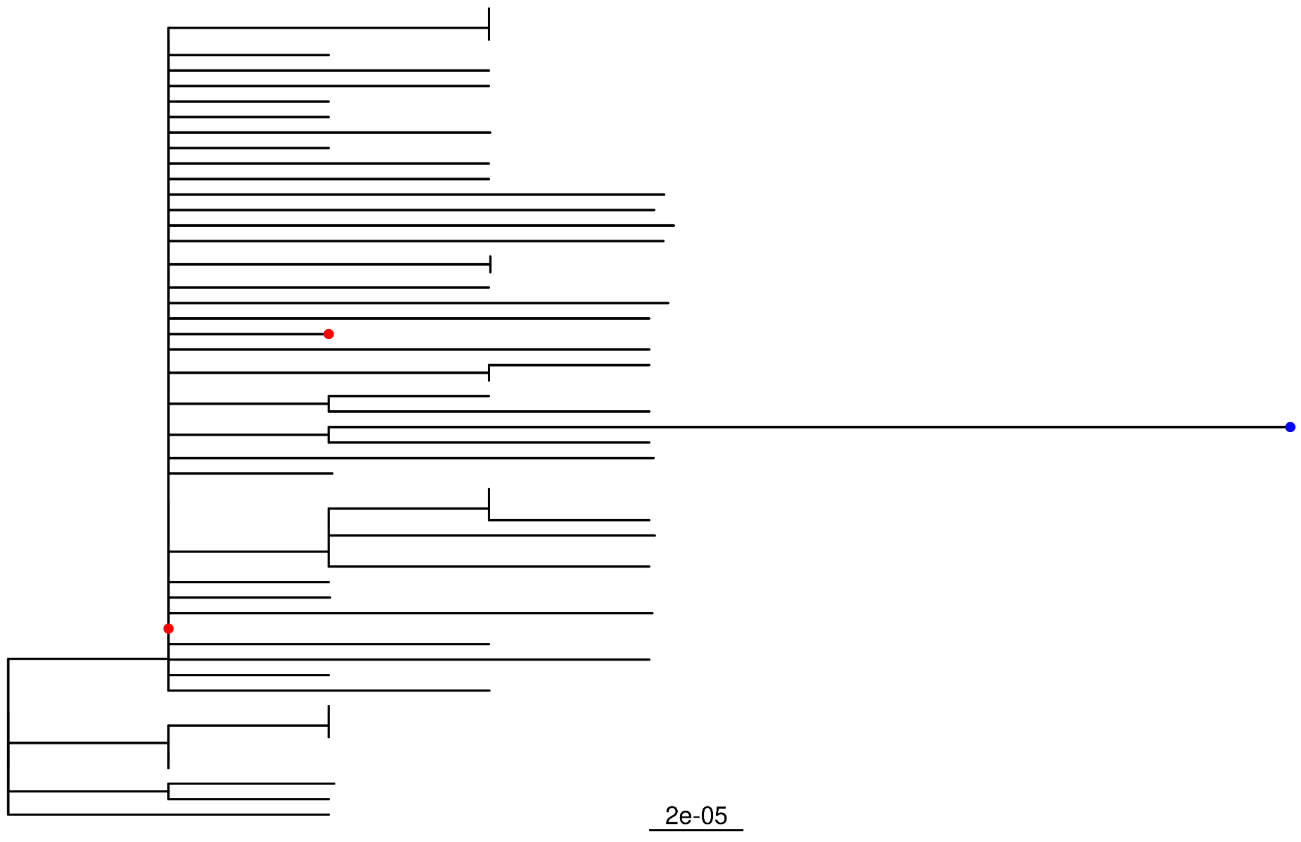

Pt-12 Civet Tree

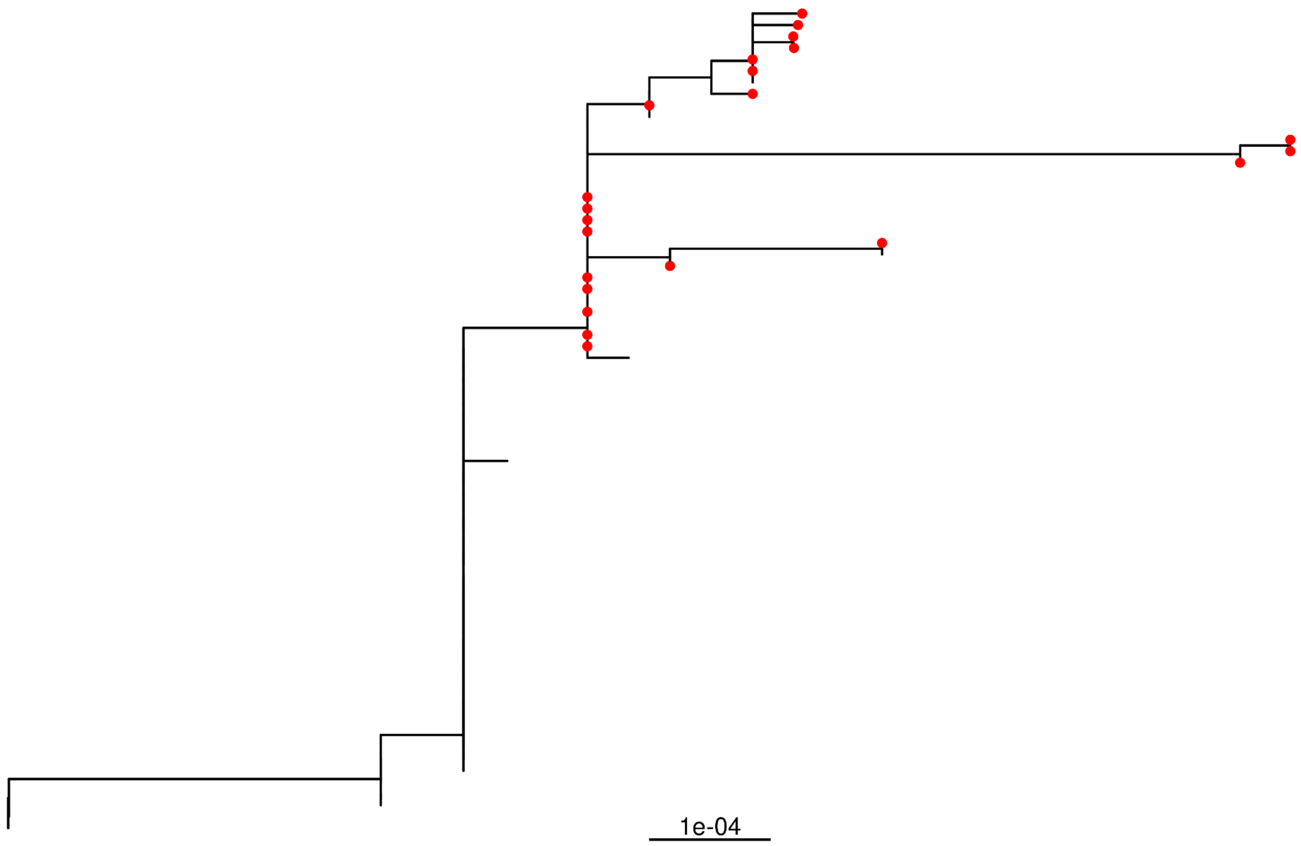


Pt-13 Civet Tree

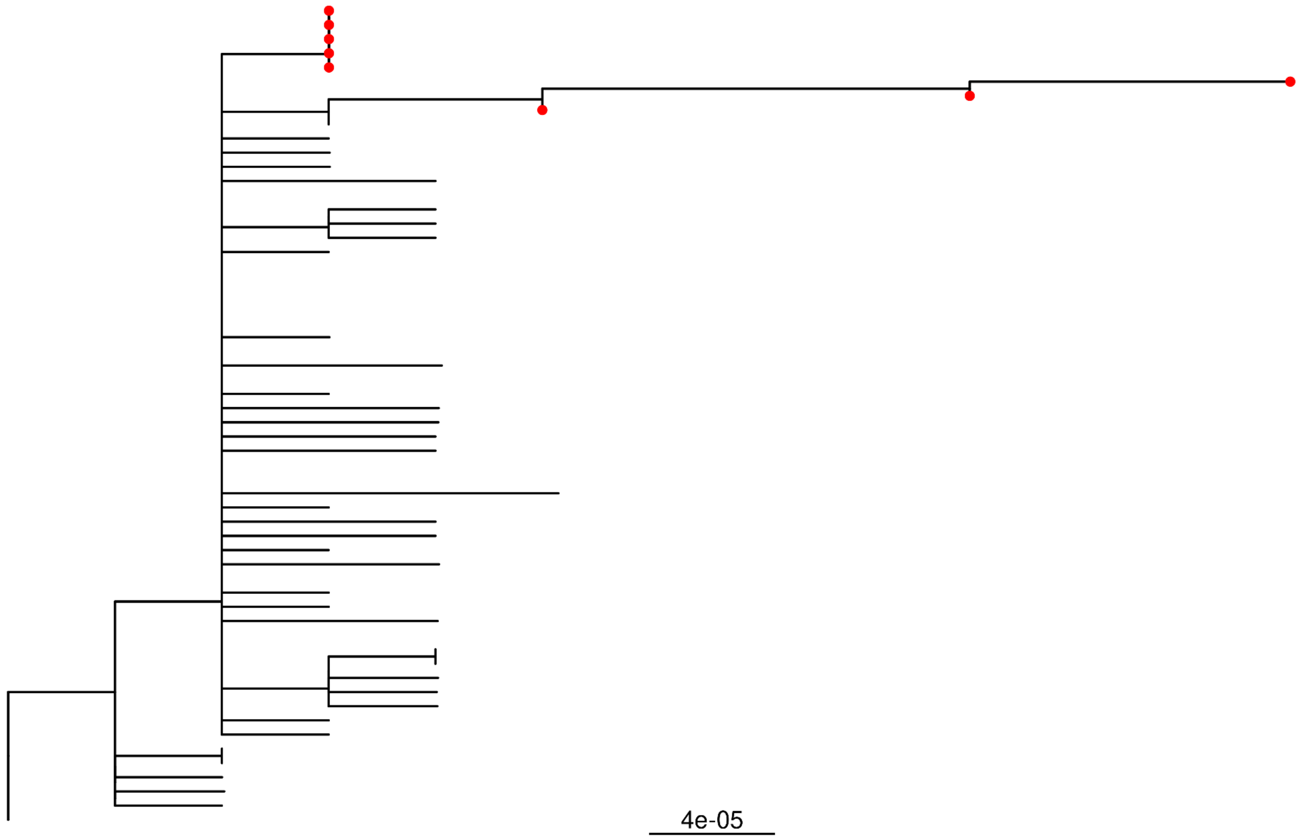

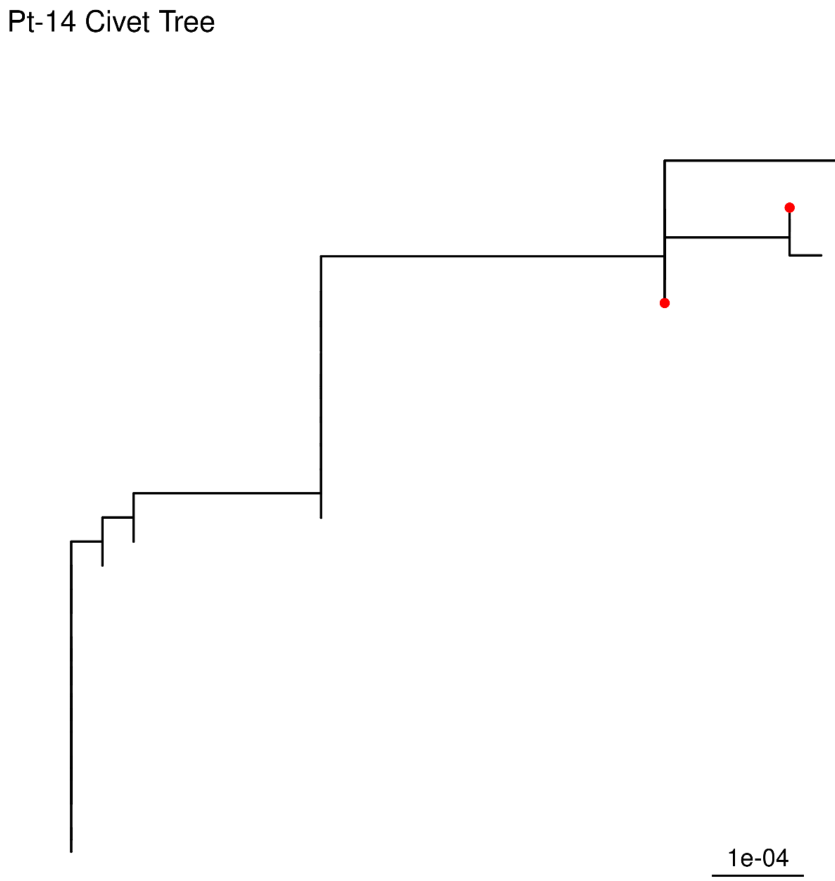
Pt-14 Civet Tree
1e-04

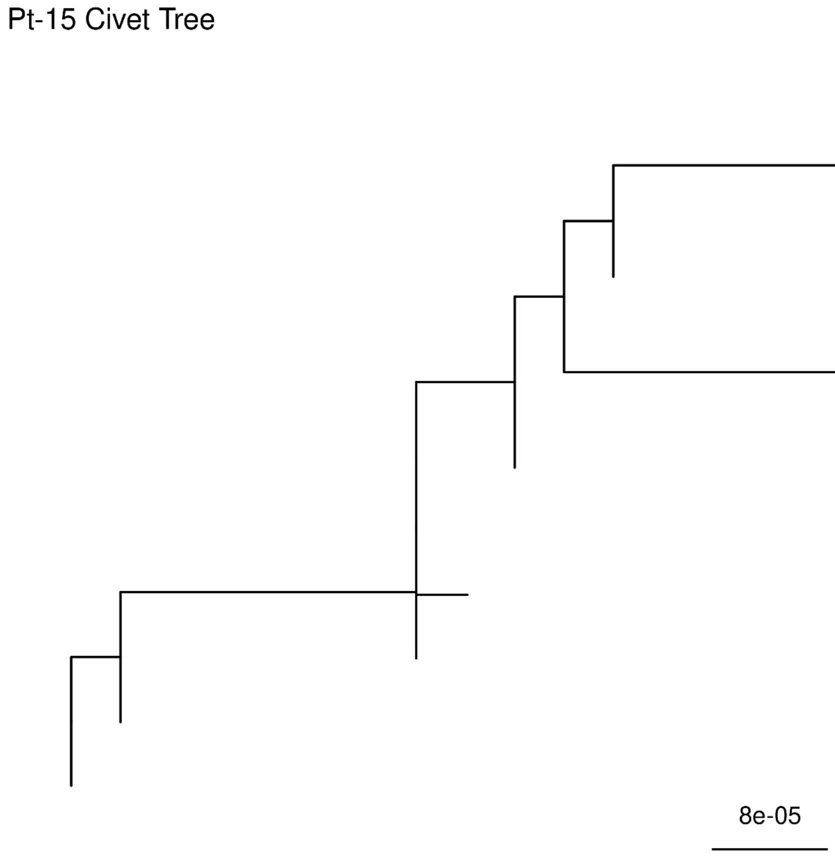
Pt-15 Civet Tree
8e-05

Pt-16 Civet Tree

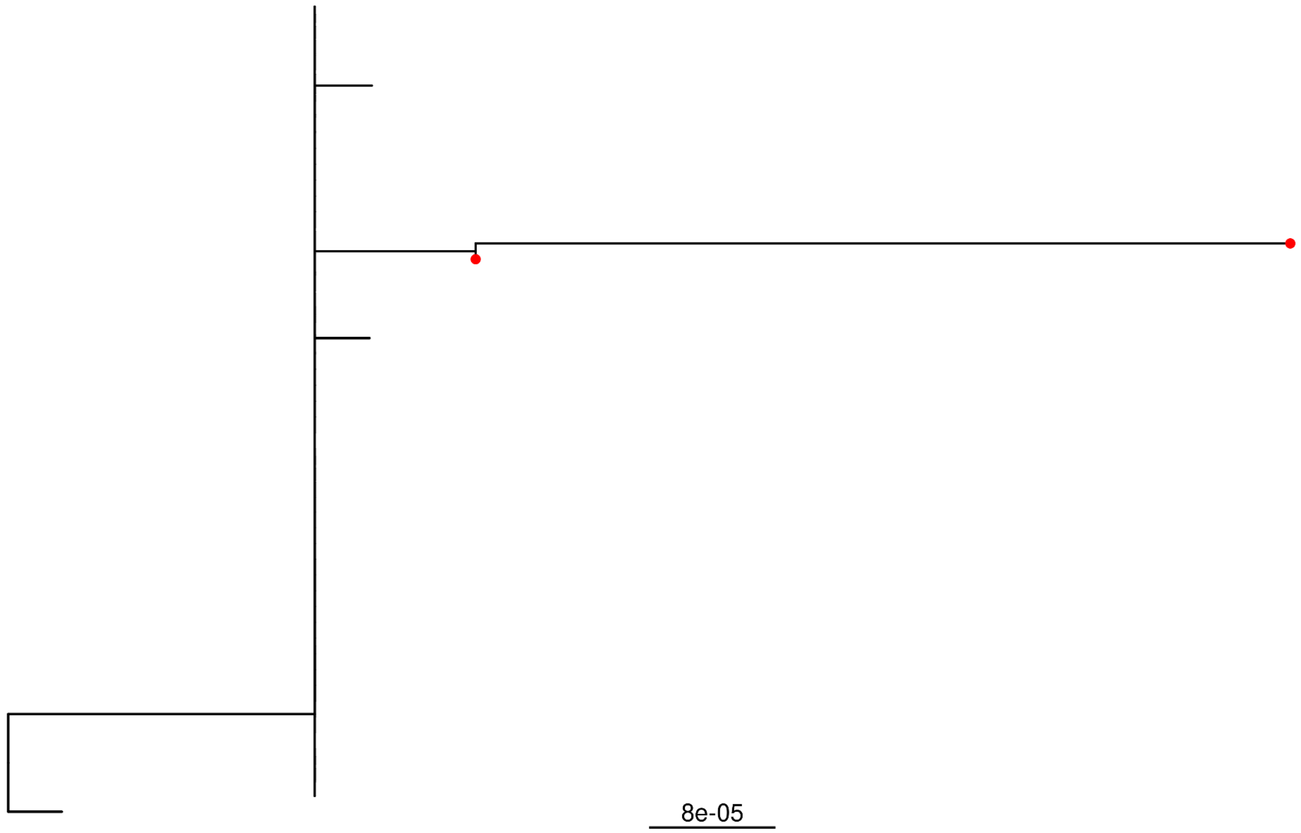


Pt-17 Civet Tree

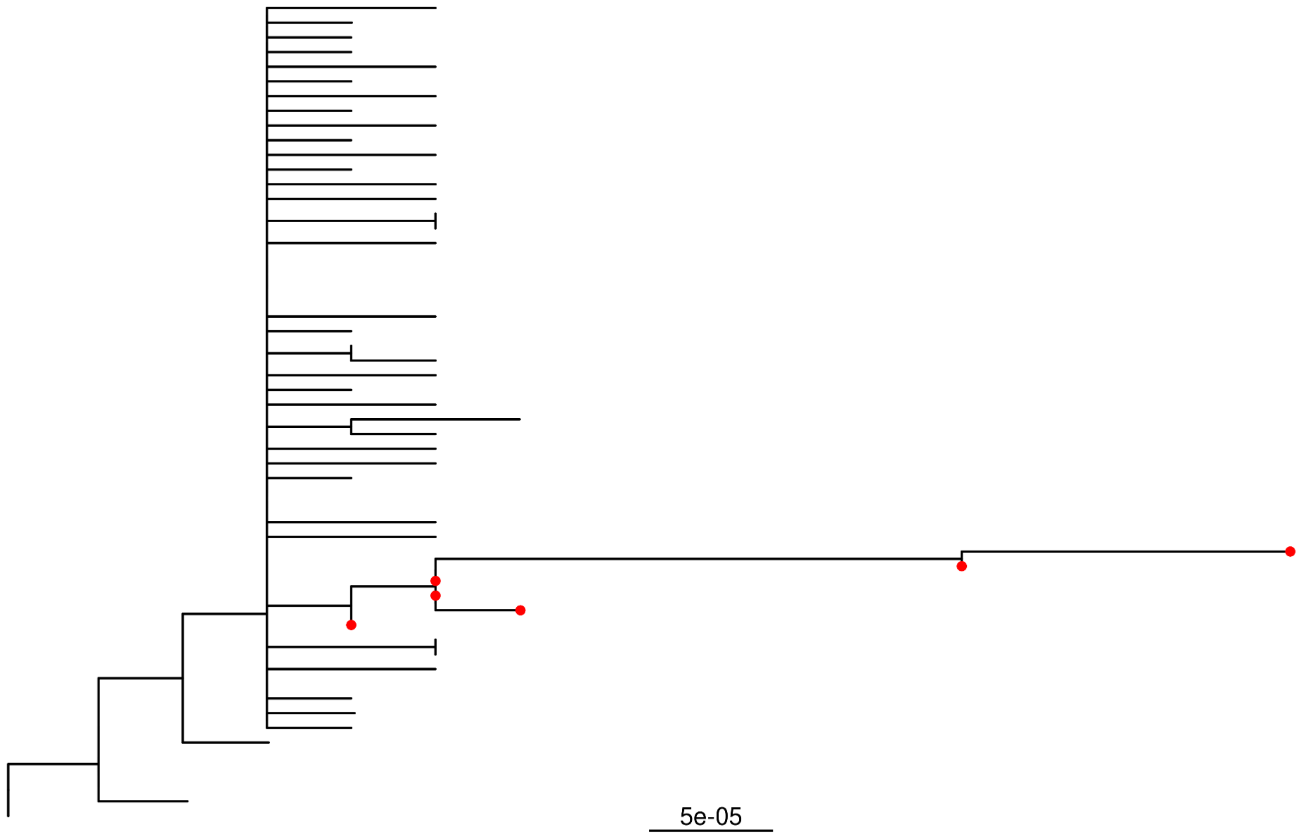

Pt-18 Civet Tree

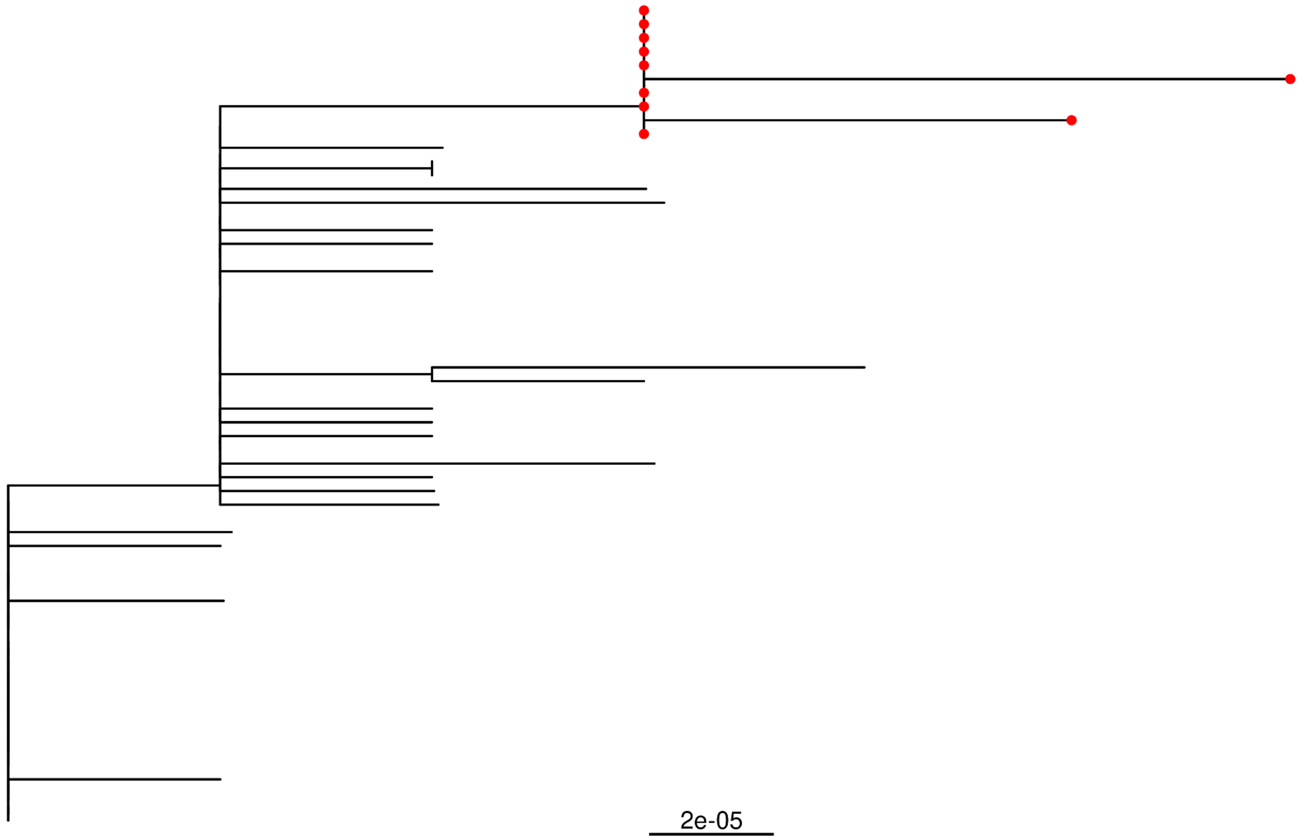


Pt-19 Civet Tree

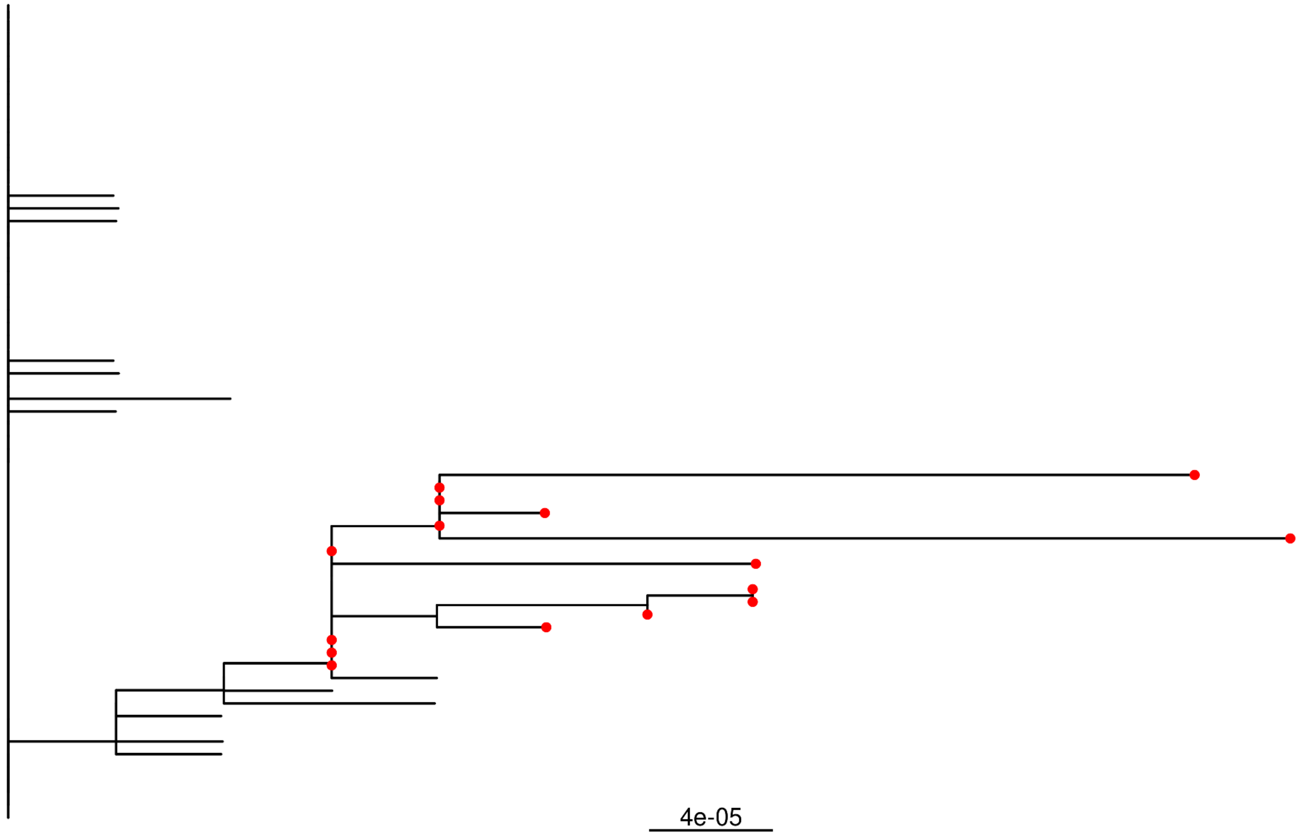

Pt-20 Civet Tree

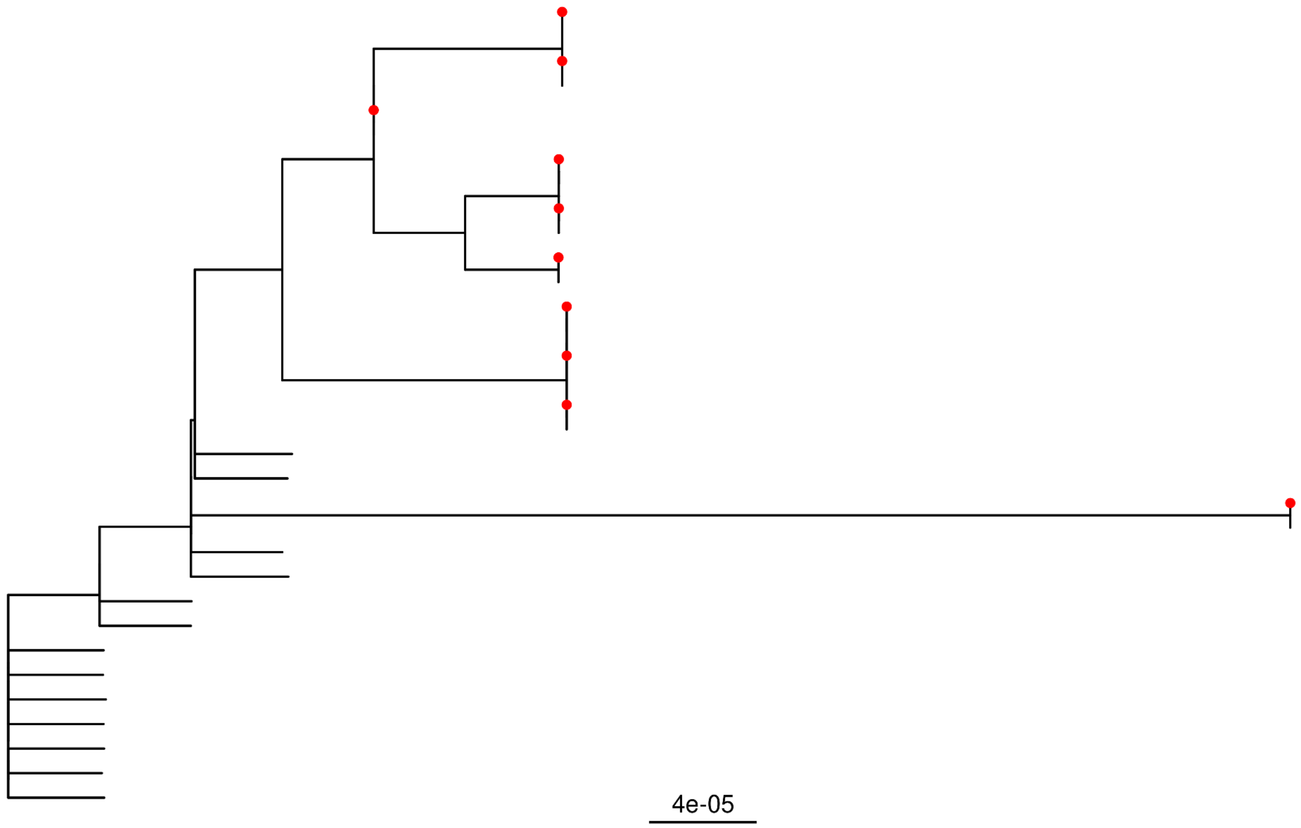


Pt-21 Civet Tree

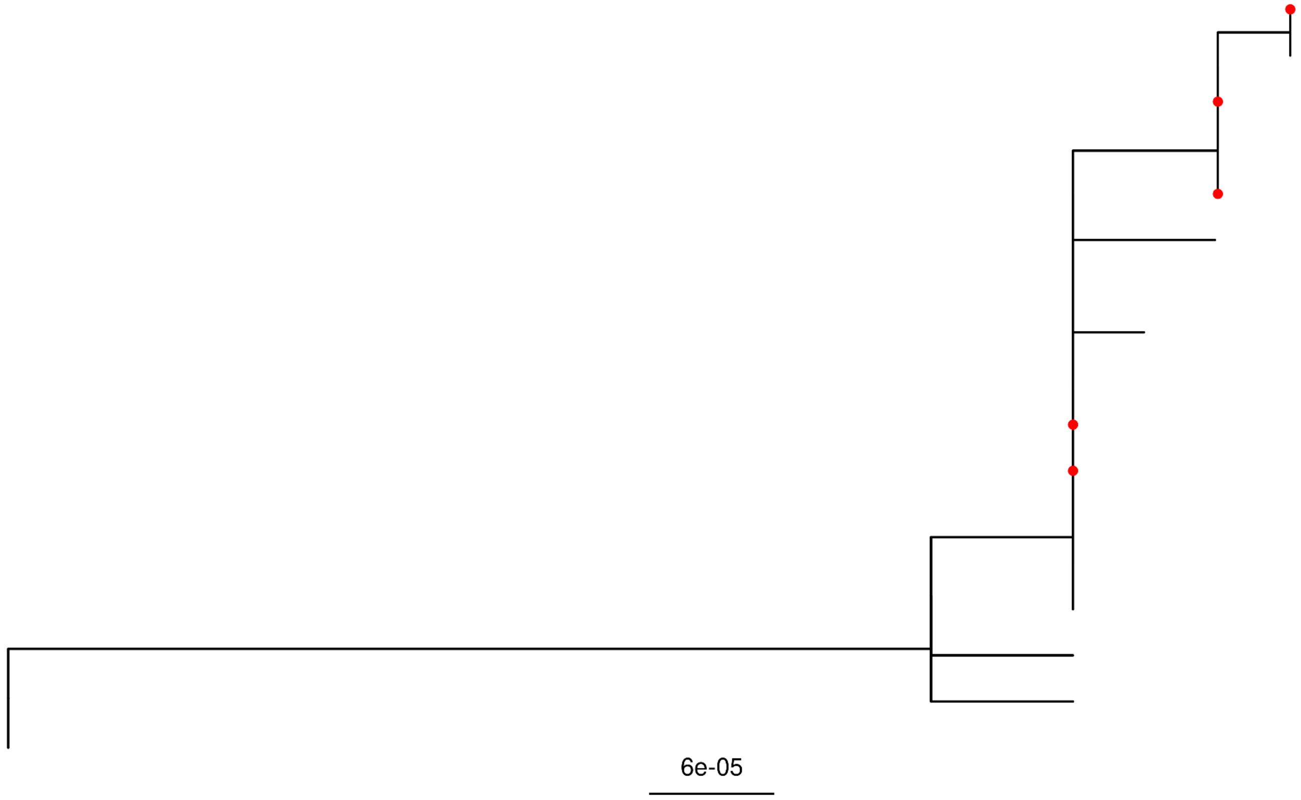

Pt-22 Civet Tree

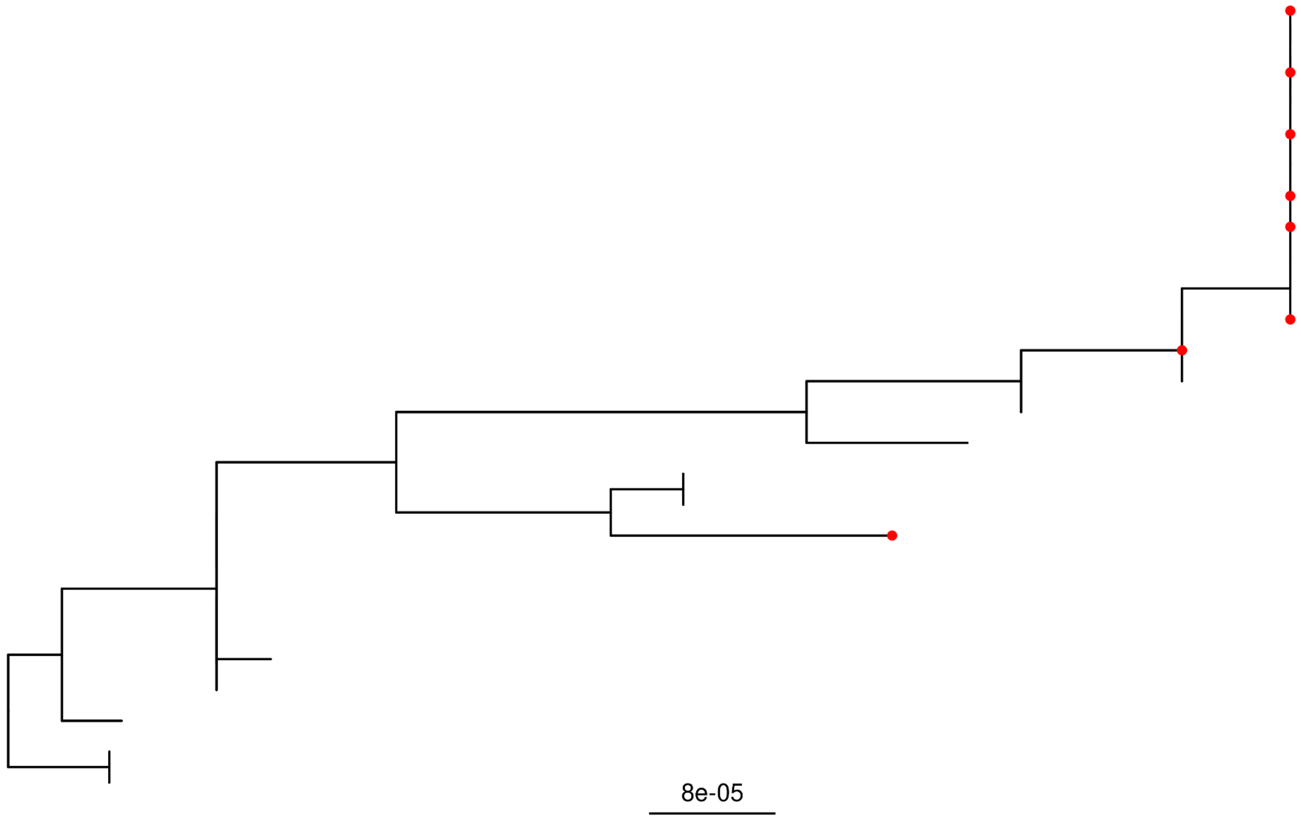


Pt-23 Civet Tree

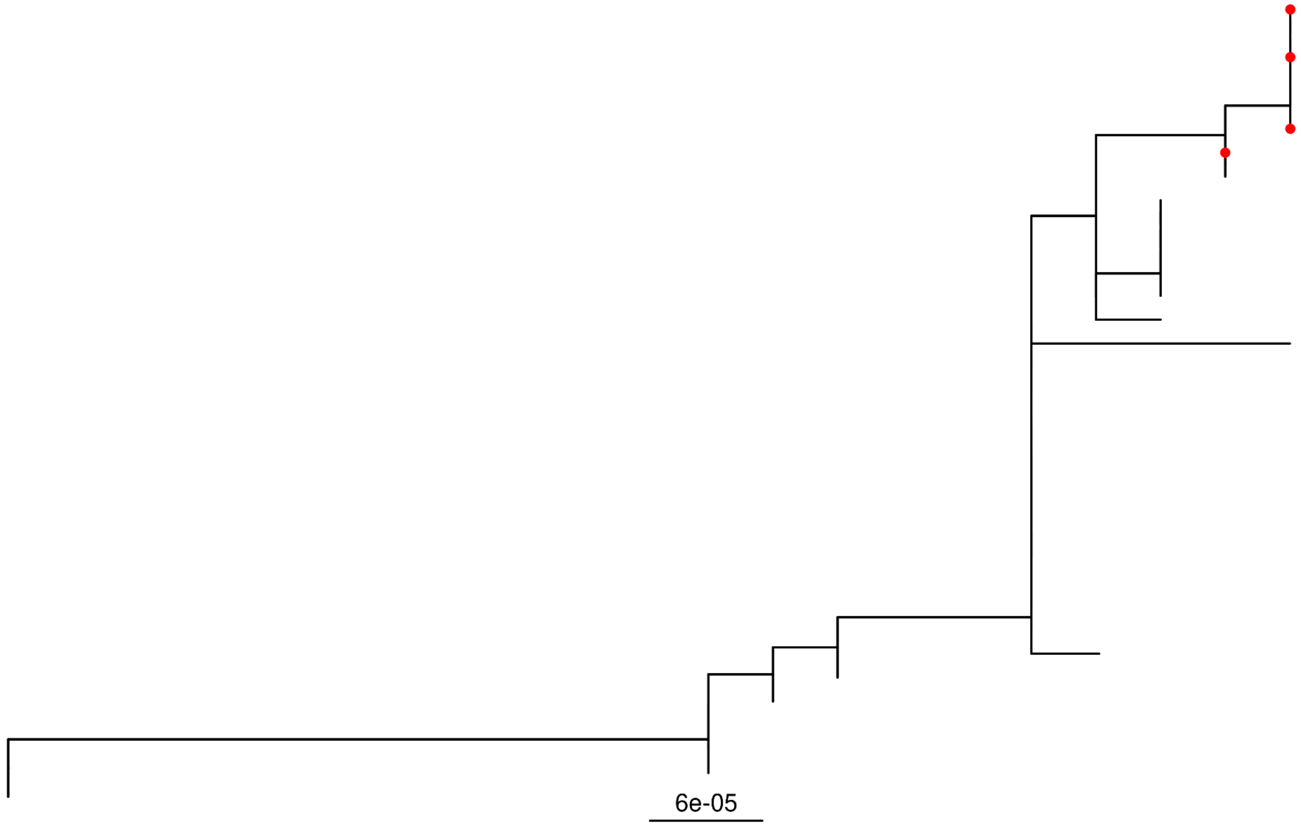

Pt-24 Civet Tree

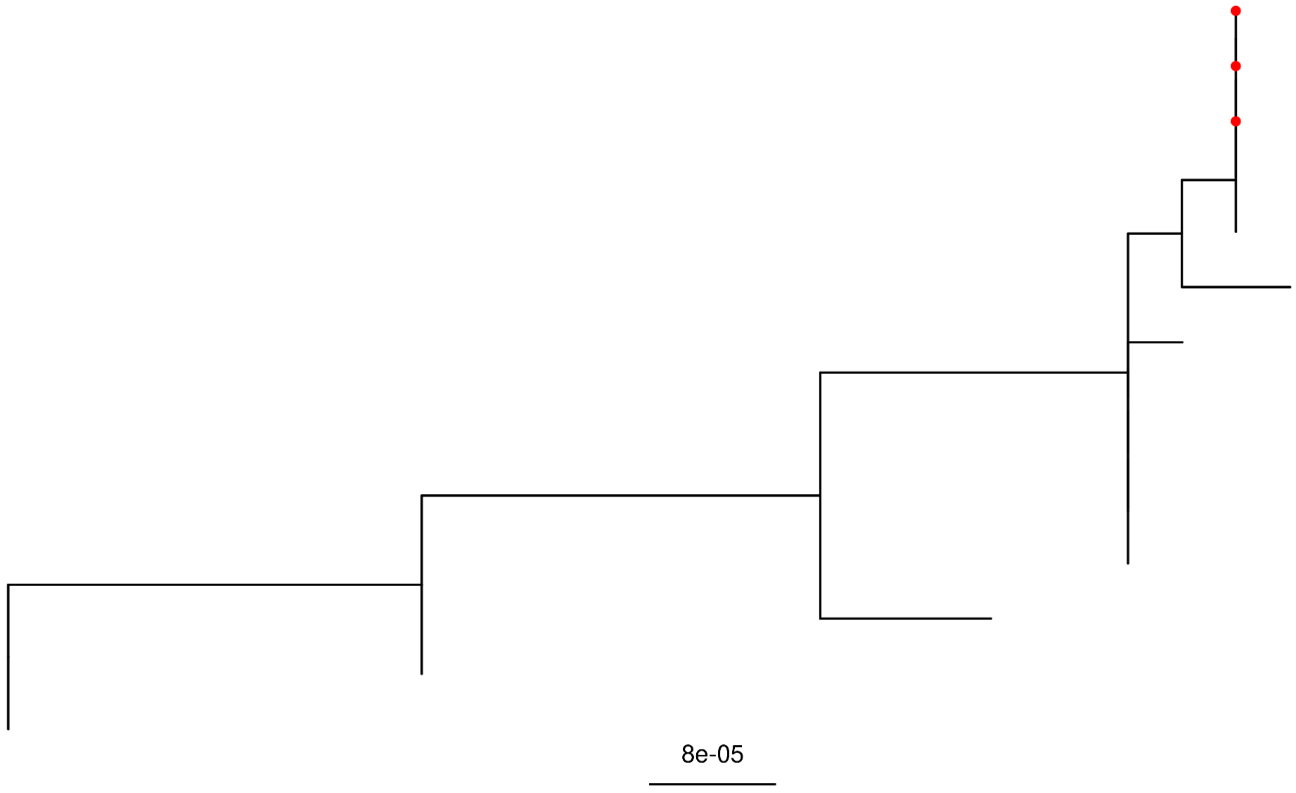


Pt-25 Civet Tree

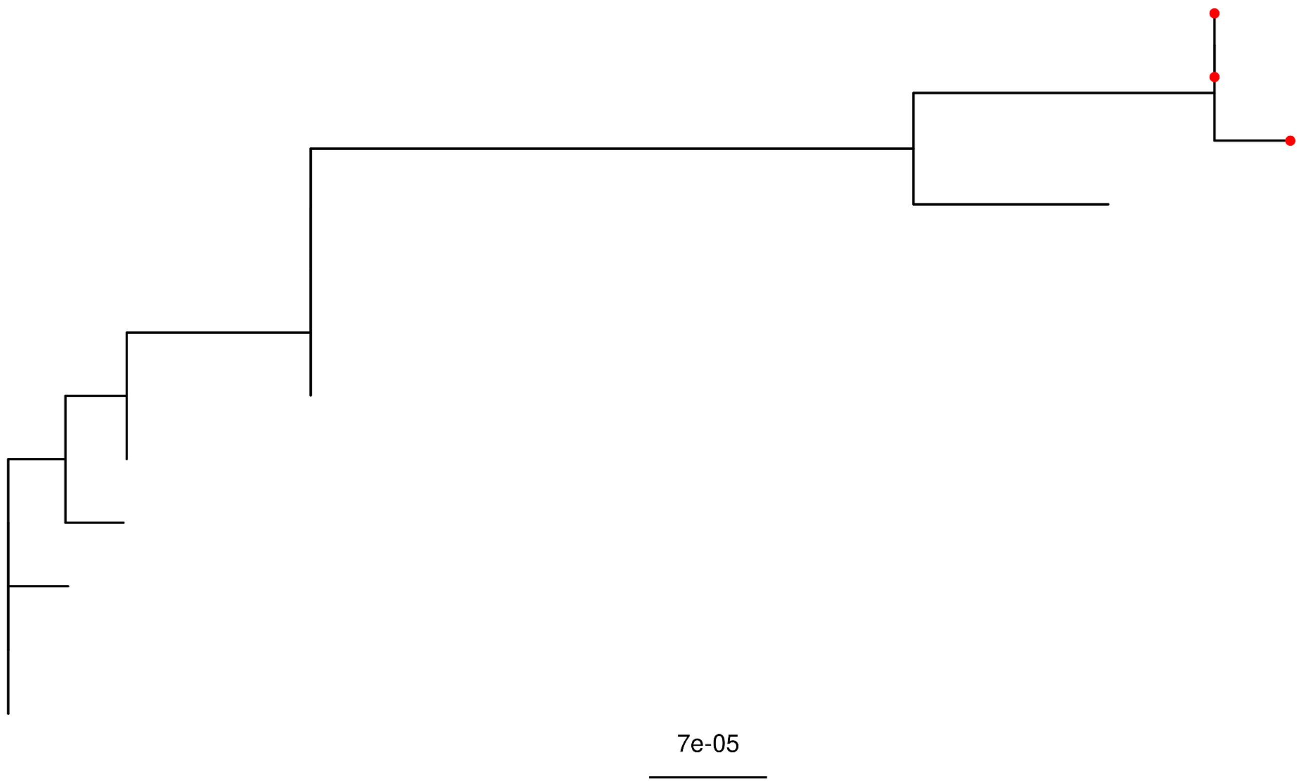

Pt-26 Civet Tree

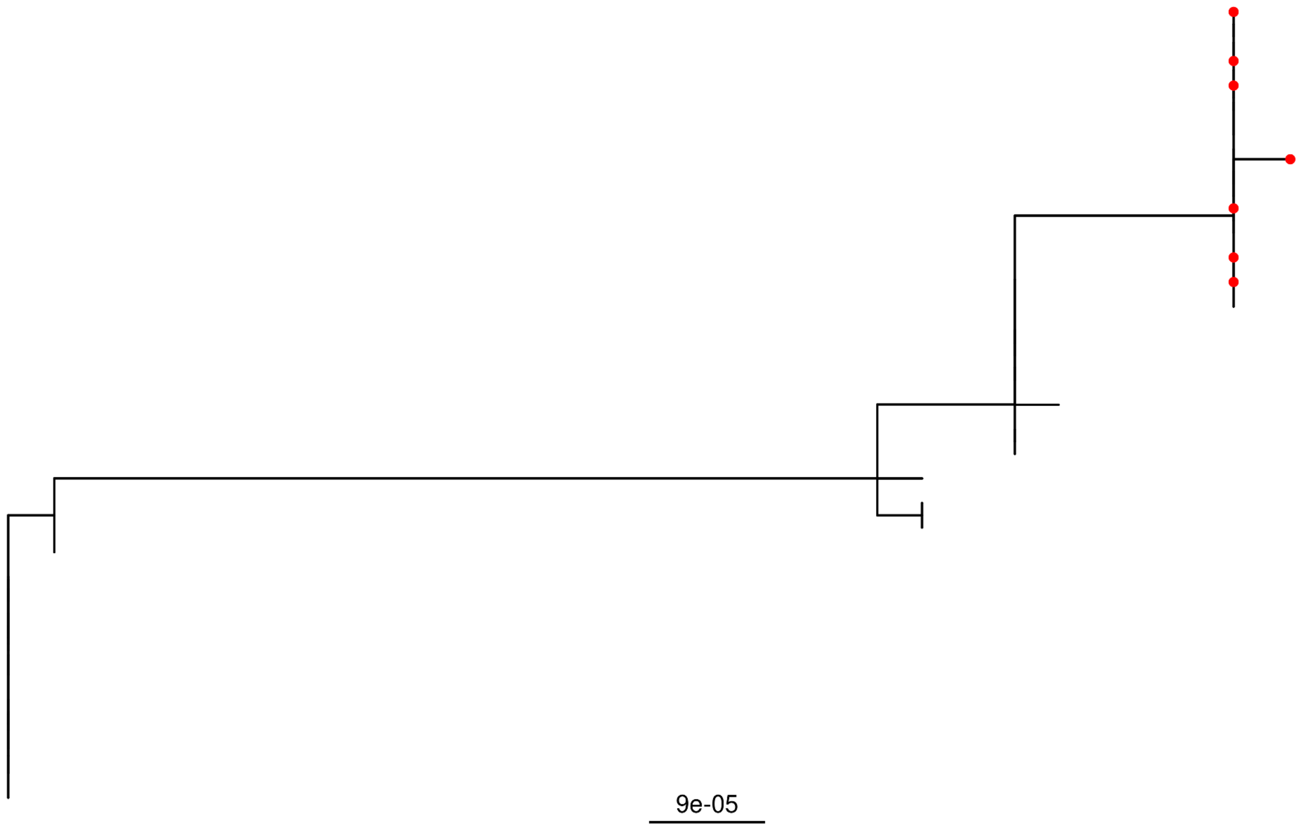

Pt-27 Civet Tree

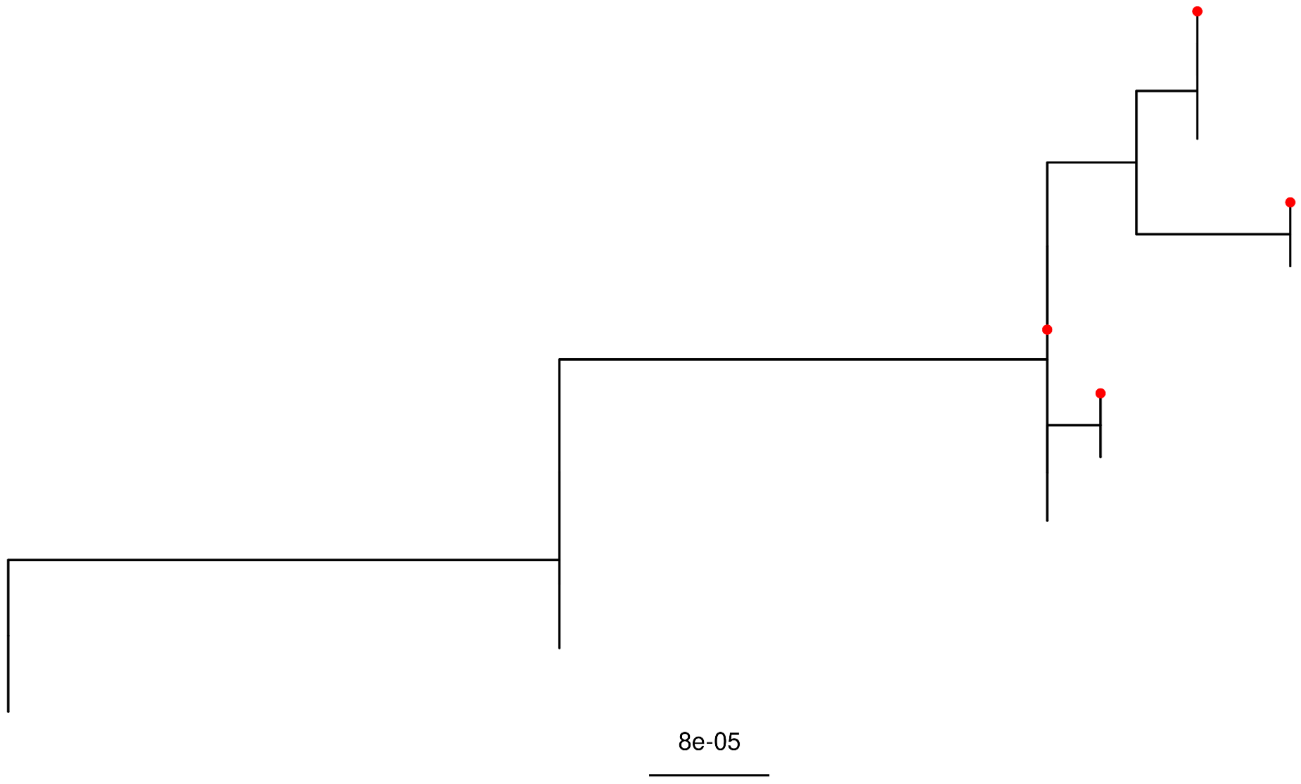


Pt-28 Civet Tree

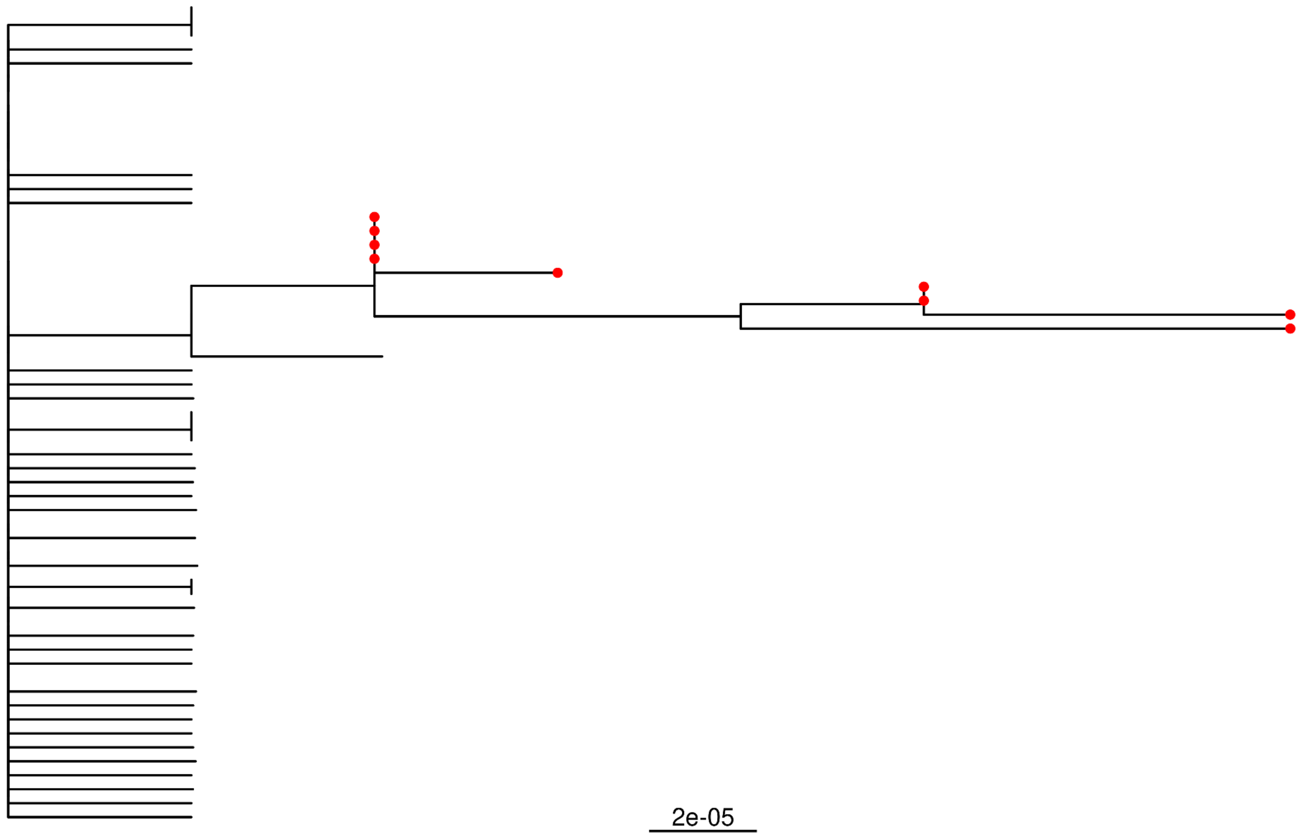
